# Targeting ATP2B1 impairs PI3K/Akt/Fox-O3 signaling and reduces SARS-COV-2 replication *in vivo*

**DOI:** 10.1101/2022.09.02.22279542

**Authors:** Pasqualino de Antonellis, Veronica Ferrucci, Francesca Bibbo, Fathemeh Asadzadeh, Francesca Gorini, Angelo Boccia, Carmen Sorice, Roberto Siciliano, Roberta Russo, Immacolata Andolfo, Vito Alessandro Lasorsa, Sueva Cantalupo, Giovanni Paolella, Giovanna Fusco, Maurizio Viscardi, Sergio Brandi, Bianca Maria Pierri, Pellegrino Cerino, Vittoria Monaco, Dong-Rac choi, Jae-Ho Cheong, Maria Monti, Achille Iolascon, Stefano Amente, Mario Capasso, Hong-Yeoul Kim, Massimo Zollo

**Author notes:** corresponding authors Correspondence (M.Z.) (H.Y.K.). co-first (Pasqualino de Antonellis, Veronica Ferrucci). co-second (Francesca Bibbo, Fathemeh Asadzadeh).

## Abstract

ATP2B1 is a known regulator of calcium (Ca^2+^) cellular export and homeostasis. Diminished levels of extra- or intra-cellular Ca^2+^ content have been suggested to block SARS-CoV-2 replication. Here, we demonstrate that a newly nontoxic caloxin-derivative compound (PI-7) inhibits ATP2B1, reduces the extra- and intra-cellular Ca^2+^ levels and impairs SARS-CoV-2 replication and propagation (VOCs: Delta and Omicron 2), as also measured by inhibition of syncytia *in vitro*. Furthermore, a FOXO3 transcriptional site of regulation of expression at the 5’ end of the *ATP2B1* locus, together with a rare homozygous intronic variant in the *ATP2B1* locus (rs11337717; chr12:89643729, T>C), are shown to be associated with severity of COVID19 (symptomatic *versus* asymptomatic patients). Here, we identify the mechanism of action during SARS-CoV-2 infection, which involves the PI3K/Akt signaling pathway, inactivation of FOXO3 (i.e., phosphorylation), and inhibition of transcriptional control of both membrane and reticulum Ca^2+^ pumps (ATP2B1 and *ATP2A1* [i.e., SERCA1], respectively). The pharmacological action of compound PI-7 on sustaining both *ATP2B1* and *ATP2A1* expression reduces the intracellular cytoplasmic Ca^2+^ pool and thus negatively influences SARS-CoV-2 replication and propagation. As compound PI-7 shows a lack of toxicity, its prophylactic use as a therapy against the COVID19 pandemic is here proposed.

**In brief:** De Antonellis et al. shows the importance of the Ca^2+^ channel pump ATP2B1 in the regulation of extracellular and intracellular Ca^2+^ levels that positively influence SARS-CoV-2 replication in human cells. Our study identifies the mechanism of action of SARS-CoV-2 in the regulation of the expression of *ATP2B1* and *ATP2A1* loci during infection via FOXO3 transcriptional factor. Furthermore, a small caloxin-derivative molecule (compound PI-7) can inhibit ATP2B1 activity, thus resulting in SARS-CoV-2 impairment. In further support, we have identified a genetic variant within the noncoding upstream region of *ATP2B1* in symtomatic patients affected by severe COVID19, thus indicating this polymorphism as a genetic predisposition factor to SARS-CoV-2 infection.

**Highlights:**

1. An anti-viral model of network of action for ATP2B1 against SARS-CoV-2 at the intracellular level that involves the PI3K/Akt signaling pathway, inactivation (i.e., phosphorylation) of FOXO3 and its transcriptional control, and inhibition of both membrane and reticulum Ca^2+^ pumps (i.e., ATP2B1, ATP2A1, respectively).
2. A new drug and its lack of toxicity “compound PI-7”, thus envisioning both preventive and therapeutic applications in patients with COVID-19.
3. The specificity of action in the context of Ca^2+^ homeostasis is one of the strategies that coronaviruses (including SARS-CoV-2 and any new VOC, including Omicron 2) use to infect host cells and promote organ dysfunction.
4. Therapeutic applications for compound PI-7 against all other viruses belonging to the Coronoviridae family (e.g., SARS-CoV, MERS-CoV), and against the main families of positive sense ssRNA viruses from other hosts (e.g., Nidovirales), as these are all Ca^2+^ dependent.
5. Identification of a rare homozygous intronic variant in the *ATP2B1* locus (rs11337717; chr12:89643729, T>C) that is associated with severity of COVID19 (i.e., symptomatic *versus* asymptomatic patients). This variant can be used as a marker to identify those patients that might show severe COVID19 following their SARS-COV-2 infection.

## Introduction

The coronavirus disease 2019 (COVID19) pandemic is one of the worst crises of our times, which prompted the urgent need to uncover the mechanisms that have pivotal roles with severe acute respiratory syndrome coronavirus 2 (SARS-CoV-2). SARS-CoV-2 uses multiple approaches to infect its host (Baggen et al., 2021), to evade the host responses that are still poorly understood (Xie and Chen, 2020). SARS-CoV-2 infection shows a wide range of clinical features, which range from asymptomatic to mild and severe, which mainly depends on both host genetic factors and virus–host interactions.

Coronaviruses (CoVs) contain positive-sense, single-stranded RNA (∼30 kb). Four major categories have been reported, with alphaCoV and betaCoV known to infect humans. Those that can replicate in the lower respiratory tract cause pneumonia, which can be fatal (Tay et al., 2020); they include SARS-CoV, Middle East respiratory syndrome-CoV, and the new SARS-CoV-2. This last CoV belongs to the betaCoV genus (Andersen et al., 2020) and has resulted in pandemic acute respiratory syndrome in humans (i.e., COVID19 disease). This can progress to acute respiratory distress syndrome, generally around 8 to 9 days after symptom onset (Huang et al., 2020). Like the other respiratory CoVs, SARS-CoV-2 is transmitted via respiratory droplets, with possible fecal–oral transmission (Huang *et al*., 2020).

When SARS-CoV-2 infects host cells, according to the discontinuous transcription mechanism its full-length positive-sense genomic RNA (gRNA) is used to produce both full-length negative-sense RNA copies (–gRNAs) and subgenomic negative-sense RNAs (– sgRNAs) that act as templates for the synthesis of positive gRNA and sgRNA, respectively. Among those sgRNAs, four encode the structural viral proteins Spike (S), Envelope (E), Membrane (M) and Nucleoprotein (N) (Kim et al., 2020; V’Kovski et al., 2021). To date, the most recognized receptor used by SARS-CoV-2 to enter host cells is angiotensin-converting enzyme 2 (ACE2), which is mainly expressed in lung and intestine, and to a lesser extent in kidney, heart, adipose, brain, and reproductive tissues (Lukassen et al., 2020; Walls et al., 2020; Wrapp et al., 2020), together with the cellular serine protease TMPRSS2. Binding to the host ACE2 receptor is mediated by the viral S protein, which consists of two noncovalently associated subunits: the S1 subunit that binds ACE2, and the S2 subunit that anchors the S protein to the membrane. The S2 subunit also includes a fusion peptide and other machinery necessary to mediate membrane fusion upon infection of a new cell. Furthermore, ACE2 engagement by the virus exposes an additional site internal to the S2 subunit, termed the S2′ site; following ACE2-mediated endocytosis, the S2’ site is cleaved by the transmembrane protease serine 2 (TMPRSS2) at the cell surface, or by cathepsin L in the endosomal compartment (Jackson et al., 2022). This “priming” process triggers the fusion of the viral envelope with cellular membranes, thereby allowing the release of the viral genome into the host cell (Hoffmann et al., 2020; Jackson *et al*., 2022). Despite data showing that ACE2 is a high-affinity receptor for SARS-CoV-2 (Lan et al., 2020), several lines of evidence have suggested that other factors are involved in the priming process.

Calcium (Ca^2+^) signals have long been known to have an essential role during the viral cycle (i.e., virion structure formation, virus entry, viral gene expression, posttranslational processing of viral proteins, and virion maturation and release [Zhou et al., 2009]). The role of Ca^2+^ in virus–host cell interactions has been shown for various types of envelope viruses (e.g., Rubella virus [Dube et al., 2014], Ebola virus [Nathan et al., 2020]), including CoVs (Berlansky et al., 2022). In this regard, the depletion of extra- and/or intra-cellular Ca^2+^ pools was shown to significantly reduce the infectivity of SARS-CoV, thus suggesting that both the plasma membrane and endosomal cell entry pathways (Lai et al., 2017) are regulated by Ca^2+^. Of importance, recent studies have also provided evidence that the use of Ca^2+^ channel blockers (e.g., amlodipine, nifedipine) can reduce mortality from COVID19 (Crespi and Alcock, 2021), thus further underlining the importance of Ca^2+^ in SARS-CoV-2 infection and replication.

Intracellular and organellar Ca^2+^ concentrations are tightly controlled via various pumps, including the calcium pumps (Ca^2+^ ATPases). Among the plasma membrane Ca^2+^ pumps, the plasma membrane Ca^2+^ ATPases (PMCAs) are ATP-driven Ca^2+^ pumps that are ubiquitously expressed in the plasma membrane of all eukaryotic cells. The PMCA proteins are encoded by four genes (*ATP2B1-4*) with numerous splice variants that modulate their tissue distribution, cellular localization, and functional diversity (Krebs, 2015). Among the *ATP2Bs* genes, the homozygous deletion of the *ATP2B1* gene in mice was shown to give rise to a lethal phenotype, thus suggesting that ATP2B1 has a role as the housekeeping isoform (Okunade et al., 2004) required for the maintenance of intracellular Ca^2+^. Indeed, ATP2B1 is critical for the maintenance of cytosolic Ca^2+^ concentrations below 300 nM (i.e., at ∼100 nM), due to its high affinity for Ca^2+^ (Kd, ∼0.2 M)^17,18^, and it represents the major Ca^2+^ efflux pathway in nonexcitable cells (Muallem et al., 1988), with an important role in regulation of the frequency of Ca^2+^ oscillations (Caride et al., 2001). ATP2B1 activity is regulated by the Ca^2+^ signaling protein calmodulin (CaM), which stimulates ATP2B1 activity through its binding to an autoinhibitory domain (Bruce, 2018).

Here, using gene expression analysis, we demonstrate the unbalancing of Ca^2+^ signaling pathways during SARS-CoV-2 infection *in vitro* using human HEK-293T cells overexpressing ACE2 on the plasma membrane (HEK293T-ACE2). This is mostly due to deregulation of ATP2B1 and ATP2A1 (i.e., SERCA1) proteins on the plasma membrane and the endoplasmic reticulum, respectively, thus also clarifying the role of Ca^2+^ in SARS-CoV-2 replication. In this regard, using human primary cells obtained from nasal brushing from a healthy donor, the downregulation of ATP2B1 expression levels promotes SARS-CoV-2 replication, as shown here by increased viral Nucleoprotein (N) levels in infected cells, by augmenting the intracellular Ca^2+^ levels. A model describing this mechanism of action on both *ATP2B1* and *ATP2A1* further describes the action of SARS-CoV-2 (variants of concern [VOCs]: Delta and Omicron 2) during their status of replication and infection. Here, viral entry into the host cells enhances PI3K/Akt signaling pathways, which in turn diminishes the transcriptional nuclear activity of the transcriptional factor FOXO3 by promoting its phosphorylation (Brunet et al., 1999), thus causing a reduction in the transcriptional activation of their target genes, including *ATP2B1* and *ATP2A1*. This, sequentially, causes an increase in the intracellular Ca^2+^ concentration that enhances SARS-CoV-2 replication and propagation. Furthermore, the inactivation of FOXO3 also causes an increase in the NF-κB inflammatory pathway (Thompson et al., 2015), thus further promoting the cytokine storm induced by SARS-CoV-2.

Of importance, using artificial intelligence screening, we identify a new non toxic caloxin-derivative compound (PI-7) that *(i)* inhibits ATP2B1 activity via reduction of extra-intracellular cellular Ca^2+^ levels and *(ii)* enhances *ATP2B1* and *ATP2A1* mRNA and protein levels via FOXO3 transcriptional regulation. Further, compound PI-7 also *(iii)* impairs SARS-CoV-2 (VOCs: Delta and Omicron 2) infection and propagation by negatively affecting the generation of syncytia, and *(iv)* prevents the release of inflammatory cytokines that are targets of NF-κB.

Furthermore, through the search of rare genetic variants associated with severe disease status we identified a rare (0.038187 global frequency) intronic homozygous ‘rs11337717’ polymorphisim (chr12:89643729, T>C) in *ATP2B1* locus that is positively associated with severity of COVID19.

Altogether, these data identify a new genetic factor that is responsible for severe COVID19 predisposition, and also report the potential use of a new nontoxic molecule in the fight against SARS-CoV-2 infection.

## Results

### SARS-CoV-2 infection reduces intracellular Ca^2+^ signaling via downregulation of ATP2B1 and ATP2A1

Ca^2+^ homeostasis has been reported to have an important role during SARS-CoV-2 viral infection (Chen et al., 2019; Zhou *et al*., 2009). Here, to study the physiological response of ATP2B1 to Ca^2+^ oscillations in the presence of SARS-CoV-2, we generated HEK293T cells that stably express its main receptor, ACE2 (i.e., HEK293T-ACE2 cells; Ferrucci et al., 2022). As a control, we used immunoblotting analyses to verify that overexpression of ACE2 does not alter the subcellular localization of the ATP2B1 protein (Figure 1A). These data show the presence of ATP2B1 protein only in the membranes enriched fraction obtained from HEK293T-ACE2 cells (and in their total protein lysates, as the positive control) (Figure 1A).

**Figure 1.**
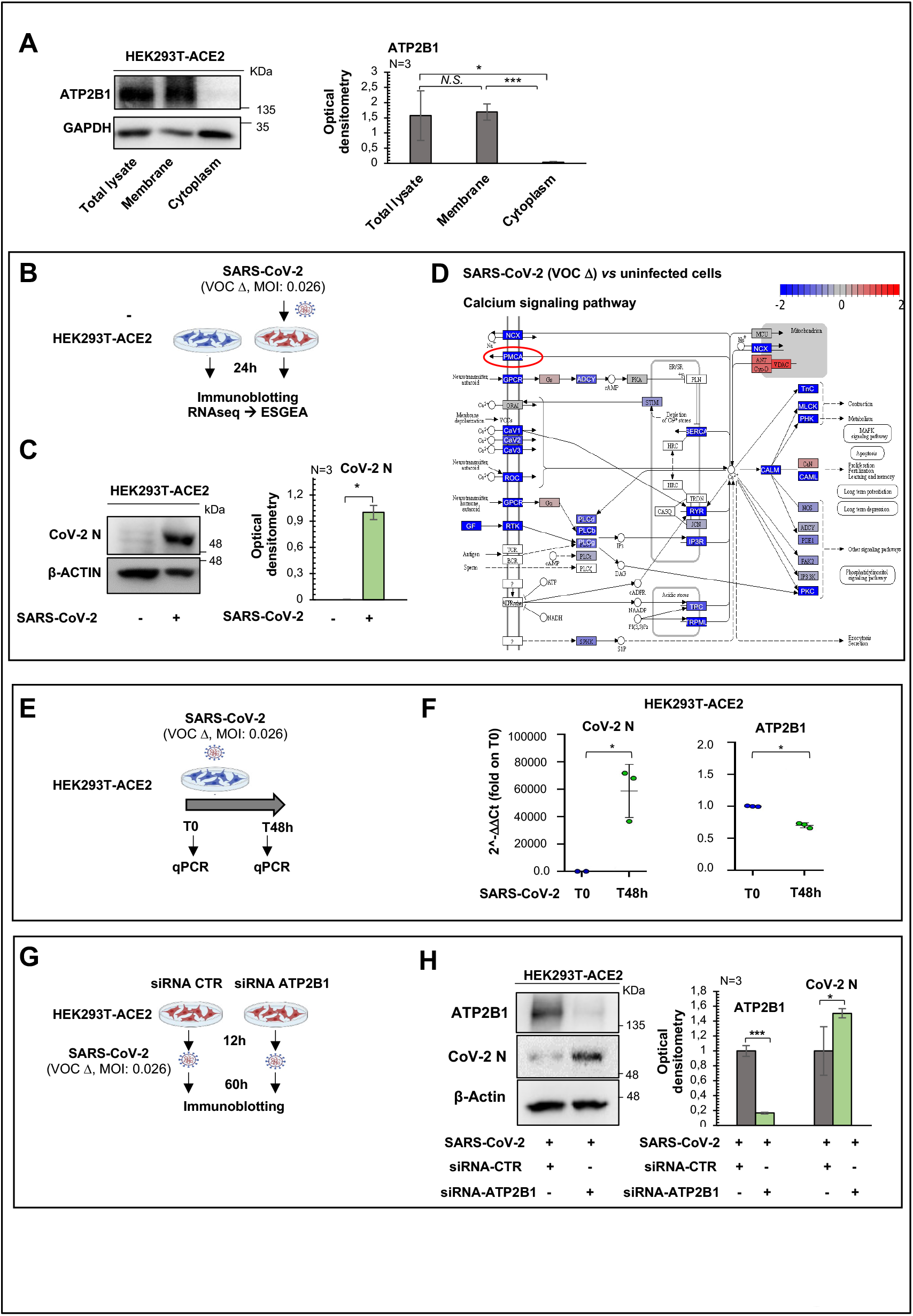
Dysregulated expression of Ca^2+^ pumps, including ATP2B1, during SARS-CoV-2 infection. **(A)** Left: Representative immunoblotting analysis of cytosolic, membrane, and total protein lysate fractions obtained from HEK293T-ACE2 cells, using antibodies against the indicated proteins. β-Actin was used as the loading control. The experiment was performed in triplicate. Right: Densitometry analysis of the ATP2B1 band intensities from blots from three independent experiments. Data are means ±SD. * p <0.05; *** p <0.0001; NS, not significant (unpaired two-tailed student’s *t*-tests; N = 3 independent experiments per group). **(B)** Experimental design for the HEK293T-ACE2 cells infected with SARS-CoV-2 (VOC Delta at 0.026 MOI) for 24 h. Noninfected cells cells were used as control. **(C)** Left: Representative immunoblotting analysis using antibodies against the indicated proteins on total protein lysates obtained from cells treated as above. Noninfected cells were used as control. All experiments were performed in triplicate. Right: Densitometry analysis of the viral N protein (CoV-2 N) band intensities in blots from three independent experiments. Data are means ±SD. * p <0.05, (N = 3 independent experiments per group). **(D)** The Reactome pathway for “Calcium Signaling” displayed in a Reactome diagram view obtained from the differentially expresed genes between noninfected and SARS-CoV-2–infected HEK293T-ACE2 cells (VOC Delta, 0.026 MOI, 24 h) found through RNAseq analyses. Blue, downregulated genes upon SARS-CoV-2 infection; red, upregulated genes upon SARS-CoV-2 infection; red circle, PMCA Ca^2+^ pumps (or ATP2Bs). **(E)** Experimental design showing analyses of HEK293T-ACE2 cells infected with SARS-CoV-2 (VOC Delta at 0.026 MOI) for 48 h. Total RNA was extracted at T0 and after 48 h. **(F)** Quantification of mRNA abundance relative to control cells at T0 (2^−ΔΔCt^) for the human *ATP2B1* and viral CoV-2 N genes. RT-PCR analysis of RNA extracted from HEK293T-ACE2 cells infected with SARS-CoV-2 VOC Delta for 48 h. Data are means ±SD. * p <0.05, (unpaired two-tailed student’s *t*-tests; N = 3 independent experiments per group). **(G)** Experimental design showing HEK293T-ACE2 cells downregulated for ATP2B1 (i.e., 12 h from transfection with the ATP2B1 siRNA) and then infected with SARS-CoV-2 (VOC Delta at 0.026 MOI) for a further 60 h. Control (CTR) cells were transfected with a pool of three unrelated siRNAs and then infected with SARS-CoV-2. **(H)** Left: Representative immunoblotting analysis using antibodies against the indicated proteins on total protein lysates obtained from HEK293T-ACE2 cells downregulated for ATP2B1 and infected with SARS-CoV-2. All experiments were performed in triplicate. Right: Densitometry analysis of the indicated band intensities in blots from three independent experiments. Data are means ±SD. * p <0.05, *** p <0.001 (unpaired two-tailed student’s *t*-tests; N = 3 independent experiments per group).

Then, to have a global picture of the transcriptome landscape in response to the “early phase” of SARS-CoV-2 infection and to confirm the above data of the importance of Ca^2+^ homeostasis in SARS-COV-2 infection, gene expression (i.e., RNAseq) analysis was performed in the HEK293T-ACE2 cellular model upon infection with viral particles belonging to the VOC Delta (multiplicity of infection [MOI]: 0.026) for 24 h (Figure 1B). Viral infection was confirmed through immunoblotting analyses, which showed the presence of the viral N protein in the HEK293T-ACE2–infected cells, in comparison with noninfected cells (Figure 1C). RNAseq analyses revealed downregulation of 1742 and upregulation of 34 gene transcripts in HEK293T-ACE2–infected cells (i.e., differentially expressed genes [DEGs]; Figure S1A).

Gene set enrichment analysis on these DEGs using both SARS-CoV-2–infected and noninfected cells showed downregulation of several signaling pathways, where after 24 h of infection, “Calcium signaling” was in the top 20 list (Figure S1B, Figure 1D, Table S1). Within this gene expression picture, there was downregulation of Ca^2+^ pump gene transcripts, including PMCAs on the cell membrane and SERCAs on the endoplasmic reticulum (Figure 1D).

Epithelial lung cells are the most common target of SARS-CoV-2. Acheampong et al. (Acheampong et al., 2022) used a single-cell RNA sequencing (RNA-seq) approach to show that a substantial number of the plasma membrane Ca^2+^ ATPases (PMCAs or ATP2B; members of the large family of Ca^2+^ ion pumps) are expressed here (Figure S1C). This analysis revealed that *ATP2B1* and *ATP2B4* mRNA expression levels are augmented in multiple cell types in the lung parenchyma (i.e., alveolar cells, including macrophages, and in alveolar epithelial cells type I and type II), in contrast to *ATP2B2* and *ATP2B3* (Figure S1C).

Here, we interrogated publicly available datasets obtained from single-nuclei RNA-seq for >116,000 nuclei sequenced from 19 COVID19 autopsy lungs and seven pre-pandemic controls (Melms et al., 2021) (https://singlecell.broadinstitute.org/single_cell/study/SCP1052/covid-19-lung-autopsy-samples). We found that the ATP2B1-4 (PMCAs) and SERCA pumps (*ATP2A1*-3) showed distinct fractional and dysfunctional changes across the lungs from patients who died with COVID19. Of interest, there were decreased levels of ATP2B1 and ATP2B4 in the lungs of COVID19 patients, while in contrast, the levels of ATP2A2 were increased (Figure S1D). Furthermore, there were undetectable expression levels of ATP2A1, ATP2A3, ATP2B2, and ATP2B3 in the lungs of both COVID19 patients and controls (Figure S1D). These results obtained from COVID19 patients further confirm those obtained from our *in-vitro* RNA-seq analyses, with downregulation of ATP2B4 and increased levels of ATP2A2 upon SARS-CoV-2 infection in these HEK293T-ACE2 cells (Table S1).

Among these Ca^2+^ pumps, we focused on *ATP2B1* because of its pivotal role in the maintenance of Ca^2+^ homeostasis in the cells (Muallem *et al*., 1988). Using the public single-nuclei RNA-seq data, we show that ATP2B1 was reduced in COVID19 patients (i.e., 0.507 vs 0.513 ATP2B1 expression; Figure S1D-E). However, its expression level was unchanged in the “early phase” of SARS-CoV-2 infection (i.e., after 24 h) in the HEK293T-ACE2 cells (Table S1).

To better dissect out the ATP2B1 expression levels during SARS-CoV-2 infection, we used the HEK293T-ACE2 cell model (Ferrucci *et al*., 2022). Thus, these cells were infected with viral particles belonging to VOC Delta for 48 h (MOI: 0.026) (Figure 1E). The data obtained from qPCR analyses (using SYBR-green approach) showed upregulated levels of viral N protein genes (as the positive control) and downregulated levels of ATP2B1 in the cells infected for 48 h, in comparison to the noninfected cells (fold: 0.7), thus suggesting that the decrease in ATP2B1 levels is not an “early event” during viral infection (i.e., within 24 h) (Figure 1F).

To better decipher the correlation between ATP2B1 expression levels and SARS-CoV-2 infection, we transiently downregulated ATP2B1 using a siRNA that specifically targets ATP2B1 (see ATP2B4 as control in Figure S2A-B), with a control of target specificity using an unrelated ATP2B1 siRNA sequence. In the same cellular model (i.e., HEK293T-ACE2 cells), after 12 h of transfection with siRNA, the cells were infected with viral particles belonging to VOC Delta (MOI: 0.026), for additional 60 h (Figure 1G). Immunoblotting analyses confirmed increased levels of the viral N protein in cells silenced for ATP2B1 (Figure 1H), thus indicating that reduced levels of ATP2B1 fosters SARS-CoV-2 replication.

Altogether, these data indicate that SARS-CoV-2 infection decreases the Ca^2+^ signaling pathways mainly due to downregulation of Ca^2+^ pumps on the cell membranes (i.e., ATP2B1). Of interest, decreased levels of ATP2B1 are also found in lungs from COVID19 patients (Figure S1D-E). Furthemore, reduced amounts of ATP2B1 during SARS-CoV-2 infection are shown to increase viral replication *in vitro* using the HEK293T-ACE2 cells (Figure 1F, H).

### Reduced ATP2B1 levels increase SARS-CoV-2 replication in human primary epithelial nasal cells

Here, the aim was to better dissect out any correlations between ATP2B1 expression, the intracellular Ca^2+^ pool, and the replication of SARS-CoV-2, through measurement of the intracellular Ca^2+^ levels upon overexpression or downregulation of *ATP2B1* in human primary epithelial cells obtained by nasal brushing from healthy donors (Ferrucci et al., 2021). This cellular model has been previously described (Ferrucci *et al*., 2021) and sequenced using whole exome sequence technology (EVA - EMBL-EBI; project ID: PRJEB42411; analyses: ERZ1700617). To this end, we transiently downregulated *ATP2B1* using siRNA approach (Figure S2A-B) in human primary epithelial nasal cells, with immunoblotting analyses performed 48 h after transfection was started. This showed a downregolution of *ATP2B1* expression (Fold 0.4) (Figure 2A). In contrast, ATP2B1-overexpressing cells (Fold 1.5) augment of 50% the expression of *ATP2B1* (Figure 2A).

**Figure 2.**
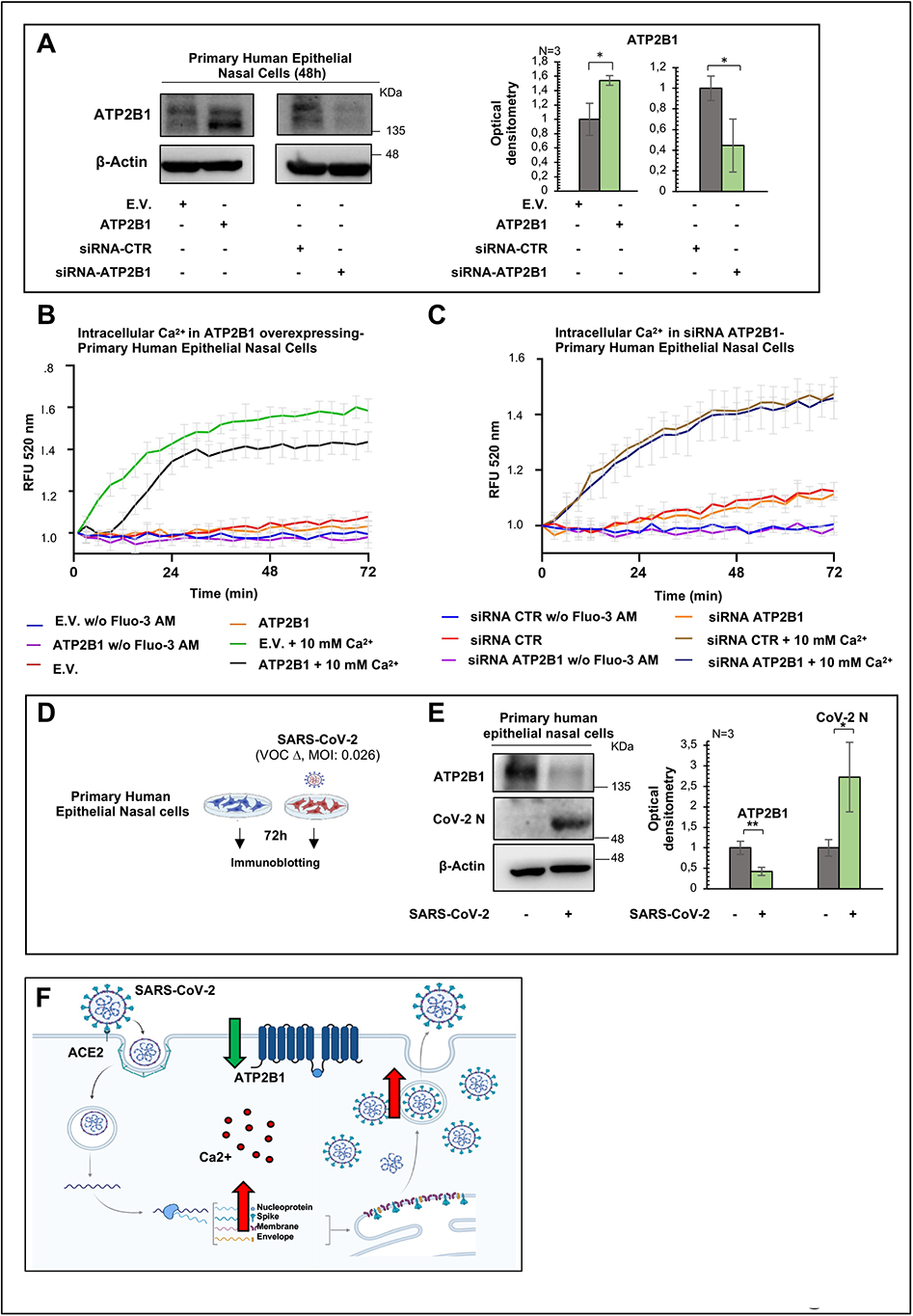
Reduced ATP2B1 protein levels promote SARS-CoV-2 replication via increasing intracellular Ca^2+^. **(A)** Left: Representative immunoblotting analysis using antibodies against the indicated proteins on total protein lysates obtained from human primary epithelial nasal cells transiently overexpressing ATP2B1 (48 h after transfection with the plasmid DNA construct pMM2-hATP2B1b) without (siRNA-CTR) and with (siRNA-ATP2B1) downregulation of ATP2B1. Cells overexpressing the empty vector (E.V.) or a pool of three unrelated siRNAs (siRNA-CTR) were used as negative controls. β-Actin was used as the loading control. All experiments were performed in triplicate. Right: Densitometry analysis of the ATP2B1 band intensities in blots from three independent experiments. Data are means ±SD. * p <0.05 (unpaired two-tailed student’s *t*-tests; N = 3 independent experiments per group). E.V., Empty Vector. **(B, C)** Quantification of relative fluorescence changes of Fluo3 as a measure of intracellular Ca^2+^ levels. Intracellular calcium (Ca^2+^) levels were measured using Fluo3-AM for up to 72 min in human primary epithelial nasal cells overexpressing ATP2B1 (48 h after transfection with the plasmid DNA construct pMM2-hATP2B1b), cells overexpressing the empty vector [E.V.] used as negative control) **(B)** and cells downregulated for ATP2B1 (48 h after transfection with the ATP2B1 siRNA; with cells transfected with a pool of three unrelated siRNAs as negative control) **(C)**. RFU, relative fluorescent units. Cells without Fluo3-AM were also used as negative control of the experiments. All experiments were performed in triplicate. N = 3 independent experiments per group. **(D)** Experimental design showing primary human epithelial nasal cells infected with SARS-CoV-2 (VOC Delta at 0.026 MOI) for 72 h. **(E)** Left: Representative immunoblotting analysis using antibodies against the indicated proteins for total protein lysates obtained from cells treated as above. Noninfected cells were used as control. All experiments were performed in triplicate. Right: Densitometry analysis of the indicated band intensities in blots from three independent experiments. Data are means ±SD. * p <0.05, ** p <0.01; (unpaired two-tailed student’s *t*-tests; N = 3 independent experiments per group). **(F)** Cartoon representation to illustrate our hypothesis for the role of ATP2B1 during SARS-CoV-2 infection. Downregulation of ATP2B1 can stimulate SARS-CoV-2 replication (increased expression of the viral N protein), via increased intracellular Ca^2+^ levels.

We then measured the intracellular Ca^2+^ levels using the Fluo3-AM substrate (as described in Star-Methods). These data showed decreased intracellular Ca^2+^ levels in ATP2B1-overexpressing human primary epithelial nasal cells (recorded for up to 72 min; Figure 2B). In contrast, the downregulation of *ATP2B1* in the same cells did not change the intracellular Ca^2+^ levels (Figure 2C). At this time, we cannot exclude that the downregulation of *ATP2B1* can be compensated by a transcriptional activation and protein synthesis of other paralog gene transcripts, which could consequentially regulate Ca^2+^ signaling. Further studies will clarify the other mechanisms responsible for Ca^2+^ homeostasis beyond *ATP2B1* downregulation.

The opposite trend between SARS-CoV-2 infection and ATP2B1 levels was then also investigated in this cellular model (i.e., human primary epithelial nasal cells). To this aim, human primary epithelial nasal cells were infected with SARS-CoV-2 viral particles belonging to VOC Delta for 72 h at 0.026 MOI, with noninfected cells used as the negative control (Figure 2D). Immunoblotting analyses showed increased viral N protein levels upon SARS-CoV-2 infection (72 h; fold: 1.5) and reduced protein levels of ATP2B1 (fold: 0.7; Figure 2E). Taken together, these data validate the opposite trend between viral infection and ATP2B1 expression at the protein level in this other cellular model. Overall, the data suggest that SARS-CoV-2 infection requires *ATP2B1* downregulation for its propagation also in the airway epithelium (Figure 2F).

### *ATP2B1* intronic polymorphism influences susceptibility for severe COVID19

As ATP2B1 has been shown to be downregulated in the lungs of COVID19 patients and here in our *in-vitro* infected cellular models (Figure S1D-E; Figure 1F), we evaluated potential genetics associations between ATP2B1 locus and the risk of developing severe COVID19. To select potential disease causative variants, we downloaded 351 coding variants of *ATP2B1* from “The Genome Aggregation Database” (GnomAD v2.1), where 13 were pathogenic according to their “Functional Analysis through Hidden Markov Models” (FATHMM) prediction scores (Table S2). However, these variants were extremely rare and were thus excluded from further investigations. The results indicate that loss of function mutations of *ATP2B1* are rare (observed/expected score = 0.08; data from GnomAD v2.1) and we postulate the hypothesis that the *ATP2B1* gene is essential for the physiological action within the cell. Further, no pathogenetic nucleotide variations within the coding protein region of ATP2B1 can be allowed for the cell to survive.

Thus, a second analysis was performed to verify the presence of noncoding variants in the genomic region of the *ATP2B1* locus that act as “expression quantitative traits loci” (eQTLs). We used an analytic approach to select candidate functional noncoding variants single nucleotide polymorphisms (SNPs; Table S3). We first selected 76 SNPs that were eQTLs for the *ATP2B1* gene (P<1 ×10^-6^) using the “Genotype-Tissue Expression” (GTex) database (Table S4). These SNPs were then annotated with prediction functional scores calculated using the “Genome-Wide Annotation of Variants” (GWAVA) tool (Table S4), and the top six SNPs were selected: rs11105352, rs11105353, rs10777221, rs73437358, rs111337717, and rs2681492. At this time we excluded rs10777221 (the most 5’ region in the extragenic region) of the *ATP2B1* locus (Figure S3A). Linkage disequilibrium (LD) analyses on the remaining SNPs showed that the only variant that is not in linkage disequilibrium was rs111337717 (Figure S3B; Figure 3A). This, thus, further indicated that this SNP deserved further analyses as a good candidate for searching for associations to COVID19 severe and asymtomatic patients.

**Figure 3.**
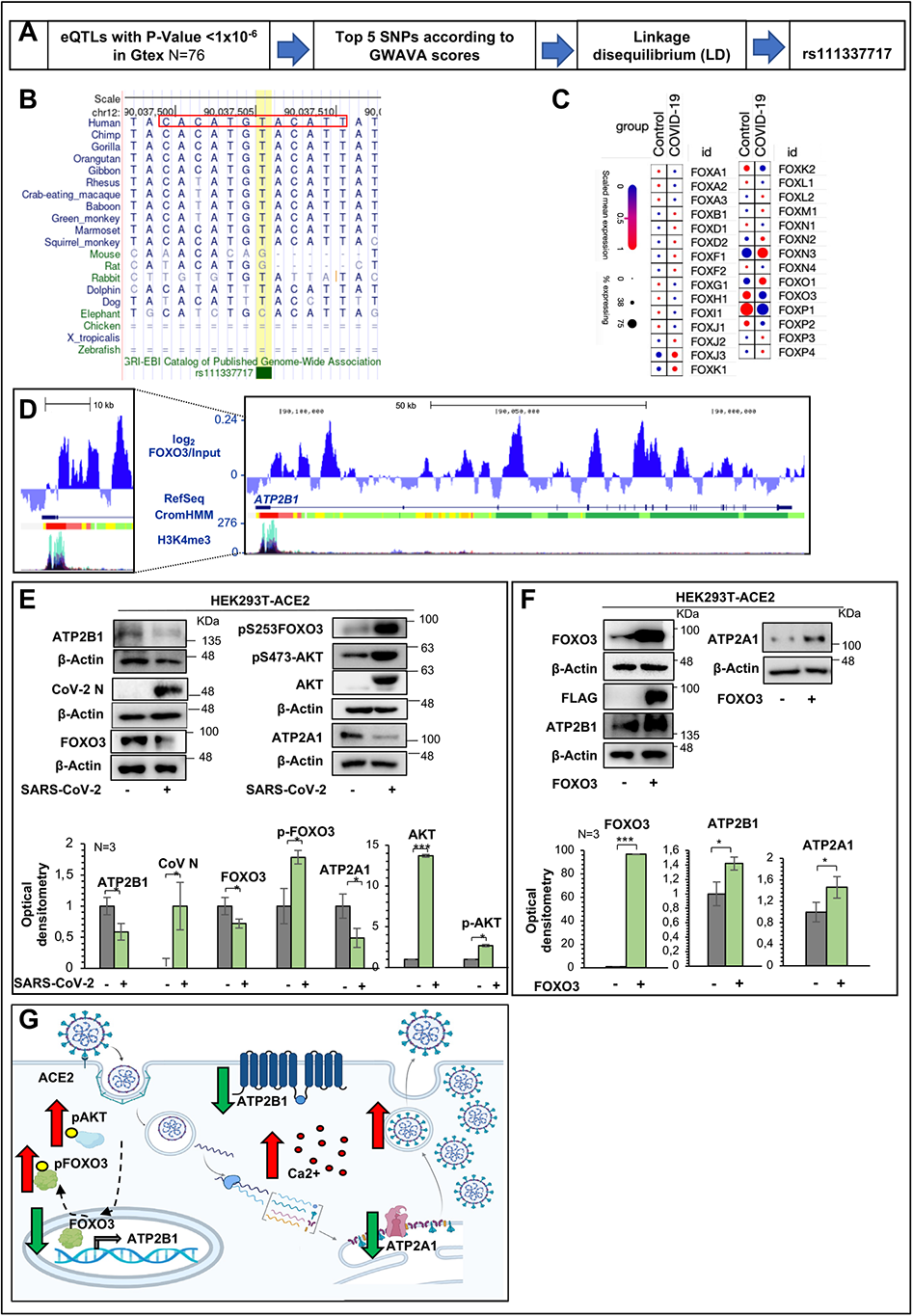
The homozygous intronic variant rs11337717 in the *ATP2B1* locus is responsible for SARS-CoV-2 increased replication in COVID patients. **(A)** *In-silico* analysis for the presence of noncoding variants in the genomic region of the *ATP2B1* locus acting as expression quantitative traits (eQTLs) located in putative elements responsible for transcriptional regulation. Here, 77 SNPs were selected that were eQTLs for the *ATP2B1* gene (P<1 ×10^-6^) using the GTex database. These SNPs were then annotated with prediction functional scores calculated by the GWAVA tool. The top five SNPs were selected to verify their linkage disequilibrium (LD). The rs111337717 SNP was not in linkage disequilibrium. **(B)** Alignment of the sequence genomic region flanking rs111337717 SNP (NC_000012.12:g.89643729T>C) SNP [(CACATG(T/C)ACATTAT)] shows the conservation through evolution across different species, with a degree of sequence identity and homology. **(C)** Literature search of publically available datasets obtained from single-nuclei RNA-seq on >116,000 nuclei from 19 COVID19 autopsy lungs and 7 pre-pandemic controls (https://singlecell.broadinstitute.org/single_cell/study/SCP1052/covid-19-lung-autopsy-samples [Melms *et al*., 2021]) to determine whether FOXOs transcriptional factors showed distinct fractional and dysfunctional changes across the lungs from the COVID19 decedents. **(D)** Genome browser screenshots showing accumulation of normalized FOXO3 signal, together with CromHMM state segmentation and the H3K4me3 signal (ENCODE), along the *ATP2B1* gene in human cells. For the CromHMM state segmentation: bright red, promoter; orange and yellow, enhancer; green, transcriptional transition; red arrow, polymorphism-containing region. The expanded view of the highlighted region (left) shows FOXO3 peaks over the *ATP2B1* promoter and enhancer regions, as marked by H3K4me3 and CromHMM (red and orange regions) respectively. **(E)** Representative immunoblotting analysis using antibodies against the indicated proteins on total protein lysates obtained from HEK293T-ACE2 cells infected with SARS-CoV-2 VOC Delta at 0.026 MOI for 72 h. Noninfected cells were used as negative control. All experiments were performed in triplicate. Densitometry analysis of the indicated band intensities in blots from three independent experiments. Data are means ±SD. * p <0.05, *** p <0.001 (unpaired two-tailed student’s *t*-tests; N = 3 independent experiments per group). **(F)** Representative immunoblotting analysis using antibodies against the indicated proteins on total protein lysates obtained from HEK293T-ACE2 cells transiently transfected with the human FOXO3 encoding plasmid for 48 h. Empty vector transfected cells were used as negative control. All experiments were performed in triplicate. Densitometry analysis of the indicated band intensities in blots from three independent experiments. Data are means ±SD. * p <0.05, *** p <0.001 (unpaired two-tailed student’s *t*-tests; N = 3 independent experiments per group). **(G)** Cartoon representation to illustrate our hypothesis for downregulation of ATP2B1 during SARS-CoV-2 infection via the FOXO3 transcriptional factor. During SARS-CoV-2 infection, the PI3K/Akt pathway is activated and enhances the phosphorylation of FOXO3, thus excluding it from the nucleus. As a consequence, the expression of the FOXO3 targets, including ATP2B1, are also reduced, thus increasing the intracellular Ca^2+^ levels and further promoting SARS-CoV-2 replication.

To this end, we tested these SNPs in a cohort of 197 patients affected by severe COVID19 and 370 asymptomatic cases (D’Alterio et al., 2022). Here, the minor allele “C” of the rs111337717 (NC_000012.12:g.89643729T>C) SNP [(CACATG(T/C)ACATTAT)] was significantly more frequent among severe COVID19 cases, when compared with asymptomatic individuals (p=0.0004; Table 1), thus suggesting that rs111337717 can be listed among the genetic risk factors for predisposition to severe COVID19. Of note, alignament sequence analyses of the genomic region flanking rs111337717 [(CACATG(T/C)ACATTAT)] showed high homology identity across different species (Figure 3B), thus suggesting a potential significant and conserved role during evolution. How this intron SNP variant influences SARS-CoV-2 infection and propagation will be an issue for future studies.

**Table 1.**
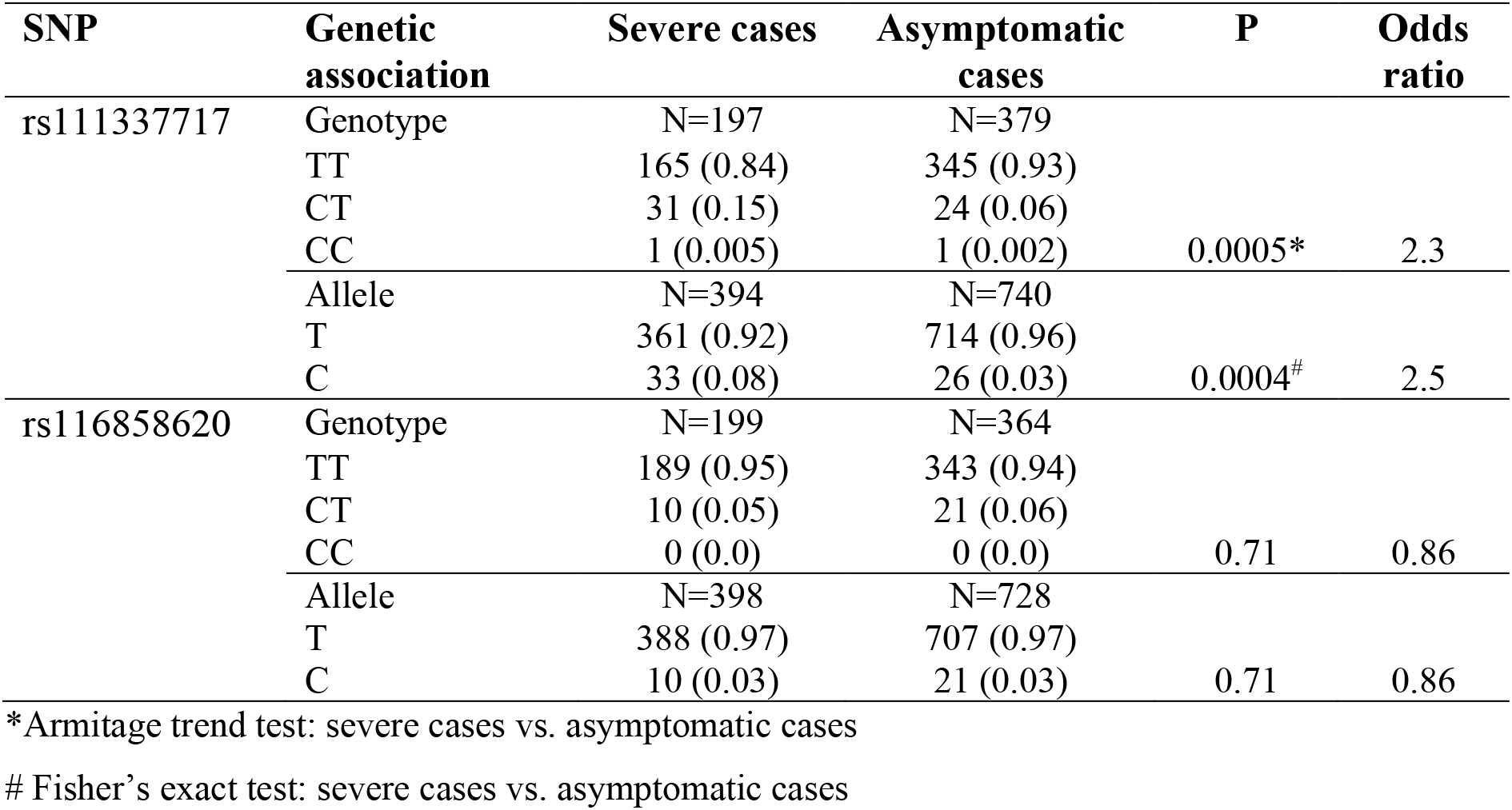
Genetic association of the rs111337717 and rs116858620 SNPs in the *ATP2B1* gene according to severe and asymptomatic cases. The rs111337717 and rs116858620 SNPs in the *ATP2B1* gene were tested in a cohort of 197 patients affected by severe COVID19 and 370 asymptomatic cases (D’Alterio *et al*., 2022). The minor allele “C” of rs111337717 (NC_000012.12:g.89643729T>C) SNP [(CACATG(T/C)ACATTAT)] is significantly more frequent among severe COVID19 cases when compared with asymptomatic individuals (p=0.0004), thus indicating that rs111337717 can be listed among the genetic risk factors for predisposition to severe COVID19.

### *ATP2B1* and *ATP2A1* are transcriptionally regulated by FOXO3 transcription factor

To determine how the locus is transcriptionally regulated, we used recent literature data showing forkhead box O (FOXO) transcription factors as strong candidates for antiviral responses against SARS-CoV-2. Their transcriptional mechanism is under epigenetic control, which in turn regulates anti-apoptotic and anti-inflammatory pathways, also acting as negative regulators of NF-κB inflammatory signaling (i.e., FOXO3) (Cheema et al., 2021).

To this end, we performed *in-silico* analysis of publicly available datasets of single-cell RNA sequencing from 19 COVID19 autopsy lungs and 7 pre-pandemic controls (Melms *et al*., 2021); https://singlecell.broadinstitute.org/single_cell/study/SCP1052/covid-19-lung-autopsy-samples). These analyses were performed to define the potential differential expression of FOXOs transcription factors in COVID19 patients. These data showed higher expression levels in the lung of FOXJ3, FOXK2, FOXN3, FOXO1, FOXO3, and FOXP1 (Figure 3C). Among these, the expression levels of FOXK2, FOXO3, and FOXP1 were lower in the lungs from COVID19 patients, compared to controls (Figure 3C), thus showing the same expression levels and trends as observed for ATP2B1 (Figure S1D-E).

The epigenetic changes regulating chromatin structure have been previously implicated in the pathophysiology of SARS-CoV-2 infection (Chlamydas et al., 2021), and, among the epigenetic drugs, the histone deacetylase inhibitor valproic acid (VPA) has been suggested to protect against the development of severe COVID19 (Collazos et al., 2022; Pitt et al., 2021). For the above reason, we have here verified which FOXO transcription factor (whose expression was found downregulated in COVID19 patients, see Figure 3C) was restored following VPA treatment. We treated HEK293T-ACE2 cells with 20 mM VPA for 16 h to investigate whether FOXO-related genes are regulated. Of note, both HEK293T-ACE2 cells and human epithelial primary nasopharyngeal cells are not characterized by SNPs in the region of the *ATP2B1* locus, as described previously (Figure S3C-D). Our qPCR data showed increased expression of FOXO3 in VPA-treated cells, compared to the vehicle control (Fold: 4.38; see Figure S3E). In contrast, the levels of *FOXK2* and *FOXP1* mRNAs were both decreased upon VPA treatment (Fold: *FOXK2*, 0.44; *FOXP1*, 0.65; see Figure S3E). Furthermore, FOXO3 also shows the same expression trend as ATP2B1, with reduced expression levels in the lung of COVID19 patients (ATP2B1, Figure S1E; FOXO3, Figure S3F).

Thus, we hypothesized that FOXO3 is a candidate transcription factor that can control ATP2B1 expression. We then used genome views of *FOXO3* ChIP-Seq signals over the *ATP2B1* locus using publicly deposited data (Figure 3D). The normalized *FOXO3* signals were visualized with the UCSC genome browser, together with layered CromHMM and H3K4me3 ENCONDE tracks, respectively. The results here indicate that FOXO3 peaks show preferential binding at the promoter and enhancer regions of both the *ATP2B1* (Figure 3D), thus indicating the potential positive transcriptional regulator activity of FOXO3.

Thus, to verify whether FOXO3 and ATP2B1 have the same downregulation trend upon SARS-CoV-2 infection *in vitro*, we infected the HEK293T-ACE2 cells with viral particles of VOC Delta (MOI 0.026) for 72 h. The immunoblotting data showed downregulated levels of ATP2B1 and FOXO3 in SARS-CoV-2 infected cells, with increased levels of phosphorylated (S253)-FOXO3 (Figure 3E). These results thus indicate increased levels of the cytosolic inactive phosphorylated-FOXO3 protein, and a reduction in the total FOXO3 and ATP2B1 protein levels following SARS-CoV-2 infection. Of interest, our data also show decreased levels of ATP2A1 upon SARS-CoV-2 infection (Figure 3E). The expression of ATP2A1 has been previously shown to be reduced upon SARS-CoV-2 infection (Figure 1D, Table S1). Of interest, the genome view of FOXO3 ChIP-Seq signals over the *ATP2A1* locus indicated that FOXO3 peaks also showed binding at the promoter region of *ATP2A1* (Figure S3G), thus indicating a potential activity of FOXO3 as a transcriptional regulator of both the *ATP2B1* and *ATP2A1* loci.

The phosphorylation of FOXO3 has been previously shown to be mainly triggered by PI3K/Akt pathway activation, which results in its exclusion from the nucleus and inhibition of the transcriptional activation of its target genes (Brunet *et al*., 1999; Manning and Cantley, 2007; Stefanetti et al., 2018). Thus, we further investigated the phosphorylation of Akt in the same cells infected with SARS-CoV-2. As expected, we confirmed the literature data showing increased levels of phosphorylated(S473)-Akt also in this *in-vitro* model (i.e., HEK293T-ACE2 cells) upon SARS-CoV-2 infection (Khezri et al., 2022) (Figure 3E).

Then, in order to validate the role of FOXO3 as a positive transcriptional regulator of *ATP2B1* and *ATP2A1*, we transiently overexpressed a FOXO3-encoding plasmid (containing the coding region of FOXO3; #14937, Addgene) in HEK293T-ACE2 cells. After 48 h from the start of transfection, immunoblotting data showed increased levels of both the FOXO3, ATP2B1 and ATP2A1 proteins (Figure 3F), thus suggesting that FOXO3 is a positive regulator of the *ATP2B1* and *ATP2A1* loci. These data confirm our hypothesis that FOXO3 is a good candidate to transcriptionally activate both membrane (i.e., ATP2B1) and endoplasmic reticulum (i.e., ATP2A1) Ca^2+^ pump expression.

Altogether, these data indicate that following SARS-CoV-2 infection, the activation of the PI3K/Akt pathway increases the levels of phosphorylated-FOXO3, thus causing its exclusion of FOXO3 from the nucleus, and, as a consequence, the inhibition of their target genes transcription, including the newly identified *ATP2B1* and *ATP2A1*. This mechanism leads to downregulation of ATP2B1 on plasma membranes and ATP2A1 on endoplasmic reticulum, thus increasing the intracellular Ca^2+^ levels, which enhances SARS-CoV-2 replication, as shown in the model presented in Figure 3G.

### Identification of an ATP2B1 targeting molecule that impairs SARS-CoV-2 infection and replication

To identify novel candidate compounds in the fight against SARS-CoV-2, we used artificial intelligence as a drug design computational tool to model the structure of ATP2B1 (PMCA1)– caloxin 2a1 (as a known ATP2B1 inhibitor; Pande et al., 2011) by docking and energy minimization modeling (Chaudhary et al., 2001) (Figure S4A). Thus, a pharmacophore model was built using the structures that we assumed for ATP2B1–exodom-2 and caloxin 2a1 (Figure S4A). Five pharmacophore features were produced. Among 230 million screened compounds, we selected 30 million by considering database filtering for solubility and absorption, 7,201 molecules by pharmacophore searches, and 1,028 molecules by docking scoring (Figure S4B). We then manually selected the top 22 molecules. Finally, two compounds (PI-7 and PI-8; Figure 4A, B) were identified and selected for further functional assays.

**Figure 4.**
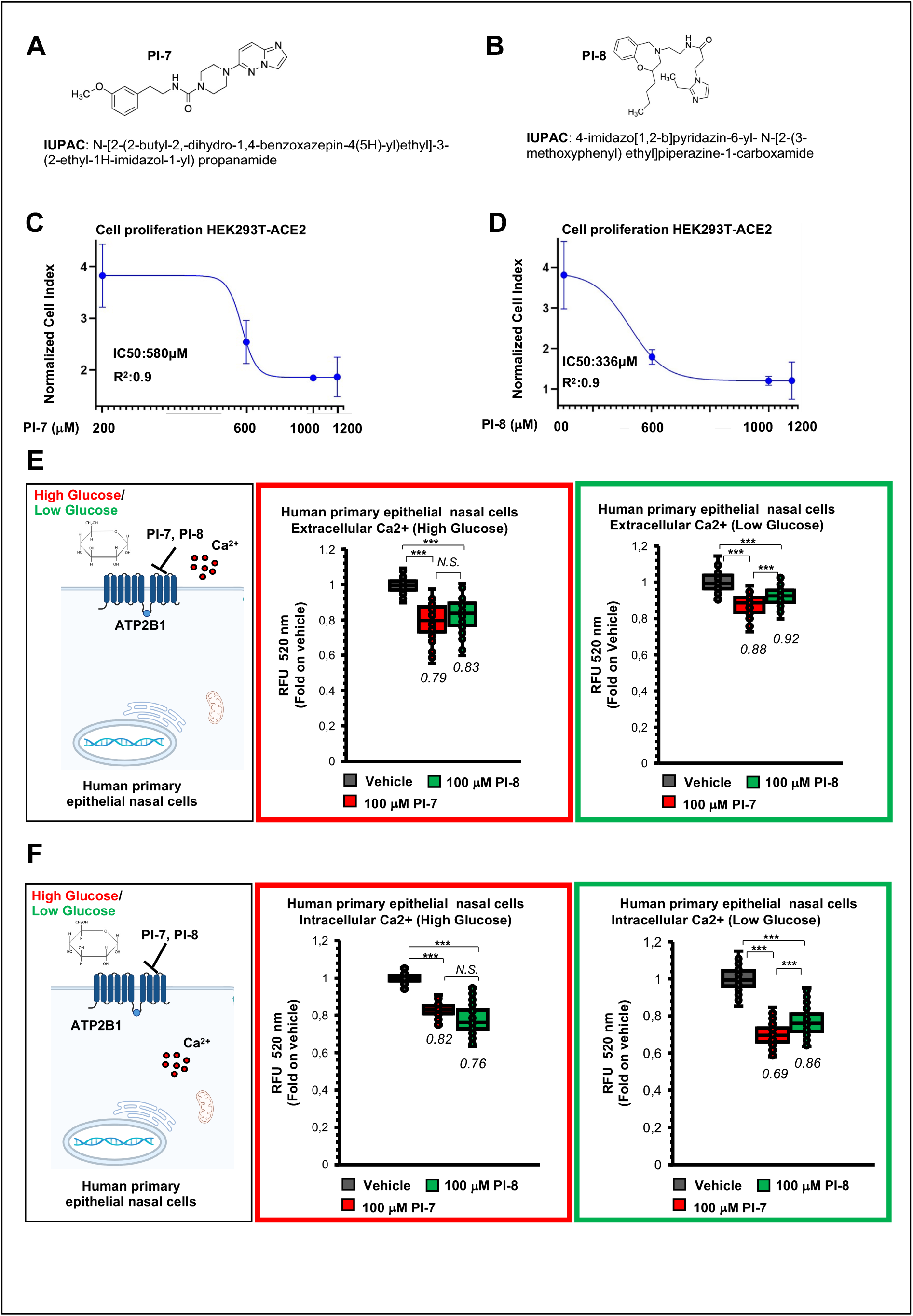
ATP2B1 impairment using the caloxin derivative (compounds PI-7 and PI-8) diminishes extra- and intra-cellular Ca^2+^ levels. **(A, B)** Molecular structures and IUPAC names of compounds PI-7 **(a)** and PI-8 **(b)** selected from the screening. **(C, D)** Real-time cell proliferation analyses in HEK293T-ACE2 cells (Cell Index; i.e., cell-sensor impedance expressed every 2 min). IC_50_ values calculated through nonlinear regression analysis performed with Graph Pad Prism 9 ([inhibitor] vs. response [three parameters]) for PI-7 **(C)** and PI-8 **(D)**. Briefly, HEK293T-ACE2 were plated and, after 2 h they were treated with the indicated concentrations of PI-7 and PI-8; with vehicle-treated cells (0.001% DMSO) as the negative control. Impedance was measured every 2 min over 48 h. The graphs were generated using Graph Pad Prism 9, with the IC50 values given (PI-7: 580 μM, R^2^ 0.9; PI-8: 336 μM, R^2^ 0.9). Data are means ±SD of three independent experiments. **(E)** Extracellular Ca^2+^ as measured using Fluo3-AM over 30 min in human primary epithelial nasal cells (1.2 ×10^4^ cells/well) plated in DMEM High Glucose or DMEM Low glucose and treated with 100 μM compounds PI-7 and PI-8, or with 0.001% DMSO as vehicle. After 24 hours from the treatment started, the cell media supernatant was used for the extracellular Ca^2+^ measurement. The graph shows quantification of relative fluorescence changes of Fluo3 as a measure of extracellular Ca^2+^ levels by showing the relative fluorescent units (RFUs) (excitation, 506 nm; emission, 526 nm) recorded every two minutes for 30 min by using a multimode plate reader (Enspire 2300, PerkinElmer). Data are means ±SD. *** p <0.001; N.S., not significant. (unpaired two-tailed student’s *t*-tests; N = 3 independent experiments per group). The fold values on vehicle controls are indicated within the graph. Red box: High Glucose; Green Box: Low glucose. Data are means ± SD of three independent experiments. **(F)** Intracellular Ca^2+^ as measured using Fluo3-AM over 90 min in human primary epithelial nasal cells (1.2 ×10^4^ cells/well) plated in DMEM High Glucose or DMEM Low glucose and treated with 100 μM compounds PI-7 and PI-8, or with 0.001% DMSO as the vehicle. After 24 hours from the treatment started, the cells were used for the intracellular Ca^2+^ measurement. The graph shows quantification of relative fluorescence changes of Fluo3 as a measure of intracellular Ca^2+^ levels by showing the relative fluorescent units (RFUs) (excitation, 506 nm; emission, 526 nm) recorded every two minutes for 90 min by using a multimode plate reader (Enspire 2300, PerkinElmer). Data are means ±SD. *** p <0.001; N.S., not significant. (unpaired two-tailed student’s *t*-tests; N = 3 independent experiments per group). The fold values on vehicle control are shown within the graph. Red box: High Glucose; Green Box: Low glucose. Data are means ±SD of three independent experiments.

We first assessed the cytotoxicity of these two compounds (PI-7 and PI-8) in terms of cell proliferation and apoptosis in HEK293T-ACE2 cells. The cell proliferation assay (based on the measurement of electrical impedance in real-time; i.e., cells index) was used to determine the half-maximal inhibitory concentrations (i.e., IC_50_) of PI-7 and PI-8. To this aim, we tested escalating doses (from 200 μM to 1200 μM) of PI-7 and PI-8 on HEK293T-ACE2 cells, and calculated the IC_50_ 48 h after the treatment started (IC_50_ values: PI-7, 580 μM, R^2^ 0.9; PI-8, 336 μM, R^2^ 0.9; Figure 4C-D and Figure S4C-D). Of interest, the lower doses of PI-7 and PI-8 (0.1-10 μM) did not alter the cell proliferation rates, as compared to vehicle-treated cells (vehicle: 0.001% dimethylsulfoxide [DMSO]) (Figure S4C-D). Similar results were obtained for apoptosis assays. Here, no activation of caspase 3 activity was shown in the HEK293T-ACE2 cells upon treatment with escalating doses of PI-7 and PI-8 (from 1 to 100 μM; Figure S5A). These data were further validated by immunoblotting analyses performed on the same treated cells with antibodies against the cleaved fragment of caspase 3 (i.e., activated caspase 3; Figure S5B, C). Altogether, these *in-vitro* data exclude anti-proliferative and pro-apoptotic actions of both PI-7 and PI-8 compounds.

Since an interplay between the Ca^2+^ homeostasis (regulated by ATPase 2B Ca^2+^ family pumps) and glucose metabolism has been since described (Dejos et al., 2020), we measured both the extracellular and intracellular Ca^2+^ level in human primary epithelial nasal cells in high (+) and low (-) glucose conditions. The data show that both extracellular and intracellular Ca^2+^ content are higher in those cells cultured in low (-) glucose conditions thus confirming literature data (Figure S5D-E on the left).

Thus, we evaluated the efficacy of both compounds PI-7 and PI-8 on *ATP2B1* pump inhibition in primary human epithelial nasal cells grown in media containing high (+) glucose and low (-) glucose content (see Figure 4E, F: red box, high (+) glucose; green box, low (-) glucose). The results indicate that both compounds (i.e., PI-7 and PI-8) reduced the extracellular Ca^2+^ content as compared to vehicle control (i.e., 0.001% DMSO) (Figure 4E, Figure S5D; high (+) glucose condition: PI-7, 0.79 fold lower [21% inhibition]; PI-8, 0.83 fold lower [17% inhibition]; low (-) glucose condition: PI-7, 0.88 fold lower [12 % inhibition]; PI-8, 0.92 fold lower [8% inhibition]). A similar trend was observed for the inhibition of the intracellular cytosolic Ca^2+^ content (Figure 4F, Figure S5E; high (+) glucose condition: PI-7, 0.82 fold lower [18% inhibition]; PI-8, 0.76 fold lower [24% inhibition]; low (-) glucose condition: PI-7, 0.69 fold lower [31% inhibition]; PI-8, 0.86 fold lower [14% inhibition]).

Taken altogether, the data show that compound PI-7 has a better performance in both extracellular and intracellular Ca^2+^ inhibition in low (-) glucose condition. This latest condition is responsible for increased intracellular and extracellular Ca^2+^ levels (Figure S5D-E on the left) that are both required for SARS-CoV-2 infection and replication.

For the above reasons, we focused on compound PI-7 for the following experiments. To dissect the intracellular alterations due to treatment with PI-7, we performed a proteomic analysis in the HEK293T-ACE2 cells upon treatment with 1 μM PI-7 for 24 h (Figure S6A). In the treated cells, there were 17 downregulated and 66 upregulated proteins (Table S5). We then generated a network of protein interactions through the “Search Tool for Retrieval of Interacting Genes/Proteins” (STRING) database, with both the upregulated and downregulated (Figure S6A) proteins. This showed that the upregulated proteins take part in the common networks mostly involved in the regulation of metabolic processes and gene expression (Figure S6A, Table S6). Of importance, among the down-regulated proteins, we found some that are involved in viral transcription and viral processes (Figure S6A in bold, Table S7).

To dissect out the potential antiviral activity of compound PI-7, we treated HEK193T-ACE2 cells with 1 μM PI-7 and then infected them with SARS-CoV-2 (VOC Delta) at 0.026 MOI for 72 h (Figure 5A). Immunoblotting data show that PI-7 decreased the viral N protein levels, thus demonstrating inhibition of viral replication (Figure 5B). Of note, the data also showed increased ATP2B1 and ATP2A1 protein levels in PI-7-treated and infected cells, as compared to vehicle control (Figure 5B). The overexpression of ATP2B1 was found to decrease the intracellular Ca^2+^ levels (Figure 2B) and the treatment with compound PI-7 was shown to diminish both the extracellular and intracellular Ca^2+^ levels (Figure 4E-F). Our data are here suggesting that compound PI-7 exerts antiviral activity by decreasing the extracellular and intracellular Ca^2+^ levels, also by potentially promoting its uptake into the endoplasmic reticulum) mostly due to inhibition of ATP2B1 activity and upregulation of ATP2A1.

**Figure 5.**
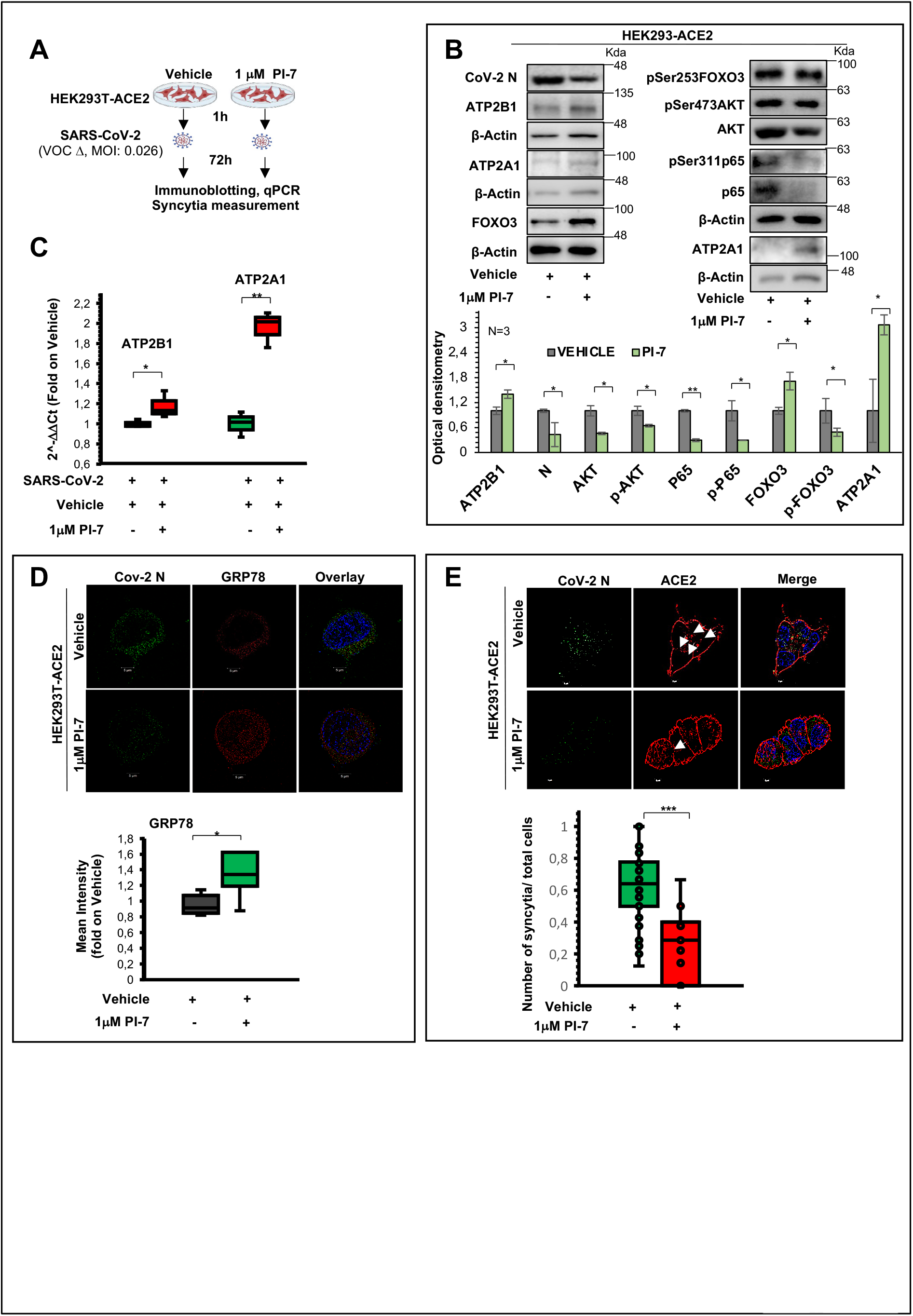
ATP2B1 impairment using the caloxin derivative (compound PI-7) diminishes SARS-CoV-2 propagation and syncytia formation. **(A)** Experimental plan for HEK293T-ACE2 cells treated with 1 μM PI-7, or 0.001% DMSO as vehicle control. After 1 h, the cells were infected with SARS-CoV-2 viral particles of VOC Delta at 0.026 MOI for 72 h. The cells were then fixed or lysed for immunofluorescence or qPCR and immunoblotting, respectively. **(B)** Representative immunoblotting analysis using antibodies against the indicated proteins on total protein lysates obtained from HEK293T-ACE2 treated as in (**b**). All experiments were performed in triplicate. Densitometry analysis of the indicated band intensities in blots from three independent experiments. Data are means ±SD. * p <0.05, ** p<0.01; (unpaired two-tailed student’s *t*-tests; N = 3 independent experiments per group). **(C)** Quantification of mRNA abundance relative to that in control (CTR) cells (2^−ΔΔCt^) for the human *ATP2B1* and *ATP2A1s* genes, from RT-PCR analysis of RNA extracted from HEK293T-ACE2 cells treated as in (**A**). Data are means ±SD. * p <0.05, ** p <0.01 (unpaired two-tailed student’s *t*-test; N = 3 independent experiments per group). **(D)** Immunofluorescence staining with an antibody against viral CoV-2 N (green) and human GRP78 (red) proteins in HEK293T-ACE2 cells treated with PI-7 molecules and infected with SARS-CoV-2 for 72 hours (i.e., treated as in **A**). The graph showing the intensity of fluorescence was shown on the left. Data are means ± SD. P-value was determined by unpaired two-tailed Student’s *t* test; N = three independent experiments per group. The SIM^2^ image was acquired with Elyra 7 and processed with Zeiss ZEN software (blue edition). Magnification, ×63. Scale bar, 5 μm. **(E)** Representative immunofluorescence staining with an antibody against the CoV-2 viral N protein (green) and human ACE2 (red) in HEK293T-ACE2 cells treated as in (**A**). The SIM^2^ image was acquired with Elyra 7 and processed with Zeiss ZEN software (blue edition). Magnification, ×63. Quantification of the relative proportions of syncytia in >300 cells per condition. Data are means ±SD, with the P-value determined by unpaired two-tailed Student’s *t*-test (N = three independent experiments per group). Scale bar, 5 μm.

Then, because of the positive transcriptional regulation of *ATP2B1* and *ATP2A1* mediated by FOXO3 (Figure 3F), we also verified its protein levels upon treatment with compound PI-7 in the same infected cells. These data show an increased level of total unphosphorylated (transcriptionally active) FOXO3 protein amount, and decreased levels of inactive phosphorylated (S253)-FOXO3 (Figure 5B). Thus, the enhanced transcriptional activity of unphosphorylated FOXO3, due to PI-7 treatment, would also explain why *ATP2B1* protein amount is found upregulated in the treated cells (Figure 5B).

Furthermore, because the phosphorylation of FOXO3, and its exclusion from nucleus, had been previously shown to be triggered by PI3K/Akt pathway activation (Brunet *et al*., 1999; Manning and Cantley, 2007; Stefanetti *et al*., 2018) also during SARS-CoV-2 infection (Figure 3E), we further investigated the phosphorylation status of Akt in those cells infected with SARS-CoV-2. As expected, our data confirmed a decrease amount of phosphorylated (S473)-AKT upon compound PI-7 treatment (Figure 5B), thus further confirming the antiviral action of this molecule against SARS-CoV-2 infection.

Altogether, compounds PI-7 inhibits AKT signaling that in turn lowers the phosphorylation of FOXO3 (on Serine 253), thus resulting in increased levels of transcriptionally active (unphosphorylated) FOXO3 that modulates the expression of its target at nuclear levels, including the transcriptional activation of *ATP2B1* and *ATP2A1* (Figure 5B). Dissecting further how AKT/FOXO3 phosphorylation mechanism of action is occurring during SARS-CoV-2 infection will be an issue of future studies.

Of interest, FoxO transcriptional factors have already been shown to have a role in immune cell maturation and inflammatory cytokines secretion (Cheema *et al*., 2021). Among the FoxOs, FoxO3 has already been shown to modulate innate immune responses to infections of the airway epithelium through modulation of secretion of several cytokines from immune cells (Xin et al., 2018). This occurs through inhibition of the NF-κB inflammatory pathway, the activation of which is exploited by the SARS-CoV-2 (Thompson *et al*., 2015). Thus, restoring the transcriptional activity of FoxO3 might relieve the inflammatory burst following SARS-CoV-2 infection.

To test this hypothesis, we investigated NF-κB inflammatory pathway through immunoblotting in the same PI-7-treated and infected cells. Our data show decreased levels of phosphorylated (Ser311)-p65 in PI-7-treated cells, as compared to the vehicle control (Figure 5B). Furthermore, in order to exclude that NF-κB inhibition is only a consequence of a reduced viral infection in these SARS-CoV-2-infected cells previously treated with compound PI-7, we have also tested the phosphorylation of p65 in non-infected human primary nasal cells treated with compound PI-7 for 24 hours (Figure S6B). Our data show decreased levels of phosphorylated (Ser311)-p65 in PI-7-treated cells in absence of SARS-CoV-2 (Figure S6B), thus showing the efficacy of compound PI-7 to impair NF-κB inflammatory pathway. To further confirm, the anti-inflammatory action of this molecule in HEK-293T-ACE2 cells upon SARS-CoV-2 infection, we measured the expression levels of some of the main cytokines targeted by NF-κB that take part in the COVID19 cytokine storm (Hu et al., 2021; Rabaan et al., 2021) (i.e., IL-1β, IL-6, and TNF-α). Our qPCR data showed a statistically significant reduction in the levels of these cytokines in PI-7-treated cells, as compared to the vehicle controls (Figure S6C).

Altogether, these results show the ability of compound PI-7 to (i) reduce phosphorylated (S473)-AKT and phosphorylated (S253)-FOXO3 and (ii) increase unphosphorylated active FOXO3 protein levels as a result of reduced viral replication in HEK293T-ACE2 cells upon infection by SARS-CoV-2 (VOC Delta, 72 h) (Figure 5B). As a consequence, this leads to impairment of the NF-κB inflammatory pathway (also mediated by activation of FOXO3) and inhibition of cytokine expression upon treatment with compound PI-7 (Figure 5B, Figure S6C). Overall this data show an anti-inflammatory property of compound PI-7 by reducing phosphorylated (S473)-AKT and phosphorylated (S253)-FOXO3 phosphorylations. When this action is taking part in infected SARS-CoV-2 cells will be issue of future investigations.

Of interest, the treatment with compound PI-7 not only restored *ATP2B1 and ATP2A1* protein amounts, but also increased their transcritpional levels, mostly as a consequence of FOXO3 transcriptional activity (Figure 5C).

As Ca^2+^ transition had been previously reported to be correlated to the endoplasmic reticulum stress(Deniaud et al., 2008), we validated our qPCR data on ATP2A1 levels through the levels of endoplasmic reticulum stress. To this aim, high-resolution immunofluorescence analyses were performed using the lattice SIM^2^ technology (ELYRA7, ZEISS), which showed increased levels of GRP78, as a marker of endoplasmic reticulum stress, in the SARS-CoV-2– infected HEK293T-ACE2 cells upon treatment with compound PI-7, in comparison with vehicle-treated cells (Figure 5D). These results suggested that treatment with PI-7 decreases intracellular Ca^2+^ levels also by restoring the expression of *ATP2A1* (whose levels were decreased in the presence of SARS-CoV-2, Figure 1D) that is involved in the translocation of Ca^2+^ from the cytosol to the sarcoplasmic reticulum lumen (Minton, 2014).

Taking all together, compound PI-7 is able to impair *ATP2B1* pump activity in terms of extracellular and intracellular Ca^2+^ decrease during SARS-CoV-2 infection, potentially because of the PI3K/AKT pathway inhibition, reduction of phosphorylation of FOXO3 and enhanced expression of *ATP2B1* (that is responsible for decreased levels of intracellular Ca2+, see Figure 2B) and *ATP2A1* (that promotes Ca^2+^ uptake into the endoplasmic reticulum, thus causing reticulum stress, see Figure 5D). A more comprehensive action at the transcriptional level of compound PI-7 will be further detailed in the near future.

As many enveloped viruses (including SARS-CoV-2) have been shown to cause fusion of the neighboring cells into multinucleated ‘syncytia’ (Braga et al., 2021), we further investigated here the antiviral activity of compound PI-7 on inhibition of syncytia generation. Thus, to determine the relative proportions of syncytia, the same HEK293T-ACE2 cells were treated with PI-7 (1 μM) or vehicle (0.001% DMSO), infected with SARS-CoV-2, and fixed for immunofluorescence analyses using an antibody against the ACE2 protein (Figure 5E). We found decreased syncytia percentages in the cells upon treatment with PI-7 (P=7.9E-5) (Figure 5E). As a further control, lower expression levels of the “syncytia marker” TMEM16 (Braga *et al*., 2021) were found in PI-7-treated HEK293T-ACE2 cells upon SARS-CoV-2 infection, (fold: 0.6, see Figure S6D).

Altogether, these data indicate the anti-viral activity of compound PI-7 against SARS-CoV-2 infection in these HEK293T-ACE2 cells, with decreased levels of the viral N protein and lowered levels of syncytia formation.

### The anti-inflammatory actions of compound PI-7 via FOXO3–NF-κB block the COVID19 cytokine storm against Omicron 2 variant

Finally, the antiviral activity of compound PI-7 was validated in human primary epithelial nasal cells infected with the latest SARS-CoV-2 variant (VOC: Omicron 2) at 0.04 MOI for 70 hours of infection (Figure 6A). Our qPCR analyses supported the antiviral activity of PI-7 also against the Omicron 2 SARS-CoV-2 variant, with a reduction in the viral N protein levels, the expression of which was increased in these infected cells (Figure 6B). Furthermore, the data show that PI-7 can restore the expression levels of the calcium pumps (i.e., ATP2B1, ATP2A1) and can also reduce the NF-κB–induced cytokines (i.e., IL-1β, IL-6, TNF-α) in these cells infected with the Omicron 2 SARS-CoV-2 variant (Figure 6B).

**Figure 6.**
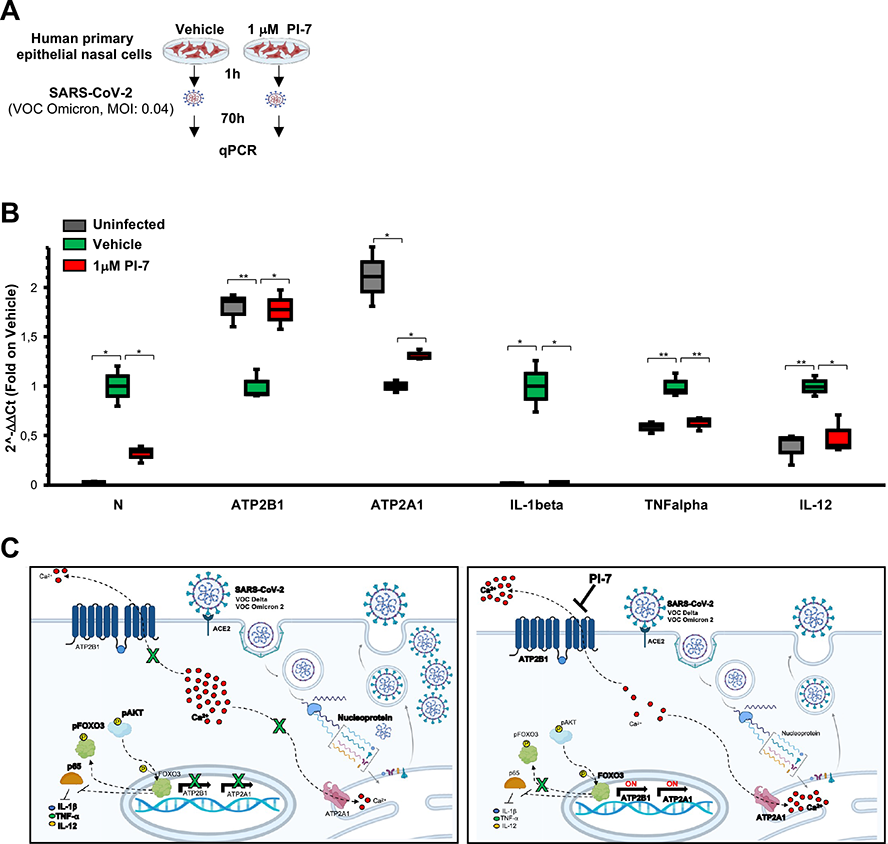
Antiviral activity of caloxin-derivative (compound PI-7) is also shown via inhibition of the cytokine storm against Omicron 2 variant. **(A)** Experimental plan for human primary epithelial cells treated with 1 μM PI-7 or 0.001% DMSO as vehicle control. After 1 h, the cells were infected with SARS-CoV-2 viral particles of VOC Omicron 2 at 0.04 MOI for 70 h. The cells were then used for qPCR analysis. **(B)** Quantification of mRNA abundance relative to that in control (Vehicle) cells (2^−ΔΔCt^) for the indicated cytokines in HEK293T-ACE2 cells treated as in (**A**). Data are means ±SD. \**P* <0.05, ** P <0.01 (as determined by unpaired two-tailed Student’s *t*-tests; n = three independent experiments per group). Uninfected cells are used as negative control. **(C)** Cartoon representation to illustrate our hypothesis for the role of *ATP2B1* during SARS-CoV-2 infection upon treatment with compound PI-7. **On the left:** during SARS-CoV-2 infection, the PI3K/Akt pathway is activated via Akt phosphorylation, thus enhancing the phosphorylation of FOXO3 to exclude it from the nucleus (Manning and Cantley, 2007, Stefanetti et al., 2018). As a consequence, the expression of the FOXO3 targets, including ATP2B1 and ATP2A1, are reduced, thus increasing the intracellular Ca^2+^ levels and further promoting SARS-CoV-2 replication. Furthermore, SARS-CoV-2 infection also leads to NF-κB activation (also due to reduction of active FOXO3; Thompson et al., 2015) and, as a consequence, the expression of the inflammatory cytokines (e.g., IL-1β, TNF-α and IL-12) belonging to the cytokine storm resulted increased. **On the right:** Treatment with the caloxin-derivative (compound PI-7) inhibits the viral propagation of SARS-CoV-2 (VOC: Delta and Omicron 2) by blocking ATP2B1 pump activity, thus reducing both the extra- and intracellular Ca^2+^ levels that are necessary for SARS-CoV-2 replication. Compound PI-7 also increases *ATP2B1* and *ATP2A1* levels, thus further reducing the intracellular Ca^2+^ levels because of its extracellular export via *ATP2B1* (on the plasma membrane), and potentially the endoplasmic reticulum import through *ATP2A1*. These increased levels of *ATP2B1* and *ATP2A1* occurred because of the enhancement of transcriptionally active FOXO3. The levels of active unphosphorylated-FOXO3 are increased due to the inactivation of PI3K/AKT pathway (due to decreased viral infection upon compound PI-7). As a consequence, the levels of nuclear active FOXO3 results increased, thus enhancing the transcriptional activation of *ATP2B1* and *ATP2A1.* Treatment with PI-7 also reduces NF-κB activation (due to enhancement of FOXO3) and as a consequence, the expression of the inflammatory cytokines belonging to the cytokine storm resulted decreased IL-1β, TNF-α and IL-12. All the gene names whose expression resulted upregulated are depicted in bold.

Overall, these data show the efficacy of compound PI-7 against SARS-CoV-2 VOC Omicron 2 via targeting inflammatory pathways in coordination of Ca^2+^ signaling.

## Discussion

Ca^2+^ as an important second messenger in excitable and non-excitable cells, where it controls essential functions including cellular signaling processes and immune responses(Brini et al., 2013). Intracellular and organellar Ca^2+^ concentrations are tightly controlled via various pumps, ATPases, ion channels, and uniporters. Among these pumps, PMCAs and SERCAs are considered an efficient line of defense against abnormal Ca^2+^ rises. During viral infection, cellular Ca^2+^ dynamics are highly affected, as dysregulation of the host cell signaling cascades is elicited by these infectious agents (Berlansky *et al*., 2022).

Two mechanisms of regulation of Ca^2+^ can be envisioned upon SARS-Cov-2 infection: one related to the extracellular virus-host interaction, and the other linked to an intracellular mode of action once the virus has entered the cell. The presence of extracellular Ca^2+^ can positively influence virus-host interactions for the correct binding of CoVs to the ACE2 receptor and during membrane fusion when the virus is entering the cell(Shang et al., 2020). Intracellular Ca^2+^ has been reported to enhance SARS-CoV-2 replication by activating downstream processes, such as alteration of the host cellular metabolism and acceleration of inflammation (Serebrovska et al., 2020).

Here, we focused on understanding how intracellular Ca^2+^ modulation can influence SARS-CoV-2 infection and replication. Gene expression data obtained using HEK293T-ACE2 cells infected with SARS-CoV-2 showed downregulation of Ca^2+^ signaling pathways, mostly mediated by decreased levels of the PMCA and SERCA Ca^2+^ pumps. This thus suggests increased levels of intracellular Ca^2+^ defined by indirect, but positive, evidence (gene expression data), as almost all of the Ca^2+^ pumps were downregulated (Figure 1D in blue and Table S1). Then in the additional validation studies we focused on ATP2B1, showing a time-dependent decrease in expression in response to SARS-CoV-2 infection. Furthermore, an opposite correlation was seen between viral replication (as shown by the viral N protein levels; and ATP2B1 levels of expression in the two different cellular models (i.e., HEK293T-ACE2 cells [see Figure 1F, H] and human primary epithelial nasal cells [Figure 2E]). At this time we envision future studies to investigate the responses to other Ca^2+^ pumps on the endoplasmic reticulum, mitochondria, and Golgi complex following SARS-CoV-2 infection, to dissect out further its mechanism of action.

Of interest, using a new caloxin-derivative molecule (i.e., compound PI-7), we show decreased intracellular Ca^2+^ levels (mimicking ATP2B1 overexpression in human primary epithelial nasal cells) and also extracellular Ca^2+^ content. Whether the addition of PI-7 results in binding and blocking of ATP2B1 pump to decrease the exports of Ca^2+^ into the extracellular environment will be the aim of future deep investigations, potentially through structural conformational studies and protein–drug crystallography. Here we hypothesized a secondary mechanism of control of expression of ATP2B1 and other Ca^2+^ pumps to maintain protein expression by juxtaposing compound PI-7 action and by controlling the intracellular and extracellular Ca^2+^ signaling.

Of importance, prophylactic treatment of HEK293T-ACE2 cells before SARS-CoV-2 infection (VOCs Delta and Omicron 2) exerts antiviral actions on viral replication (as measured by decreased viral N protein levels). The results showing decreased syncytia formation in the PI-7-treated cells, compared to vehicle controls, provide further support here (see Figure 5E). This is a result of impairment of the generation of syncytia activation by the SARS-CoV-2 Spike protein at the level of the cell plasma membrane. Furthermore, Ca^2+^ is known to be of importance for syncytia generation; indeed, drugs that inhibit TMEM16 activity (a Ca^2+^-activated ion channel; e.g., niclosamide) blunted Ca^2+^ oscillations in Spike-expressing cells, and as a consequence, inhibited Spike-driven syncytia formation (Braga *et al*., 2021). The expression of TMEM16 in our assays further confirmed these results (see Figure 5E and S6D). Of importance, our data also show restoration of ATP2A1 (ATPase sarcoplasmic/endoplasmic reticulum Ca^2+^ transporting 1; SERCA1) (see Figure 5B, C). Results supporting this hypothesis are also presented in a model upon compound PI-7 treatment of SARS-Cov-2 infection (Figure 6C).

Overall, our data indicate that upon SARS-CoV-2 infection there is downregulation of Ca^2+^ pumps on the cell membrane (ATP2B1) and the endoplasmic reticulum (ATP2A1) due to a further mechanism of action responsible for increased intracellular Ca^2+^ levels during viral infection and replication. How and when Ca^2+^ can pass between the endoplasmic reticulum, cytoplasm, and plasma membrane during SARS-CoV-2 replication are questions to address in the near future.

One of the mechanisms responsible for reduction of expression of ATP2B1 and ATP2A1 during SARS-CoV-2 infection involves the FOXO3 transcription factor and its functional regulation. Our study also demonstrates positive regulation of the *ATP2B1* and *ATP2A1* locus by FOXO3. Of interest, during SARS-CoV-2 infection, we observed increased levels of inactive phosphorylated-(Ser253)-FOXO3 and a substantial reduction in its total protein content under these specific cellular conditions (see Figure 3E). The phosphorylation of FOXO3 has been previously shown to be triggered by the PI3K/Akt pathway, thus causing its exclusion from the nucleus and inhibiting the transcriptional activation of its target genes (Brunet *et al*., 1999; Manning and Cantley, 2007; Stefanetti *et al*., 2018), including the newly identified target here (ATP2B1 and ATP2A1). Of note, over-activation of PI3K/Akt pathway during SARS-CoV-2 infection has also been reported (Khezri *et al*., 2022), and is here confirmed in our *in-vitro* model.

Thus, we hypothesized that one of the mechanisms responsible for the increased FOXO3 phosphorylation that inactivates the transcriptional regulation function of FOXO3 will accordingly decrease ATP2B1 and ATP2A1 levels during SARS-CoV-2 infection. Of interest, activated Akt is also required for intracellular Ca^2+^ release during other viral infections (e.g., Herpes simplex virus; Cheshenko et al., 2013). Thus, taken together, activation of ATP2B1 and ATP2A1 by FOXO3 is a mechanism of escape from virus replication of infected cells, with ATP2B1 being both responsible for the pumping of Ca^2+^ from the cytoplasm (and consequently out of the cells) together with the action of ATP2A1, which stores the cytoplasmic Ca^2+^ in the endoplasmic reticulum. Together, these two controlled mechanisms of action will impair viral replication. Future studies will further investigate the link between PI3K/Akt and the intracellular Ca^2+^ release during SARS-Cov-2 infection (see model presented in Figure 6C).

Of importance, in a cohort of infected symptomatic patients (n. 197 patients affected by severe COVID19 and 370 asymptomatic cases), we identified a rare homozygous intron variant in the *ATP2B1* locus as a novel genetic factor responsible for severe COVID19 predisposition (Table 1). It remains to be identified how the nucleotide region that contains the polymorphism (i.e., rs111337717) can act as a modulator (enhancer or inhibitor). According to our model, we postulate the presence of this SNP variant would influence negatively on the regulation of the ATP2B1 locus and its expression. Future experiments need to be demonstrated to address this hypothesis.

To date, variants in the *ATP2A1* genes that are responsible for COVID19 predisposition have not been reported. The data presented here underline the marker identification to stratify those people who retain the C/C variant in ATP2B1 (rs111337717), as they might be subjected to severe COVID19 following virus infection and replication (see Table 1).

As summarized in (Figure S4 and S5), compound PI-7 was tested here at nontoxic concentrations (i.e., 1 μM), in terms of cell proliferation and apoptosis. It has the potential to reduce the intracellular Ca^2+^ levels (Figure 4E-F) that are necessary for SARS-CoV-2 replication and propagation, as measured by the reduced levels of the viral N protein (see Figure 5B) (the activity of which is essential for genome/ subgenome transcription and maturation of newly formed virions) and reduced numbers of syncytia (Figure 5E).

Finally, the literature data show negative cross-talk between FOXO3 and the NF-κB inflammatory pathway (Thompson *et al*., 2015), which allowed us to visualize the anti-inflammatory actions of PI-7 in SARS-CoV-2–infected human cells. Treatment with compound PI-7 increased FOXO3 protein levels, and definitively reduced the activation of NF-κB, thus further diminishing the levels of inflammatory cytokines with a known role in the cytokine storm in COVID19 patients (see Figure 6C and Figure S6C). We thus envision compound PI-7 in preventive nd therapeutic manner, and with this anti-inflammatory action could prevent the cytokine storm during SARS-CoV-2 infection. At this time, it remains to investigate whether additional NPRL3 inflammasome signaling is further affected (Lee et al., 2012; Nieto-Torres et al., 2015). However, how an increase in cytosolic Ca^2+^ promotes NLRP3 inflammasome activation is not yet clear. It has been proposed that an increase in cytosolic Ca^2+^ causes Ca^2+^ overloading of the mitochondria, resulting in mitochondrial dysfunction, and thus leading to NLRP3 inflammasome activation. Moreover, the Golgi complex has a key role as an intracellular Ca^2+^ store, in synergy with the endoplasmic reticulum. Regulation of the NLPR3 inflammasome by compound PI-7 is today unclear or uninvestigated and will be an issue of future studies.

Thus, considering the ubiquitous action of compound PI-7 on the mechanism of regulation of the virus machinery that influences both virus replication and propagation during the early status of infection, we envision its use in preventive therapeutic strategies to fight the COVID19 pandemia. As compound PI-7 affects Ca^2+^ cellular homeostasis and the expression of both membrane ATP2B1 and endoplasmic reticulum specific ATP2A1 Ca^2+^-dependent cellular pumps, and most importantly, as it has anti-inflammation properties with downregulation of the pro-inflammatory function of NF-κB, we envision here that infection by any emerging SARS-CoV-2 variants (e.g., VOC Delta and Omicron 2) will also be inhibited. Indeed, both SARS-Cov-2 Delta and Omicron 2 showed inhibition here and impairment of the most related cytokines of the early storm (see Figure 6B and S6C). Finally, knowing the impaired PI3K/Akt signaling of phosphorylation by compound PI-7, we predict here its application in cancer, in those tumors where such enhanced signalings is evident and pronounced. It is not a surprise that an anti-COVID19 drug can provide benefits in the fight against cancer, and we foresee its use in fragile or cancer-affected patients with poor innate and adaptive immunity (Hosseini et al., 2020; Schultze and Aschenbrenner, 2021; Sette and Crotty, 2021). In light of the determining effect of Ca^2+^ signaling in viral infection and replication and in the immune response, we used the alternative approach of molecular docking to screen for compounds that target the host machinery hijacked by the virus during infection. This approach represents a promising therapeutic strategy that instead of looking for the development of antiviral therapeutic molecules that target viral proteins, looks directly at the host Ca^2+^ channels, resulting in broad applicability.

However, progress to bring such drugs to the clinic faces important challenges. There are three main reasons for hesitation. First, Ca^2+^ signaling might affect several biological pathways. Indeed, side effects might be observed in patients treated with these types of drugs. Secondly, genetic loss-of-function studies in mice and in cells suggest incorrect regulation of the inflammatory responses caused by global Ca^2+^ misregulation. Thirdly, and more critically, animal studies with these small molecules to sustain ATP2B1 expression have to confirm the absence of risk of overt adverse effects. At this time, the mechanism of action and compound PI-7 represent additional weapons to impair further waves of infections by COVID19, as new variants are now expected.

## ETHICAL COMMITTEE APPROVAL

The Ethical Committee approvals for the COVID19 samples use in this study were as follows: (i) protocol no. 141/20; date: 10 April 2020, CEINGE TaskForce Covid19; Azienda Ospedaliera Universitaria Federico II, Direzione Sanitaria, protocol no. 000576 of 10 April 2020; (ii) protocol no. 157/20; date: 22 April 2020, GENECOVID, with the experimental procedures for the use of SAR-CoV-2 in a biosafety level 3 (BSL3) laboratory were authorized by Ministero della Sanità and Dipartimento Di Medicina Molecolare e Biotecnologie Mediche, Università degli Studi di Napoli Federico II and Azienda Ospedaliera Universitaria Federico II, Direzione Sanitaria protocol no. 0007133 of 08 May 2020; (iii) protocol no. 18/20; date: 10 June 2020, Genetics CEINGE TaskForce Covid19; Azienda Ospedaliera Universitaria Federico II, Direzione Sanitaria protocol no. 000576 of 10 April 2020.

## STAR-METHODS

### Cell culture

HEK-293T cells (CRL-3216, ATCC) and HEK-293T stable clones overexpressing human ACE2 (HEK293T-ACE2 cells) were grown in a humidified 37 °C incubator with 5% CO_2_. The cells were cultured under feeder-free conditions using Dulbecco’s modified Eagle’s medium (DMEM; 41966-029; Gibco) with 10% fetal bovine serum (10270-106; Gibco), 2 mM L-glutamine (25030-024; Gibco), and 1% penicillin/streptomycin (P0781; Sigma-Aldrich), with the medium changed daily.

Freshly isolated human nasal epithelial cells were collected by nasal brushing of healthy donors (as previously described; Ferrucci *et al*., 2021). These primary human epithelial cells (EVA-EMBL-EBI; project ID: PRJEB42411; analyses: ERZ1700617) were cultured in PneumaCult (no. 05009; STEMCELL Technologies) with 2 mM L-glutamine (25030-024, Gibco), and 1% penicillin/streptomycin (P0781, Sigma-Aldrich). The cells were dissociated with Trypsin-EDTA solution (T4049, Sigma-Aldrich) when the cultures reached ∼80% confluency.

### Generation of HEK293T-ACE2 stable clones

HEK293T-ACE2 stable clones were generated as previously described (Ferrucci *et al*., 2022). Briefly, HEK293T cells (at ∼70% confluency) were transfected with 1 μg DNA plasmid pCEP4-myc-ACE2 (#141185, Addgene) using X-tremeGENE 9 DNA Transfection Reagent (06365779001; Sigma-Aldrich) diluted with serum-free DMEM (41966-029; Gibco), to a concentration of 3 μL reagent/100 μL medium (3:1 ratio [μL]). The transfection complex was added to the cells after 15 min of incubation, in a dropwise manner. At 48 h from transfection, the cell culture medium was changed, and the cell clones were selected using 800 mg/mL hygromycin.

### Transient transfections

#### ATP2B1-overexpressing cells

Primary human nasal epithelial cells (5 ×10^5^) were seeded into 6-well culture plates, and after 24 h they were transiently transfected with the plasmid DNA construct pMM2-hATP2B1b (#47758, Addgene). Transient transfections were performed with X-tremeGENE 360 Transfection Reagent (XTG360-RO, #08724105001, Roche), according to the manufacturer instructions. Briefly, XTG360-RO DNA Transfection Reagent was diluted with serum-free DMEM (41966-029; Gibco) (10 μL reagent/500 μL medium). Then, 5 μg DNA plasmid was added to 500 μL diluted X-tremeGENE 9 DNA Transfection Reagent. The transfection reagent:RNA complex was incubated for 15 min at room temperature. The transfection complex was then added to the cells in a dropwise manner. Twelve hours after transfection, the cell culture medium was changed. At 48 h after transfection started, the cells were used for Ca^2+^ assays and immunoblotting analyses.

#### FOXO3-overexpressing cells

HEK-293T-ACE2 cells (3×105) were seeded in a 6 well plate and transfected for 48 hours with Flag-FOXO3 (Addgene Flag-FOXO3 #153142) plasmid. Transient transfections were performed with X-tremeGENE™ 360 Transfection Reagent (XTG360-RO, #08724105001, Roche), according to the manufacturer’s instructions. Briefly, XTG360-RO DNA Transfection Reagent diluted with serum-free Dulbecco’s modified Eagle’s medium (41966-029; Gibco) (10 μl reagent/500 μl medium). Then, 2 μg per well of DNA plasmid were added to 250 μl of diluted X-tremeGENE 360 DNA Transfection Reagent. The transfection reagent:DNA complex was incubated for 15 minutes at room temperature. The transfection complex was then added to the cells in a dropwise manner. Twelve hours after transfection, the cell culture medium was changed. After 48 hours from the transfection started, the cells were used for immunoblotting analyses.

### RNA interference

#### Primary human nasal epithelial cells

Primary human nasal epithelial cells (5 ×10^5^) were seeded into 6-well culture plates, and after 12 h they were transiently transfected with the ATP2B1 siRNA (sc-42596, Santa-Cruz). A pool of a three siRNAs (sc-37007, sc-44230, sc-44231, Santa-Cruz) was used as the negative control. RNA interference via siRNAs was performed using X-tremeGENE 360 Transfection Reagent (XTG360-RO), according to the manufacturer instructions. Briefly, X-tremeGENE 360:DNA (1μg/μL) (3:1) were mixed in DMEM without fetal bovine serum, and incubated at room temperature for 20 min. The mixture was then added to the cells in a dropwise manner and incubated for 48 h and 72 h for intracellular Ca^2+^ assays (as described below).

#### HEK293T-ACE2 cells

HEK-293T-ACE2 cells (3 ×10^5^) were seeded into 6-well culture plates, and after 24 h they were transiently transfected with the ATP2B1 siRNA (sc-42596, Santa-Cruz). A pool of three siRNAs (sc-37007, sc-44230, sc-44231, Santa-Cruz) was used as the negative control. Transient transfections were performed with Lipofectamine RNAimax (13778-150, Invitrogen), according to the manufacturer instructions. Briefly, Lipofectamine RNAimax was diluted with serum-free DMEM (41966-029; Gibco) (9 μL reagent/150 μL medium). The siRNAs were then diluited in serum-free DMEM (3 μL siRNA/150 μL medium) to obtain the 30 pmol concentration. The transfection reagent:RNA complex was then incubated for 5 min at room temperature. The transfection complex was then added to the cells in a dropwise manner. After 12 h and 24 h the cells were infected with SARS-CoV-2 viral particles or lysed for immunoblotting analyses.

### *In-vitro* treatment with compound PI-7

HEK293T-ACE and primary human nasal epithelial cells (2.5 ×10^5^) were plated in 6-well plates and treated with 1 μM compound PI-7, or with 0.001% DMSO as the vehicle control. The proteomic analyses were performed after 24 h of treatment. The viral infections were performed after 1 h of treatment.

### *In-vitro* treatment with Valproic Acid (VPA)

HEK293T-ACE2 cells were plated (2 ×10^5^ cells/well) into 6-well plates. They were them treated with 20 mM Valproic Acid (VPA, #P4543, Sigma-Aldrich) or 0.01% DMSO (as negative control) for 16 h (as previously described; Zhang et al., 2013). Later the cells were lysed to investigate whether FOXO related genes are regulated.

### Cell proliferation assays (i.e., cell index)

Real-time cell proliferation analysis for the Cell Index (i.e., the cell-sensor impedance was expressed every two minutes as a unit called “Cell Index”). HEK-293T-ACE2 cells (1.5 ×10^4^) were plated. After 2 hours cells were treated with the indicated concentrations of PI-7 and PI-8 (from 200 to 1200 uM or from 0,1 to 10 uM); with vehicle-treated cells were the negative control. Impedance was measured every 2 min over 48 hours. The IC50 values were calculated through nonlinear regression analysis performed with Graph Pad Prism 9 ([inhibitor] vs. response (three parameters). Data are means ± SD of three independent experiments.

### Ca^2+^ assays

#### Extra- and intra-cellular Ca^2+^ assays in human primary epithelial nasal cells treated with compound PI-7

Human nasal primary epithelial cells were plated (1.2 ×10^4^ cells/well) into 24-well plates previously coated with PureCol (1:30,000; #5005, Advanced BioMatrix) with DMEM High Glucose (4500 mg/L, 41966-029; Gibco) or DMEM Low Glucose (1 g/L, D6046, Sigma-Aldrich). The cells were then treated with 100 μM compound PI-7 or vehicle (i.e., 0.001% DMSO) as negative control. After 24 hours from the treatment started, the cell media supernatant was used for the extracellular Ca^2+^ measurement, while the cell lysates for the intracellular Ca^2+^ relevation.

Regarding to the extracellular Ca^2+^, the cell media supernatant (100 μl) was transfer into 96-well plates (3917, Costar) and incubated for 30 min at room temperature, and further 30 min at 37°C, with a solution (1:1, v:v; i.e., 100 μl) of MEM without phenol red and fetal bovine serum containing 2.5 µM Flu-3-AM (F1241, Invitrogen) supplemented with Pluronic F-127 (1:1, v:v; P3000MP, Invitrogen) and 2.5 mM probenecid inhibitor (57-66-9, Invitrogen). The relative fluorescent units (RFUs) were acquired (excitation, 506 nm; emission, 526 nm) and recorded every two minutes (for 30 min) by using a multimode plate reader (Enspire 2300, PerkinElmer). Regarding the intracellular Ca^2+^ measurement, the treated human nasal primary epithelial cells were washed two times with minimum essential medium (MEM; 21090-022, Gibco) without phenol red and fetal bovine serum. They were then incubated for two hours at room temperature with a solution (300 μl) of MEM without phenol red and fetal bovine serum containing 2.5 µM Flu-3-AM (F1241, Invitrogen) supplemented with Pluronic F-127 (1:1, v:v; P3000MP, Invitrogen) and 2.5 mM probenecid inhibitor (57-66-9, Invitrogen). The cells were subsequently washed three times with MEM containing probenecid. The cells were then further incubated for 1 hour at room temperature with 300 μl of 2 mM probenecid inhibitor dissolved in MEM, to allow hydrolysis of the AM ester bond, and then rinsed with MEM containing probenecid. The cells were then mechanically lysed and 100 μl were transferred into 96-well plates (3917, Costar) for intracellular (cytoplasmic) Ca^2+^ measurment. The relative fluorescent units (RFUs) were acquired (excitation, 506 nm; emission, 526 nm) every two minutes (up to 90 min) using a multimode plate reader (Enspire 2300, PerkinElmer).

#### Intracellular Ca2+ assays in human primary epithelial nasal cells overexpressing-or downregulating-ATP2B1

Nontransfected human nasal primary epithelial cells and those transfected with the ATP2B1 siRNA (for 48 h) were plated (3 ×10^4^ cells/well) into 96-well plates (3917, Costar) previously coated with PureCol (1:30,000; #5005, Advanced BioMatrix). The cells were washed two times with minimum essential medium (MEM; 21090-022, Gibco) without phenol red and fetal bovine serum. They were then incubated for 30 min at room temperature with a solution of MEM without phenol red and fetal bovine serum containing 2.5 µM Flu-3-AM (F1241, Invitrogen) supplemented with Pluronic F-127 (1:1, v:v; P3000MP, Invitrogen) and 2 mM probenecid inhibitor (57-66-9, Invitrogen). The cells were subsequently washed three times to remove any dye that was nonspecifically associated with the cell surface, with MEM containing probenecid. The cells were then further incubated for 20 min at room temperature with 2 mM probenecid inhibitor dissolved in MEM, to allow hydrolysis of the AM ester bond, and then rinsed with MEM containing probenecid. The cells were then treated with 10 mM CaCl_2_ (C3306-500G, Sigma) in MEM containing probenecid. The relative fluorescent units (RFUs) were immediately acquired (excitation, 506 nm; emission, 526 nm) using a multimode plate reader (Enspire 2300, PerkinElmer).

### Caspase assay

HEK-293T-ACE2 (3×10^4^) were plated in 96-well plate and treated with escalating doses (1 uM, 10 uM, 100 uM) of the drug PI-7 or the drug PI-8. Vehicle-treated (0.001% DMSO) cells were used as negative controls. Staurosporine-treated cells (10 μM) were used as positive controls of the assays. The assay was performed by using Caspase-3 Activity Assay Kit (#5723, Cell Signaling), by following the manufacture’s instruction. Briefly, after 18 hours from the treatment started, cell medium was removed and 30 ul of cell lysis buffer (PI-7018, Cell Signaling) were added. The plate was then incubated for 5 minutes on ice. Later, 25 ul of cell lysate were mixed with 200 ul substrate solution B (#11734S, Cell Signaling) in a black 96 well plate. Fluorescence was detected by using a multimode plate reader (Enspire, 2300, PerkinElmer). The positive control AMC (25 μl, #11735S, Cell Signaling) mixed with 200 ul 1X assay buffer A (#11736S, Cell Signaling) was used as positive control of fluorescence.

### SARS-CoV-2 isolation

SARS-CoV-2 was isolated from a nasopharyngeal swab obtained from an Italian patient, as previously described (Ferrucci *et al*., 2021). Briefly, Vero E6 cells (8 ×10^5^) were plated in DMEM (41966-029; Gibco) with 2% fetal bovine serum in a T25 flask, to which the clinical specimen was added. The inoculated cultures were grown in a humidified 37 °C incubator with 5% CO_2_. When cytopathic effects were observed (7 days after infection), the cell monolayers were scraped with the back of a pipette tip, and the cell culture supernatant containing the viral particles was aliquoted and frozen at −80 °C. Viral lysates were used for total nucleic acid extraction for confirmatory testing and sequencing (GISAID accession numbers: VOC Delta, EPI_ISL_3770696; VOC Omicron2, EPI_ISL_10743523).

### SARS-CoV-2 infection

HEK293T-ACE2 and human primary epithelial nasal cells treated with 1 μM compound **PI-7** or vehicle (0.001% DMSO) were infected with SARS-CoV-2 viral particles. Noninfected cells were used as the negative control of infection. After 24, 48 and 72 h of infection, the cells were lysed for qPCR, immunoblotting, and RNA-seq, or fixed for immunofluorescence analyses. These experiments were performed in a BLS3-authorized laboratory.

### RNA extraction and qPCR assays

RNA samples were extracted using TRIzol RNA Isolation Reagent (15596026, Invitrogen), according to the manufacturer instructions. Reverse transcription was performed with 5× All-In-One RT MasterMix (g592; ABM), according to the manufacturer instructions. The reverse transcription products (cDNA) were amplified by qRT-PCR using an RT-PCR system (7900; Applied Biosystems, Foster City, CA, USA). The cDNA preparation was through the cycling method, by incubating the complete reaction mix as follows: cDNA reactions: (37 °C for 10 min and 60 °C for 15 min); Heat-inactivation: 95 °C for 3 min; Hold stage: 4 °C.The targets were detected with the SYBR green approach, using BrightGreen BlasTaq 2× PCR MasterMix (G895; ABM). Human *ACTB* was used as the housekeeping gene to normalize the quantification cycle (Cq) values of the other genes. These runs were performed on a PCR machine (Quantstudio5, Lifetechnologies) with the following thermal protocol: Hold stage: 50 °C for 2 min; Denaturation Step: 95 °C for 10 min; Denaturation and annealing (×45 cycles): 95 °C for 15 s and 60 °C for 60 s; Melt curve stage: 95 °C for 15 s, 60 °C for 1 min, and 95 °C for 15 s. Relative expression of the target genes was determined using the 2^−ΔΔCq^ method, as the fold increase compared with the controls. The data are presented as means ±SD of the 2^−ΔΔCq^ values (normalized to human ACTB) of three replicates.

### Primers sequences for qPCR analyses

The details of the primers used in these SYBR green assays are provided in the supplementary information.

CoV-2 N1 Forward: GACCCCAAAATCAGCGAAAT

CoV-2 N1 Reverse: TCTGGTTACTGCCAGTTGAATCTG

ACTB Forward: GACCCAGATCATGTTTGAGACCTT

ACTB Reverse: CCAGAGGCGTACAGGGATAGC

IL-1β Forward: ATGATGGCTTATTACAGTGGCAA

IL-1β Reverse: GTCGGAGATTCGTAGCTGGA

TNFA Forward: CTCTTCTGCCTGCTGCACTTTG

TNFA Reverse: ATGGGCTACAGGCTTGTCACTC

ATP2B1 Forward: CGATCTCACTGGCTTATTCAGTCA

ATP2B1 Reverse: TAGCTGTAGCATTTCCCATGGTT

ATP2A1 Forward: AAC GAG GCC AAA GGT GTC TA

ATP2A1 Reverse: TCG AGA GGC TTC TCA CAT C

IL-12 Forward: TGA TGG CCC TGT GCC TTA GT

IL-12 Reverse: GGA TCC ATC AGA AGC TTT GCAT

### Immunoblotting

Cells were lysed in 20 mM sodium phosphate, pH 7.4, 150 mM NaCl, 10% (v/v) glycerol, 1% (w/v) sodium deoxycholate, 1% (v/v) Triton X-100, supplemented with protease inhibitors (Roche). The cell lysates were cleared by centrifugation at 16,200× *g* for 30 min at room temperature, and the supernatants were removed and assayed for protein concentrations with Protein Assay Dye Reagent (BioRad). The cell lysates (20 μg) were resolved on 10% SDS-PAGE gels. The proteins were transferred to PVDF membranes (Millipore). After 1 h in blocking solution with 5% (w/v) dry milk fat in Tris-buffered saline containing 0.02% [v/v] Tween-20, the PVDF membranes were incubated with the primary antibody overnight at 4 °C: anti-SARS-CoV-2 N protein (1:250; 35-579; ProSci Inc.); anti-PMCA ATPase (1:500; 5F10; # MA3-914, Invitrogen); anti-β-actin (1:10000; A5441; Sigma); anti-p65 (1:1000; sc-372, Santa Cruz Biotechnology); anti–p-(Ser^311^)p65–NF-κB (1:500; sc-101748. Santa Cruz Biotechnology); anti-Caspase-3 (1:500, ab49822, Abcam); anti-phosphorylated-Ser473-AKT (1:250, ab81283, Abcam); anti-AKT1 (1:500, #2967, Cell Signaling); anti-p-(Ser^253^)FOXO3A (1:500, ab47285, Abcam); anti-FOXO3A (1:1000, ab47409, Abcam). The membranes were then incubated with the required secondary antibodies for 1 h at room temperature, as secondary mouse or rabbit horseradish-peroxidase-conjugated antibodies (NC 15 27606; ImmunoReagents, Inc.), diluted in 5% (w/v) milk in TBS-Tween. The protein bands were visualized by chemiluminescence detection (Pierce-Thermo Fisher Scientific Inc., IL, USA). Densitometry analysis was performed with the ImageJ software. The peak areas of the bands were measured on the densitometry plots, and the relative proportions (%) were calculated. Then, the density areas of the peaks were normalized with those of the loading controls, and the ratios for the corresponding controls are presented as fold-changes. Immunoblotting was performed in triplicate. Densitometry analyses shown were derived from three independent experiments. Membrane and cytoplasmic protein fractions of the cultured cells were obtained with Mem-PER Plus Membrane Protein Extraction kits (89842, Thermo Scientific).

### High resolution immunofluorescence

SARS-CoV-2–infected HEK293T-ACE2 cells were fixed in 4% paraformaldehyde in phosphate-buffered saline (PBS) for 30 min, washed three times with PBS, and permeabilized for 15 min with 0.1% Triton X-100 (215680010; Acros Organics) diluted in PBS. The cells were then blocked with 3% bovine serum albumin (A9418; Sigma) in PBS for 1 h at room temperature. The samples were incubated with the appropriate primary antibodies overnight at 4 °C: anti-ACE2 (1:1000; ab15348; Abcam); anti-SARS N protein (1:100; 35-579; ProSci Inc); or anti-GRP78 BiP (1:1000, ab21685, Abcam). After washing with PBS, the samples were incubated with the secondary antibody at room temperature for 1 h, as anti-mouse Alexa Fluor 488 (1:200; ab150113; Abcam) or anti-rabbit Alexa Fluor 647 (1:200; ab150075; Abcam). DNA was stained with DAPI (1:1000; #62254, Thermo Fisher). The slides were washed and mounted with cover slips with 50% glycerol (G5150; Sigma-Aldrich). Microscopy images were obtained using the Elyra 7 platform (Zeiss) with the optical Lattice SIM^2^ technology (with the ZEN software, Zeiss, blue edition), using the 63× oil immersion objective.

### Differential proteomics analysis

Differential proteomic analysis was carried out using a shotgun approach. In detail, three biological replicates of HEK293T-ACE2 cells treated for 24 h with compound PI-7 or with the vehicle control (0.001% DMSO) were lysed with lysis buffer (50 mM ammonium bicarbonate, 5% sodium dodecyl sulfate), and the protein extracts were quantified by BCA assay. The equivalent of 50 µg of each protein extract was reduced, alkylated, and digested onto S-Trap filters according to the Protifi protocol (Protifi, Huntington, NY), as previously reported. Peptide mixtures were dried in a SpeedVac system (Thermo Fisher, Waltham, MA) and an aliquot for each replicate was subjected to a clean-up procedure using a C18 zip-tip system (Merck KGaA, Darmstadt, Germany). Desalted peptide mixtures were resuspended in a solution of 0.2% HCOOH in LC-MS grade water (Waters, Milford, MA, USA) and 1 μL of each was analyzed using an LTQ Orbitrap XL (Thermo Scientific, Waltham, MA) coupled to the nanoACQUITY UPLC system (Waters). Samples were initially concentrated onto a C18 capillary reverse-phase pre-column (20 mm, 180 μm, 5 μm) and then fractionated onto a C18 capillary reverse-phase analytical column (250 mm, 75 μm, 1.8 μm), working at a flow rate of 300 nL/min. Eluents “B” (0.2% formic acid in 95% acetonitrile) and “A” (0.2% formic acid, 2% acetonitrile, in LC-MS/MS grade water, Merck) were used with a linear two-step gradient. The first step went from 5% B to 35% B in 150 min, and the second step from 35% B to 50% B in 10 min. MS/MS analyses were performed using Data-Dependent Acquisition (DDA) mode, after one full scan (mass range from 400 to 1800 *m/z*), with the 10 most abundant ions selected for the MS/MS scan events, and applying a dynamic exclusion window of 40 s. All of the samples were run in technical duplicates. The raw data were analyzed with MaxQuant 1.5.2 integrated with the Andromeda search engine. For the MaxQuant, the following parameters were used: a minimum of four peptides including at least two unique for protein identification; a false discovery rate (FDR) of 0.01 was used. Protein quantification was performed according to the label free quantification intensities, using at least four unmodified peptides razor + unique.

### Protein-protein interaction networks

To investigate the interactions between the protein products of the top differentially expressed proteins in the HEK293T-ACE2 cells following treatment with compound PI-7, a protein interaction network was generated using the Search Tool for the Retrieval of Interacting Genes/ Proteins (STRING) database (https://string-db.org). The nodes consisted of genes and the edges were derived from experimentally validated protein-protein interactions.

### RNA sequencing (RNA-seq)

#### RNA isolation and library construction and sequencing

Total RNA was isolated from the HEK293T-ACE2 cells using TRIzol RNA Isolation Reagent (#15596018; Ambion, Thermo Fisher Scientific). It was then quantified in a NanoDrop One/OneC Microvolume UV-Vis spectrophotometer (Thermo Scientific), checked for purity and integrity, and submitted to Macrogen Europe B.V. for sequencing. Libraries were prepared using TruSeq Stranded mRNA Library Prep kits according to the protocols recommended by the manufacturer (i.e., TruSeq Stranded mRNA Reference Guide # 1000000040498 v00). Trimmed reads were mapped to the reference genome with HISAT2 (https://ccb.jhu.edu/software/hisat2/index.shtml), a splice-aware aligner. After the read mapping, Stringtie (https://ccb.jhu.edu/software/stringtie/) was used for transcript assembly. The expression profile was calculated for each sample and transcript/gene as read counts, FPKM (fragment per kilobase of transcript per million mapped reads) and TPM (transcripts per kilobase million).

#### Analysis of differentially expressed genes

Differentially expressed genes analysis was performed on a comparison pair (SARS-CoV-2–infected *vs* noninfected cells). The read count value of the known genes obtained through the -e option of the StringTie was used as the original raw data. During data preprocessing, low quality transcripts were filtered out. Afterwards, Trimmed Mean of M-values (TMM) normalization was performed. Statistical analysis was performed using Fold Change, with exactTest using edgeR per comparison pair. For significant lists, hierarchical clustering analysis (Euclidean method, complete linkage) was performed to group the similar samples and genes. These results are depicted graphically using a heatmap and dendogram. For the enrichment test, which is based on Gene Ontology (http://geneontology.org/), DB was carried out with a significant gene list using g: Profiler tool (https://biit.cs.ut.ee/gprofiler/). Pathway enrichment analysis was performed using the KEGG database (http://www.genome.jp/kegg/).

### ChIP-Seq analysis

ChIP-Seq of FOXO3 and the input control were downloaded from E-MTAB-2701.XC1. Reads were quality checked and filtered with Trimmomatic(Bolger et al., 2014). Alignments to the reference genome (hg19) were performed with BWA aln(Li and Durbin, 2010), using the default parameters. SAMtools rmdup (Li et al., 2009) was used to remove potential PCR duplicates. SAMtools sort and index were used to sort and index the bam files. Uniquely mapped reads of the FOXO3 signal were normalized over genomic input log_2_[FOXO3/Input]) using the bamCompare tool from Deeptools suite(Diaz et al., 2012) with the exactScaling method as the scaling factor. The normalized FOXO3 signal was visualized with UCSC genome browser.

### Phenotype definition

The COVID19 asymptomatic cohort were selected as previously described [PMID:33815819]. The cohort included individuals from Campania (Italy) screened in May 2020 for SARS-CoV-2 who were positive for SARS-CoV-2 antibodies but without any COVID19 symptoms in the three previous months, such as hospitalization requirement, fever, cough or at least two symptoms among sore throat, headache, diarrhea, vomiting, asthenia, muscle pain, joint pain, smell or taste loss, and shortness of breath (Lavezzo et al., 2020). The COVID19 hospitalized cohort was selected as previously described (Russo et al., 2021;,Andolfo et al., 2021).

#### In-silico analysis

The list of 351 coding variants of ATP2B1 was obtained from gnomAD (https://gnomad.broadinstitute.org/). FATHMM scores were used to assess the pathogenicity of the coding variants(Shihab et al., 2013). The GTEx database(Consortium, 2020)was used to select the eQTLs for ATP2B1, setting the level of significance at P<1 ×10^-6^. The GWAVA tool was used to assign a functional score to each SNP(Ritchie et al., 2014). RegulomeDB was used to evaluate the effects of SNPs on altering TFBS(Boyle et al., 2012).

#### DNA extraction, SNP genotyping

Genomic DNA of cases was extracted from peripheral blood using Maxwell RSC Blood DNA kits (Promega, Madison, WI, USA), and DNA concentrations and purities were evaluated using a NanoDrop 8000 spectrophotometer. The DNA samples were genotyped by the TaqMan SNP Genotyping Assay for the SNPs rs111337717 and rs116858620 (Applied Biosystems by Thermo Fisher Scientific, Waltham, MA, USA).

### In-silico analysis of single-cell RNA-sequencing data

Publicly available single-cell RNA-sequencing data for adult human lung from COVID19 patients were used, from https://singlecell.broadinstitute.org/single_cell/study/SCP1219/columbia-university-nyp-covid-19-lung-atlas (Melms et *al.*, 2021). The proportions of ATP2B1-, ATP2B2-, ATP2B3-, and ATP2B4-positive cells in different lung cell types were calculated. In a similar manner, the expression of ATP2B1, ATP2B2, ATP2B3, ATP2B4, ATP2A1, ATP2A2, ATP2A3, and FOXOs in these lungs from COVID19 patients and controls were investigated.

### Statistical analyses

Statistical significance was defined as P <0.05 by unpaired two-tailed Student’s *t*-tests. All of the data are given as means ±SD. In the Figures, statistical significance is represented as follows: *P <0.05, **P <0.01, and ***P <0.001.

All of the data from the qRT-PCR assays were analyzed using un-paired two-tailed Student’s *t-*tests, by comparing: (i) noninfected *versus* vehicle control infected cells (i.e., cells infected and treated with the vehicle as a control for the PI-7 treatment); and (ii) PI-7-treated cells *versus* vehicle control infected cells. Caspase-3 activities were analyzed using unpaired two-tailed Student’s *t-*tests, by comparing the RFU values measured after 1 h of incubation to those obtained at the experimental starting point (i.e., time 0). The IC_50_ values for PI-7 and PI-8 were calculated by nonlinear regression analysis, performed with Graph-Pad Prism 9 (using [inhibitor] *versus* response [three parameters]). All of the experiments were performed in triplicate. Allele and genotype frequencies were compared using chi-square or Armitage tests. A two-sided p <0.05 was considered statistically significant. Proteomic data were statistically analyzed using the Perseus software (Perseus version 1.6.15.0). Briefly, a Student’s *t*-tests were applied with a false discovery rate cut-off of 0.05 and with a log_2_FC cut-off of 0.5.

## RESOURCE AVAILABILITY

### Lead contact

Further information and requests for resources and reagents should be directed to and will be fulfilled by the lead contact, Prof. Massimo Zollo.

#### Materials availability

This study did not generate new unique reagents.

#### Data and code availability

- Gene expression (RNAseq) data are deposited on RNA data bank at EBI (https://www.ebi.ac.uk), July 11- 2022 (code: E-MTAB-11973).
- Any additional information required to reanalyze the data reported in this paper is available from the lead contact upon request.

## Data Availability

Further information and requests for resources and reagents should be directed to and will be fulfilled by the lead contact, Prof. Massimo Zollo.
This study did not generate new unique reagents.
Gene expression (RNAseq) data are deposited on RNA data bank at EBI (https://www.ebi.ac.uk), July 11. 2022 (code: E-MTAB-11973).
Any additional information required to reanalyze the data reported in this paper is available from the lead contact upon request.

## Acknowledgements

We thank P. Forestieri (President, CEINGE) and M. Giustino (CEO, CEINGE) for collaborative support of the program within the Regione Campania Covid19 Taskforce. We thank Christofer Berrie for providing professional and critical editing of the manuscript. We thank Prof. Francesco Broccolo for his valuable discussion on the therapeutic uses of the drug in a future therapeutic settings.

## Funding

This study was supported by the project “CEINGE TaskForce COVID19,” code D64I200003800 by Regione Campania for the fight against COVID19 (DGR no. 140; 17 March 2020). We further thank for support the Italian Association for Cancer Research (AIRC) Grant IG no. 22129 (M.Z.), the European School of Molecular Medicine for a doctorate program Fellowship (F.A.) and the Molecular Medicine Doctorate program Fellowship (F.B.), Fondazione Celeghin Italiana (M.Z.), and Ministero dell’Università e della Ricerca Italiana (PRIN) grant no. 2017FNZRN3 (M.Z.). We also thank NRF 2018R1A5A2025079–Korean Ministry of Science and ICT (J.-H.C.).

## Author contributions

P.D.A. generated the HEK293T-ACE2 stable cell clones; V.F. performed immunofluorescence analyses and syncytia measurements. F.B. and F.A. performed immunoblotting analyses. R.S. prepared RNA and performed qPCR analyses; C.S. prepared proteins from cell lysates. F.A. performed intracellular Ca^2+^ assays and caspase analyses. F.G. and S.A. performed Chip-Seq analyses. A.B. and G.P. performed bioinformatic analyses. G.F., S.B., M.V., B.M.P., and P.C. performed SARS-CoV-2 infections in a BSL3-authorized laboratory; R.R., V.A.L., S.C., M.C., and A.I. performed polymorphisms analyses. J.H.C. and H.Y.K. performed docking analyses. M.Z. designed the experiments, discussed the experimental plans with the authors, and wrote the manuscript. All of the authors discussed the results and commented on the manuscript.

## Conflicts of interest

The authors declare that they have no conflicts of interest.

## Supplemental Informations

**Figure S1.**
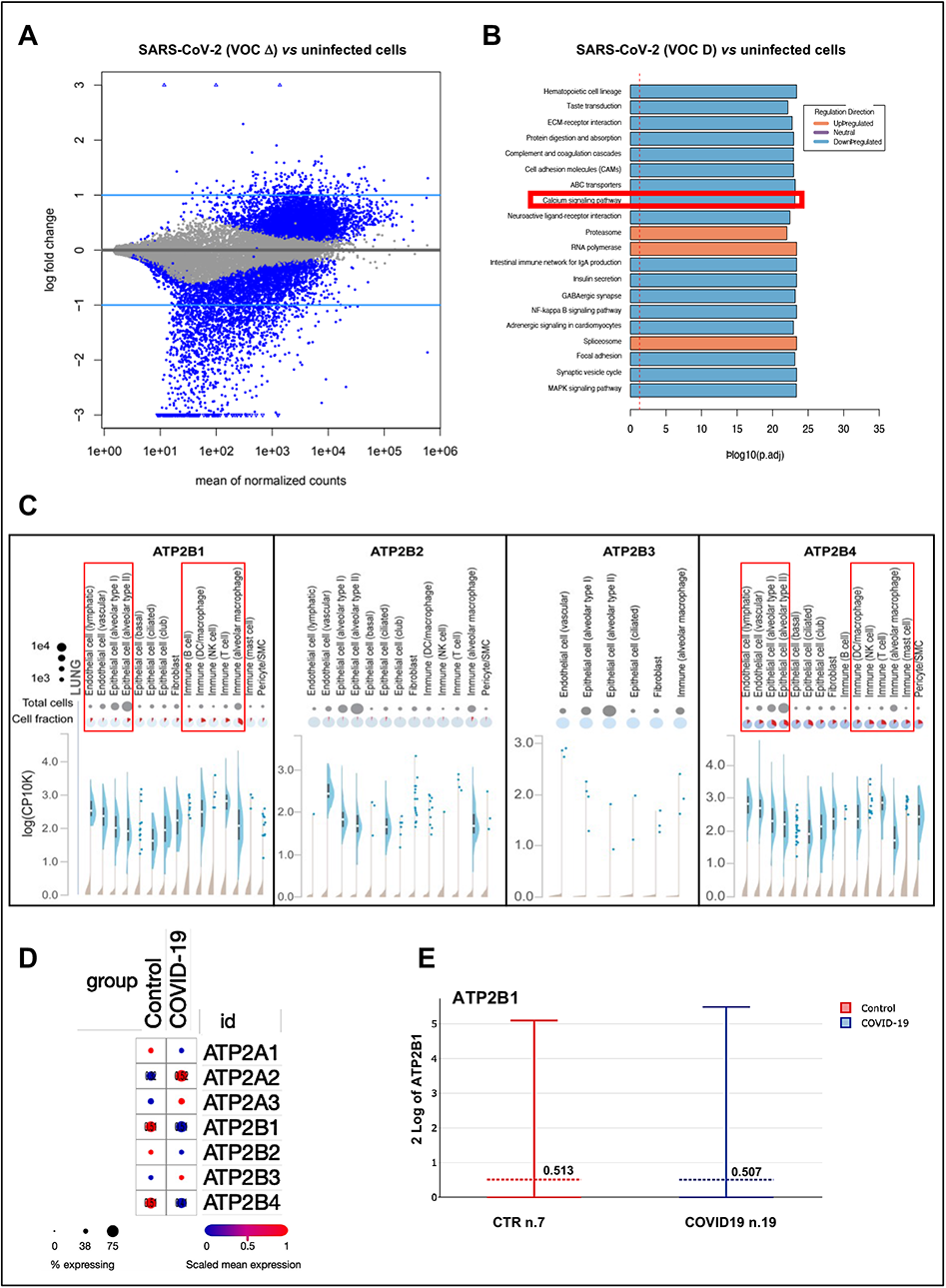
Ca2+ pumps, including ATP2B1, expression are deregulated during SARS-CoV-2 infection. Related to Figure 1. **(A)** RNA Sequencing (RNA-seq) analyses was performed in HEK293T-ACE2 cells infected by SARS-CoV-2 (VOC Delta, 0.026 MOI for 24 hours). The mean of normalized cound here shown to get an overview over similarities and dissimilarities between SARS-CoV-2-infected non infected cells. The analysis of the transcriptional changes in gene transcripts identified 234 up-regulated and 1742 down-regulated genes in SARS-CoV-2-infected cells compared to not-infected cells as differentially expressed (i.e., DEG). **(B)** The Gene Set Enrichment Analysis (GSEA) was then applied for the identification of deregulated key genes and pathways. In total, 234 up-regulated and 1742 down-regulated genes in SARS-CoV-2-infected cells compared to not-infected cells were mapped to KEGG pathways (P <0.05). The sorted terms by P-value (P < 0.05) are shown. KEGG pathway enrichment analysis indicate those significant deregulated genes were highly clustered in calcium signaling pathway (red box). P adj: adjusted P-values. **(C)** In silico analysis of publicly available data sets of single-cell RNA sequencing (https://singlecell.broadinstitute.org) for the expression of the plasma membrane calcium ATPases members of the large family of type Calcium ion pumps (PMCAs or ATP2B1-4) in multiple cell type in the lung parenchyma (including alveolar macrophages and in the alveolar epithelial cells type I and type II). **(D)** Literature public search on available datasets obtained from a single-nuclei RNA-seq (snRNA-seq) on >116,000 nuclei from n.19 COVID-19 autopsy lungs and n.7 pre-pandemic controls (Melms et al., 2021); https://singlecell.broadinstitute.org/single_cell/study/SCP1052/covid-19-lung-autopsy-samples) to verify if PMCAs and SERCAs pumps (ATP2B1-4 and ATP2A1-3 genes, respectively) showed distinct fractional and dysfunctional changes across the lungs from COVID-19 decedents. **(e)** Expression levels of ATP2B1 in single-nuclei RNA-seq (snRNA-seq) analyses performed on >116,000 nuclei from n.19 COVID-19 autopsy lungs and n.7 pre-pandemic controls (Melms *et al*., 2021); https://singlecell.broadinstitute.org/single_cell/study/SCP1052/covid-19-lung-autopsy-samples).

**Figure S2.**
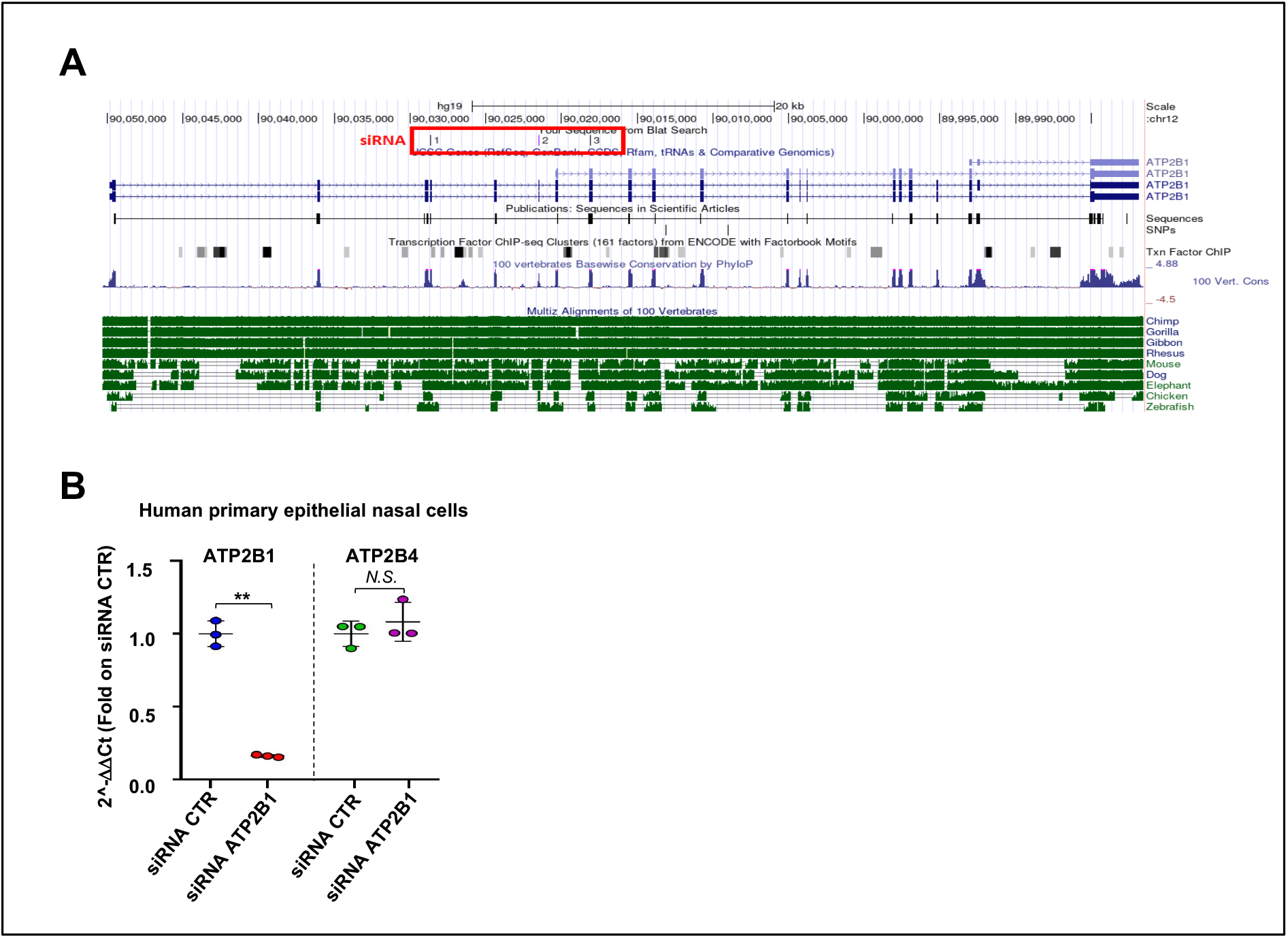
ATP2B1 reduced protein amount promotes SARS-CoV-2 replication. Related to Figure 2. **(A)** Representation of human ATP2B1 region recognized by siRNA as reported in the UCSC Genome Browser on Human Dec. 2013 (GRCh38/hg38) Assembly (https://genome.ucsc.edu/). At the bottom, the alignment of this genomic region among different species is shown. **(B)** Quantification of mRNA abundance relative to that in control (CTR) cells (2^−ΔΔCt^) for ATP2B1 and ATP2B4. RT-PCR analysis of RNA extracted from human primary epithelial nasal cells downregulating PMCA (after 48 hours from transfection with the ATP2B1 siRNA). Cells overexpressing a pool of three siRNA control were used as negative control of the experiment. Data are means ± SD. ** p < 0.01,; NS, not statistic (unpaired two-tailed student’s t test; N = 3 independent experiments per group

**Figure S3.**
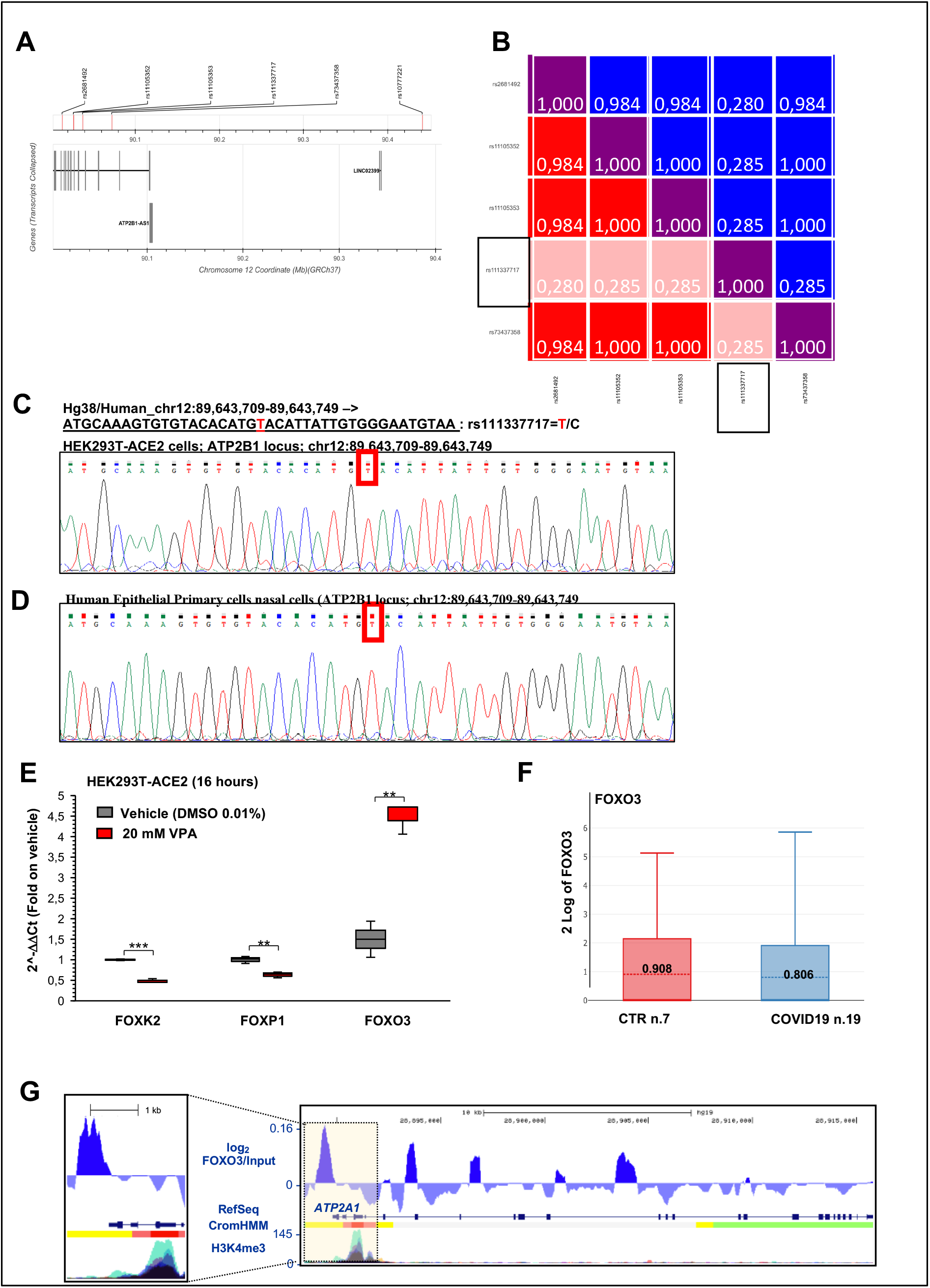
The homozygous intronic variant rs11337717 in ATP2B1 locus is responsible for SARS-CoV-2 increased replication in COVID19 patients. Related to Figure 3. **(A, B)** Linkage disequilibrium (LD) analyses on the top 5 SNPs (rs11105352; rs11105353; rs73437358; rs111337717; rs2681492) in order to select those independent. The SNP rs10777221 was excluded from this analyses because located at most 5’ region in extragenic ATP2B1 locus region (**A**). The graph in (**B**) shows the only SNP not in LD is rs111337717 (black boxes). **(C)** Sanger sequencing of the genomic region of ATP2B1 locus (chr12:89,643,709-89,643,749) in HEK293T-ACE2 cells to exclude the presence of intronc variance potentially responsible for alterated transcriptional levels of ATP2B1 gene. The red box indicate the nucleotide wild type allele for the SNP here studied. **(D)** Sanger sequencing of the genomic region of ATP2B1 locus (chr12:89,643,709-89,643,749) in human primary epithelial nasal cells to exclude the presence of intronic variants potentially responsible for alterated transcriptional levels of ATP2B1 gene. **(E)** Quantification of mRNA abundance relative to that in control (CTR) cells (2−ΔΔCt) for FOXK2, FOXP1 and FOXO3 genes. RT-PCR analysis of RNA extracted from HEK293T-ACE2 cells treated with 20 mM valproc acid (VPA) for 16 hours or 0.01% DMSO as vehicle control. Data are means ± SD. ** p<0.01, ***P<0.001 (unpaired two-tailed student’s t test; N = 3 independent experiments per group). **(F)** Expression levels of FOXO3 in single-nuclei RNA-seq (snRNA-seq) analyses performed on >116,000 nuclei from n.19 COVID-19 autopsy lungs and n.7 pre-pandemic controls (Melms *et al*., 2021). **(G)** Genome browser screenshots showing accumulation of normalized FOXO3 signal, together with CromHMM state segmentation and H3K4me3 signal (ENCODE), along the ATP2A1 gene in human cells. ForCromHMM state segmentation colors indicate: Bright Red – Promoter; Orange and yellow - enhancer; Green - Transcriptional transition. The expanded view of the highlighted region, on the left, shows FOXO3 peaks over ATP2A1 enhancer regions, as marked by yellow region of CromHMM.

**Figure S4.**
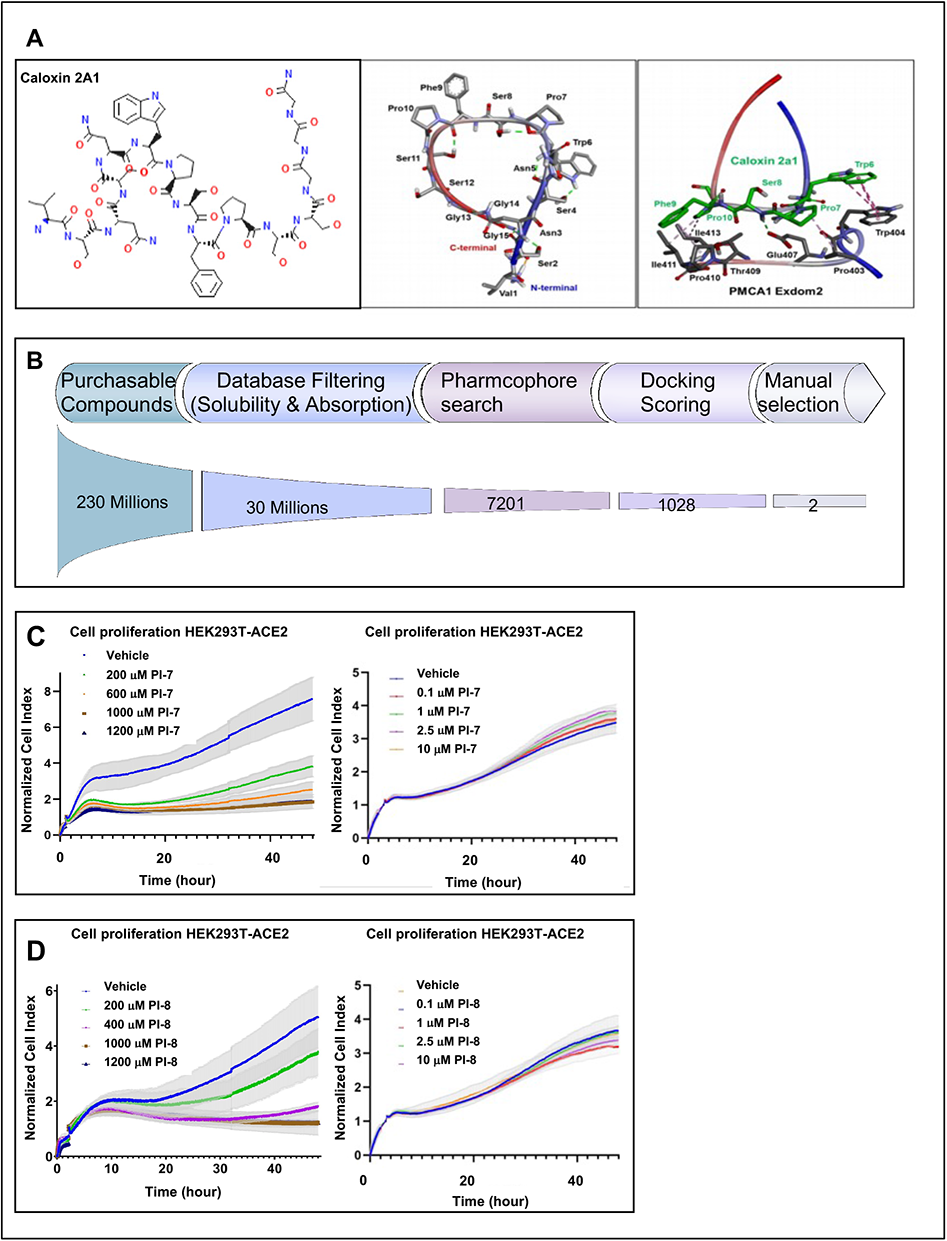
ATP2B1 impairment via Caloxin-derivative (PI-7) molecule. Related to Figure 4. **(A) On the left:** The sequence of caloxin2a1 sequence, as peptide, is shown. **On the right:** The molecular modeling of ATP2B1-caloxin2a1 structure by docking and energy minimization modeling via artificial intelligence as a drug design computational tool is shown. The pharmacophore model by using the structures ATP2B1-exodom-2 and caloxin 2a1 is also shown. Five pharmacophore features were produced. **(B)** The scheme of the pipeline of drug discovery are shown. Among 230 million purchasable compounds, 30 million molecules were selected by considering database filtering for solubility and absorption, 7,201 molecules by pharmacophore search, and 1028 molecules by docking scoring. The top 22 molecules were manually selected. Finally, two molecules were chosen. **(C, D)** Real-time cell proliferation analyses for the Cell Index (i.e., the cell-sensor impedance was expressed every two minutes as a unit called “Cell Index”). HEK293T-ACE2 were plated and, after two hours, treated with the indicated concentrations (escalating doses) of PI-7 **(C)** or PI-8 **(D)**; with vehicle-treated cells were the negative control. Impedance was measured every 2 min over 48 hours. The graphs showing “normalized cell index” were generated using Graph Pad Prism 9.

**Figure S5.**
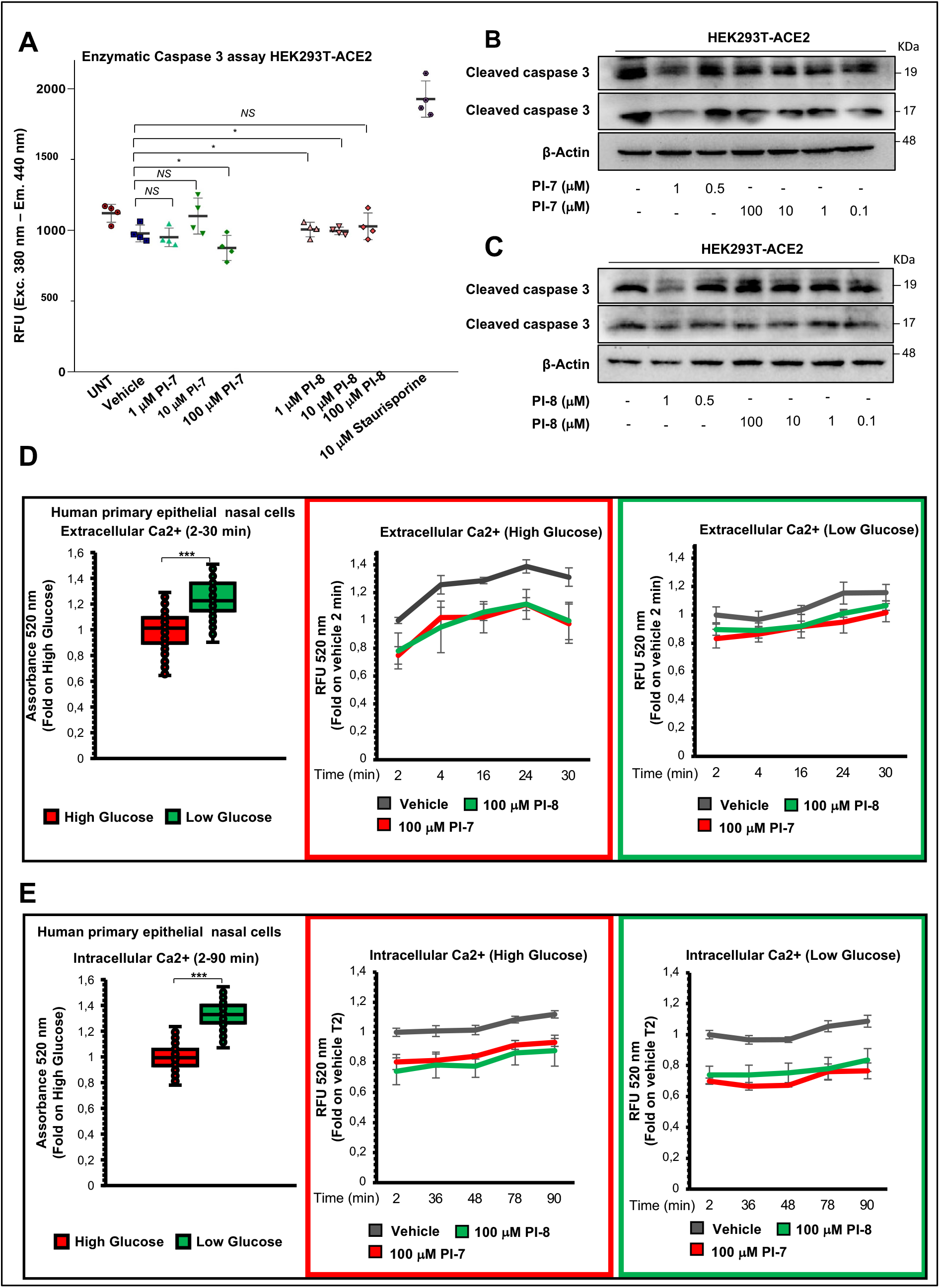
ATP2B1 impairment via Caloxin-derivative (PI-7) molecule diminishes extracellular and intracellular Ca^2+^ levels. Related to Figure 4. **(A)** Caspase 3 activity measured in HEK293T-ACE2 cells (5×10^5^ cells/well in a 24-well plate and incubated overnight) treated with increasing concentrations of compound PI-7 and PI-8 for 18 hours. Vehicle-treated cells and cells treated with 10 μM staurosporine were used as negative and positive controls, respectively. Data are presented as relative fluorescent units (RFUs; excitation: 380 nm; emission: 460 nm) measured after 1 hour of incubation (T1h). Data are means ± SD. \**P* < 0.05; N.S., nonsignificant (as determined by unpaired two-tailed Student’s *t* test; n = three independent experiments per group). **(B, C)** Representative immunoblotting analyses on total protein lysates obtained from HEK293T-ACE2 treated with escalating doses of PI-7 **(B)** and PI-8 **(C)** molecules using antibodies against the indicated proteins. β-Actin was used as the loading control. Vehicle-treated cells (i.e., 0.001% DMSO) are used as negative control of the experiment. All experiments were performed in triplicate. **(D) On the left:** Extracellular Ca^2+^ as measured using Fluo3-AM over 30 min in human primary epithelial nasal cells (1.2 ×10^4^ cells/well) plated in DMEM High Glucose or DMEM Low glucose for 24 hours. Data are means ± SD. ***P<0.001 (unpaired two-tailed student’s t test; N = 3 independent experiments per group). **On the right:** Extracellular Ca^2+^ as measured using Fluo3-AM over 30 min in human primary epithelial nasal cells (1.2 ×10^4^ cells/well) plated in DMEM High Glucose or DMEM Low glucose and treated with 100 μM compounds PI-7 and PI-8, or with 0.001% DMSO as vehicle. After 24 hours from the treatment started, the cell media supernatant was used for the extracellular Ca^2+^ measurement. The graph shows quantification of relative fluorescence changes of Fluo3 as a measure of extracellular Ca^2+^ levels by showing the relative fluorescent units (RFUs) (excitation, 506 nm; emission, 526 nm) recorded for 30 min by using a multimode plate reader (Enspire 2300, PerkinElmer). Red box: High Glucose; Green Box: Low glucose. Data are means ±SD of three independent experiments. **(E) On the left:** Intracellular Ca^2+^ as measured using Fluo3-AM over 90 min in human primary epithelial nasal cells (1.2 ×10^4^ cells/well) plated in DMEM High Glucose or DMEM Low glucose for 24 hours. Data are means ± SD. ***P<0.001 (unpaired two-tailed student’s t test; N = 3 independent experiments per group). **On the right:** Intracellular Ca^2+^ as measured using Fluo3-AM over 90 min in human primary epithelial nasal cells (1.2 ×10^4^ cells/well) plated in DMEM High Glucose or DMEM Low glucose and treated with 100 μM compounds PI-7 and PI-8, or with 0.001% DMSO as vehicle. After 24 hours from the treatment started, the cells were used for the intracellular Ca^2+^ measurement. The graph shows quantification of relative fluorescence changes of Fluo3 as a measure of intracellular Ca^2+^ levels by showing the relative fluorescent units (RFUs) (excitation, 506 nm; emission, 526 nm) recorded for 90 min by using a multimode plate reader (Enspire 2300, PerkinElmer). Red box: High Glucose; Green Box: Low glucose. Data are means ±SD of three independent experiments.

**Figure S6.**
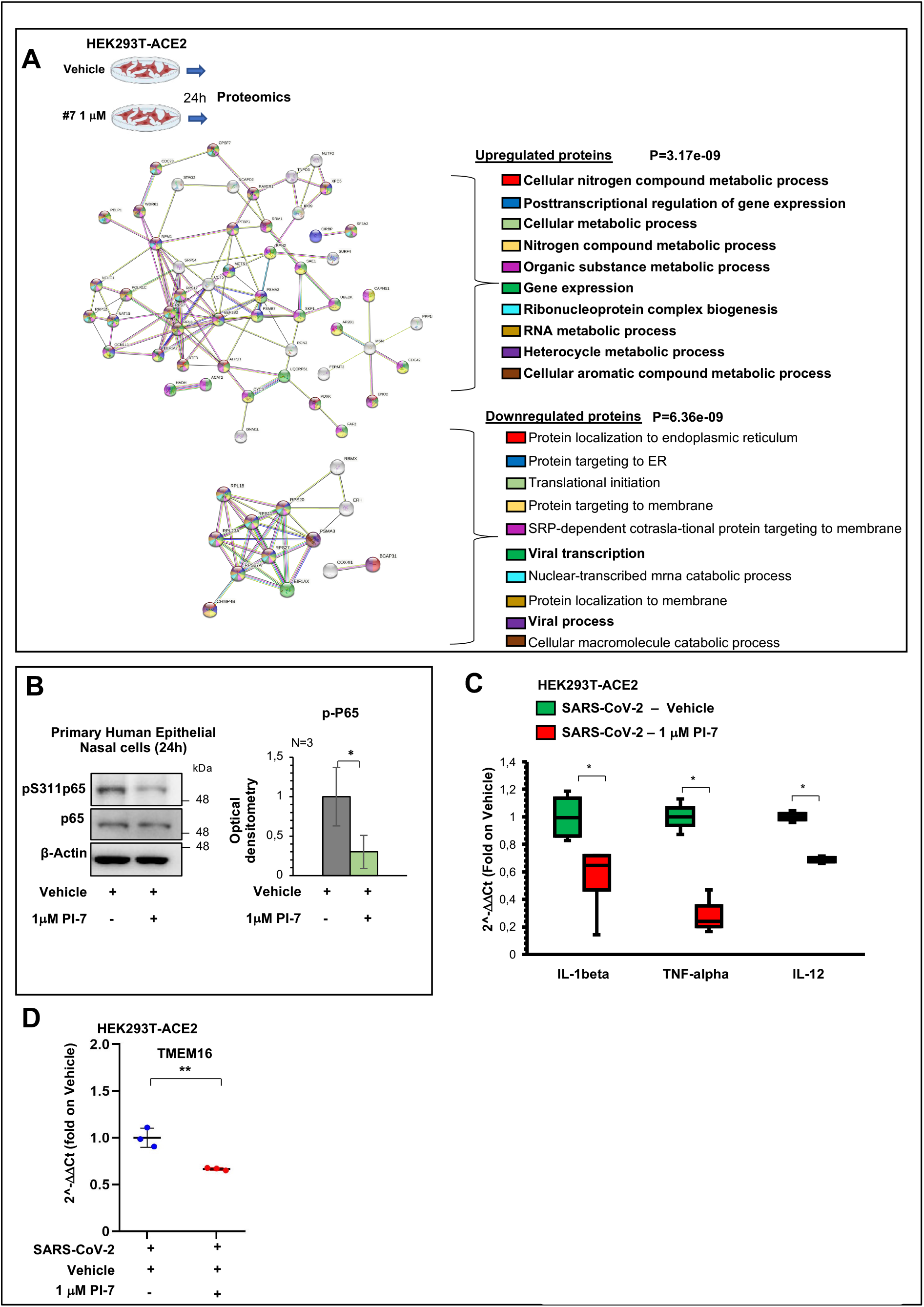
ATP2B1 impairment via Caloxin-derivative (PI-7) molecole diminishes SARS-CoV-2 propagation and syncytia formation. Related to Figures 5. **(A)** Proteomic assay based on LC-MS/MS approach performed on HEK-293T-ACE2 cells treated with PI-7 molecule (1 μM) for 24 hours. **On the top:** A protein interaction network was generated using the Search Tool for the Retrieval of Interacting Genes/ Proteins (STRING) database (https://string-db.org) by using only those proteins found upregulated in PI-7-treated cells (i.e., n.66 upregulated protein). **On the bottom:** A protein interaction network was generated using the Search Tool for the Retrieval of Interacting Genes/ Proteins (STRING) database (https://string-db.org), using only those proteins that were downregulated in PI-7-treated cells (i.e., 18 downregulated proteins). Among the pathways in which these downregulated proteins are involved, there were “viral process” and “viral transcription” (in bold). **(B)** Representative immunoblotting analysis using antibodies against the indicated proteins on total protein lysates obtained from HEK293T-ACE2 treated with compound PI-7 (1 μM) for 24 hours. All experiments were performed in triplicate. Densitometry analysis of the indicated band intensities in blots from three independent experiments. Data are means ±SD. * p <0.05 (unpaired two-tailed student’s t-tests; N = 3 independent experiments per group). **(C)** Quantification of mRNA abundance relative to that in control (CTR) cells (2−ΔΔCt) for IL-1beta, TNF-alpha and IL-12 genes. RT-PCR analysis of RNA extracted from HEK293T-ACE2 cells infected with SARS-CoV-2 (VOC Delta, 0.026 MOI for 72 hours) and treated with 1 μM PI-7 molecule or 0.001% DMSO as vehicle control. Data are means ± SD. * p<0.01 (unpaired two-tailed student’s t test; N = 3 independent experiments per group). **(D)** Quantification of mRNA abundance relative to that in control (CTR) cells (2^−ΔΔCt^) for human TMEM16 gene. RT-PCR analysis of RNA extracted from HEK293T-ACE2 cells treated as in (A). Data are means ± SD. * p < 0.05 (unpaired two-tailed student’s t test; N = 3 independent experiments per group).

### Supplementary Tables

**Table S1:**
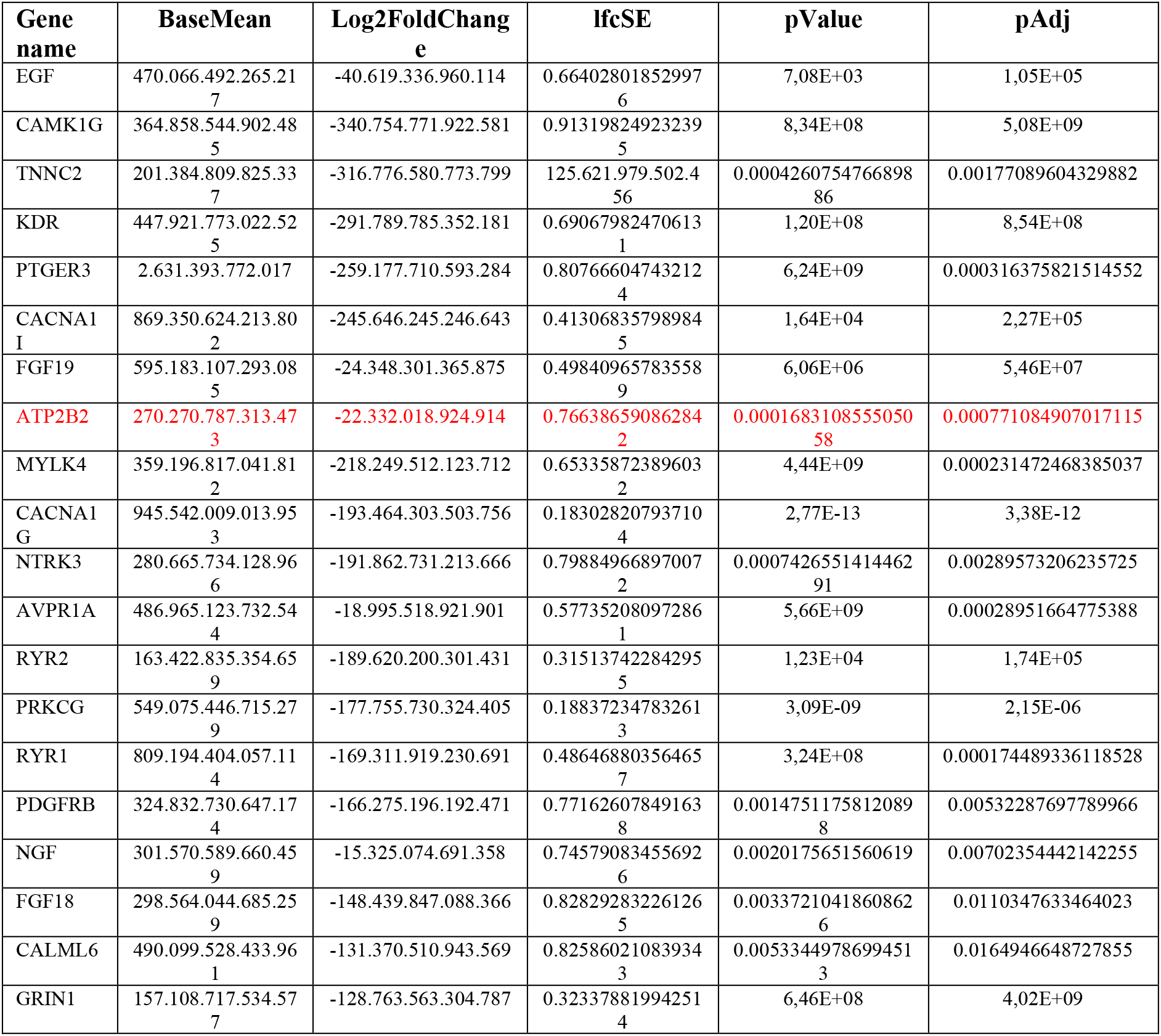
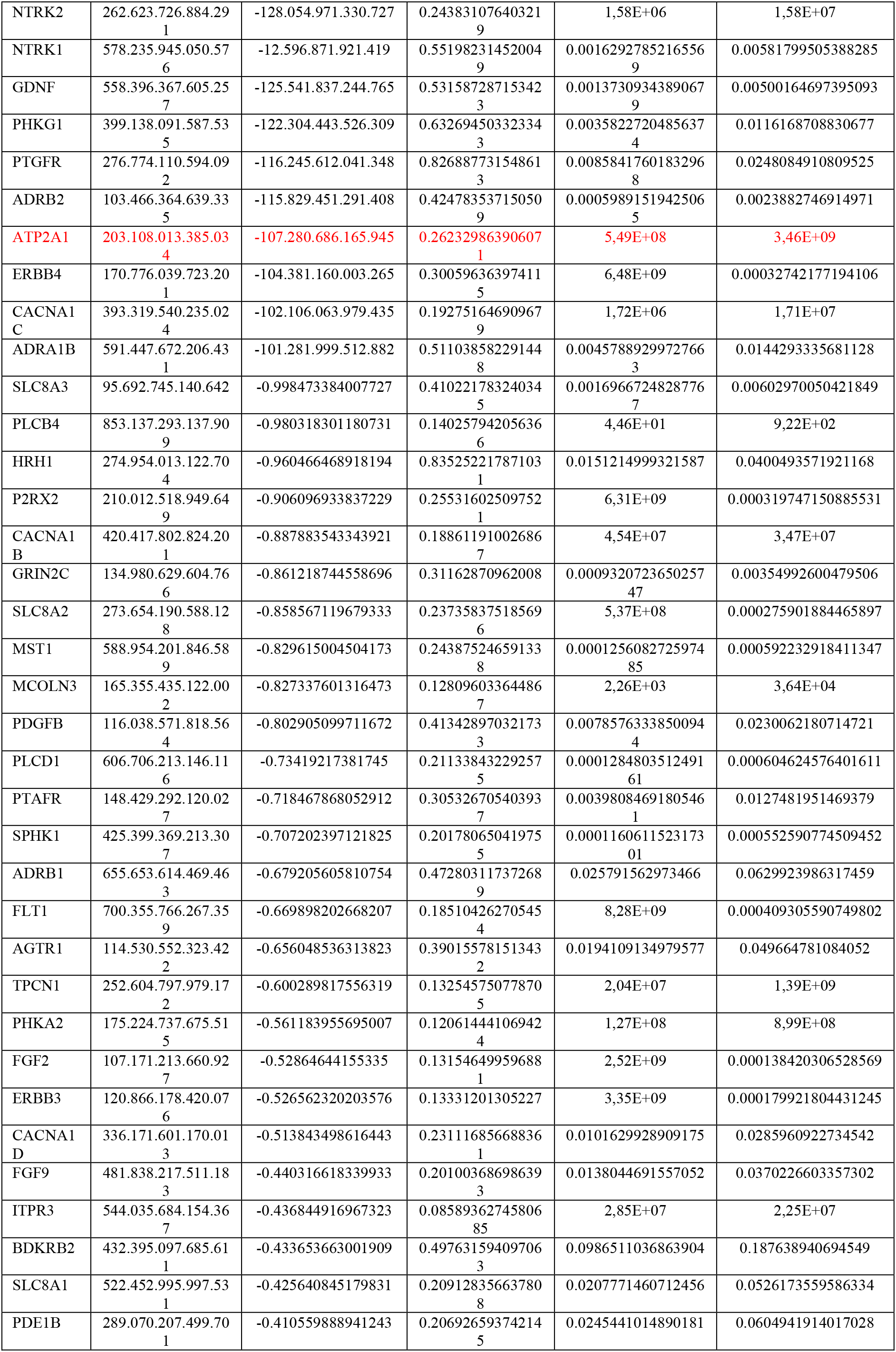
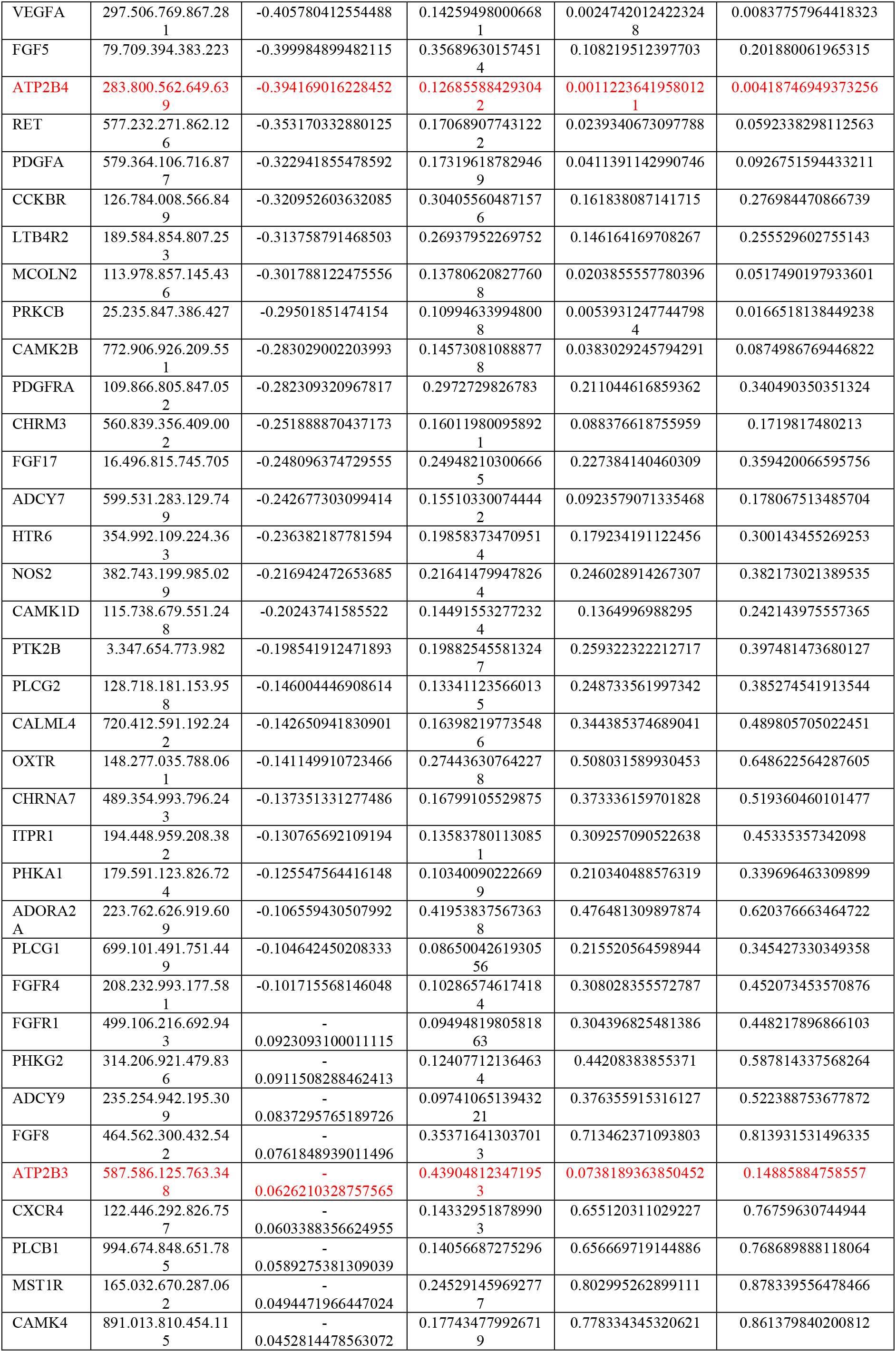
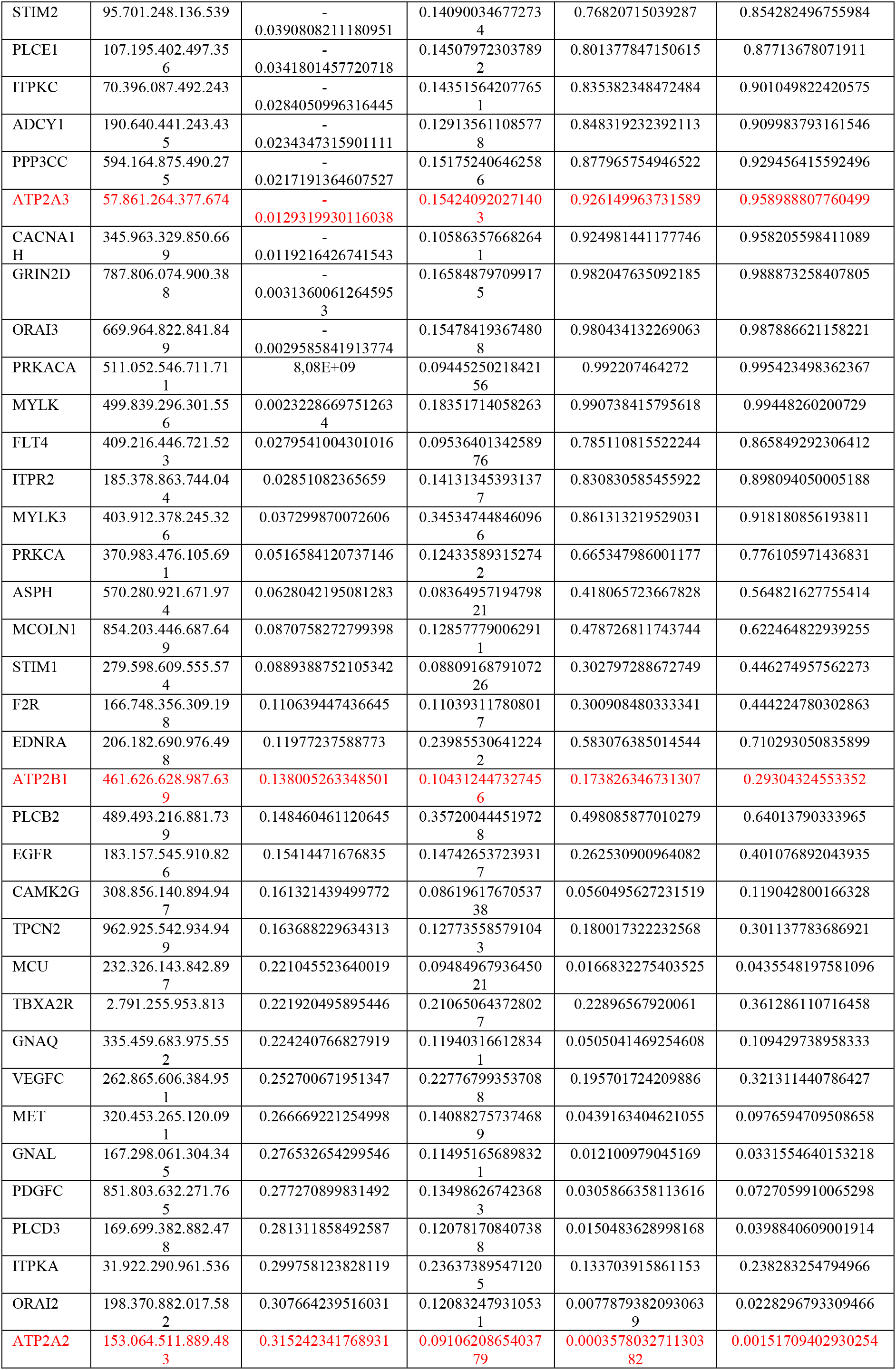
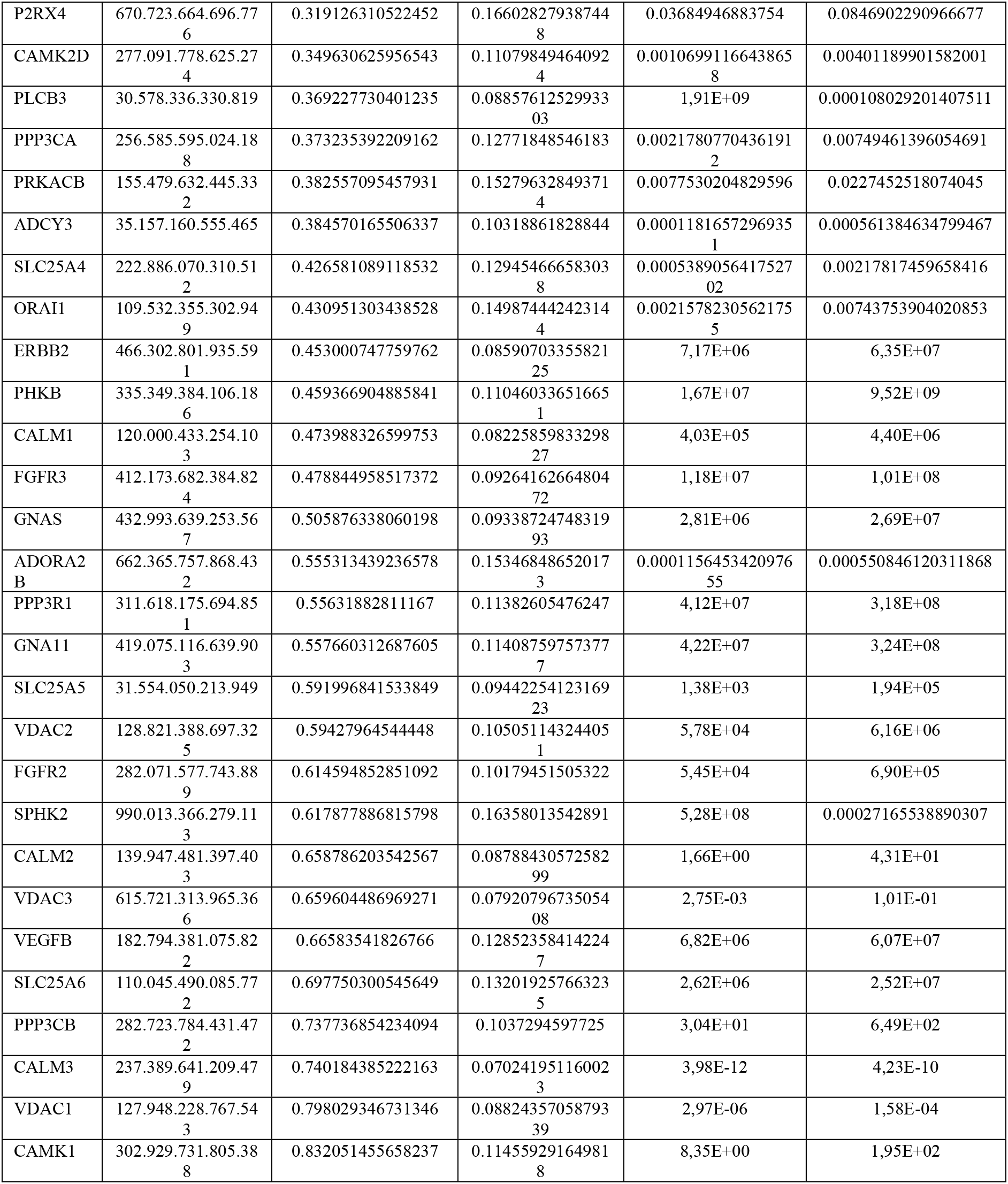
Differentially expressed genes (DEGs) upon SARS-CoV-2 early infection in HEK-293T-ACE2 taking part to Calcium Signalling. The genes taking part to “Calcium signalling” in REACTOME pathway enrichment analysis of those differentially expressed genes (DEGs) from RNAseq analyses performed in HEK293T-ACE2 cells infected with SARS-CoV-2 (VOC Delta, 24hours) vs untreated cells. N.157 genes statistically deregulated by SARS-CoV-2 after 24 hours involved in Calcium signalling were found. The gene name, the baseMean (the average of the normalized count values), the log2FoldChange, the lfcSE (standard error value), the pValue and the adjusted p values (i.e., padj) are listed. In red, the membrane and endoplasmic reticulum Ca2+ pumps (i.e., ATP2Bs and ATP2As).

**Table S2:**
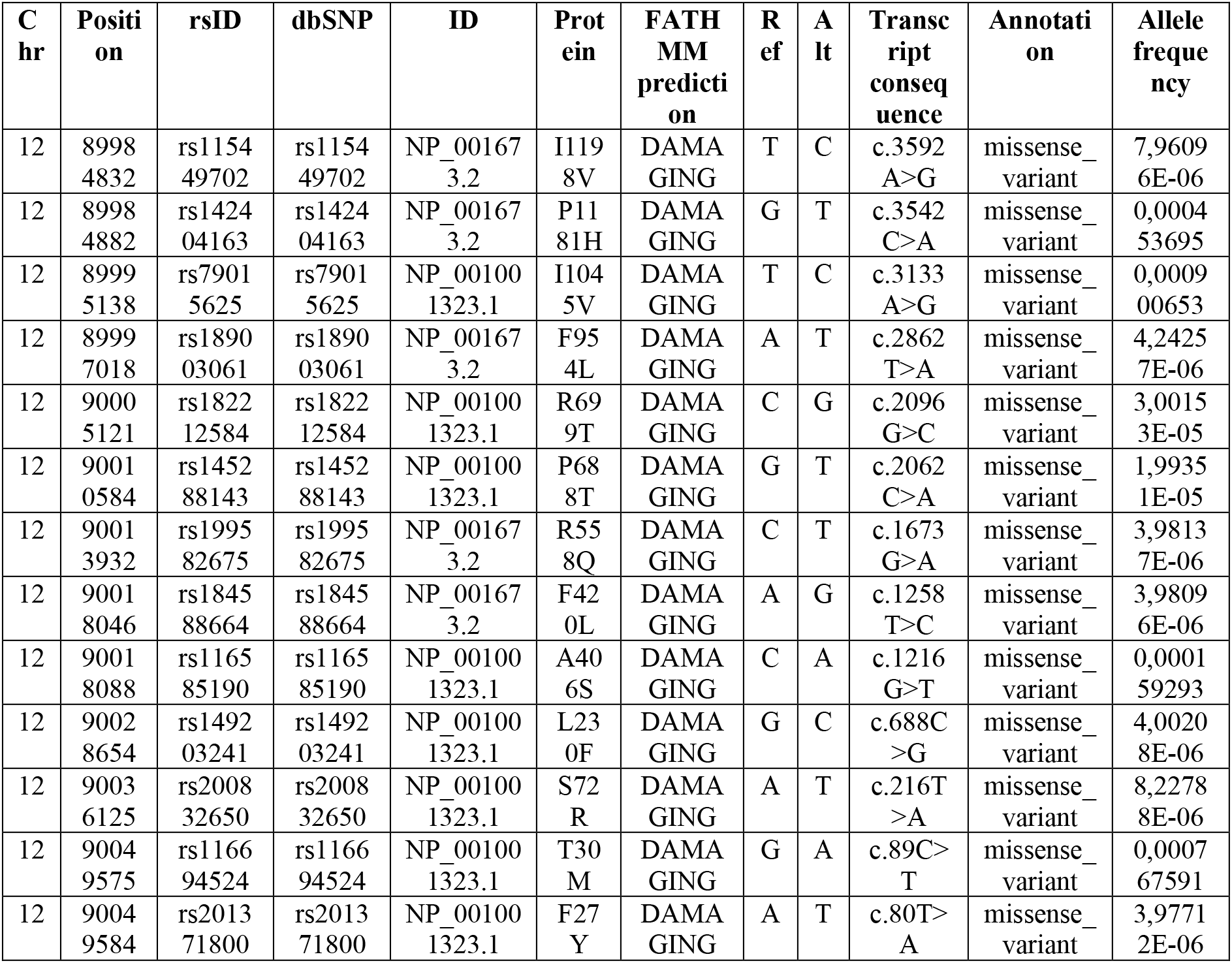
Rare coding pathogenic variants in ATP2B1 locus. Among the n.351 coding variants of ATP2B1 from “The Genome Aggregation Database” (GnomAD v2.1), n.13 are reported as “pathogenic” according to “Functional Analysis through Hidden Markov Models” (FATHMM) prediction score. These variants are extremely rare and are thus excluded from further investigations. The chromosomal locus, position, rsID, dbSNP ID, the protein consequence, the fathmm prediction, the reference and alternative alleles, the transcript consequence, the variant annotation and the allele frequency are listed.

**Table S3:** Non coding variants in the genomic region of the ATP2B1 locus acting as “expression quantitative traits loci” (eQTLs). The genecode ID, gene symbol, Variant ID (chromosomal position, alternative and reference alleles), the SNP ID, P-Value, normalized enrichment score (NES) and tissue (by using “Genotype-Tissue Expression” GTex database) are listed.

| Genecode Id | Gene symbol | Variant Id | SNP Id | P-value | NES | Tissue |
| --- | --- | --- | --- | --- | --- | --- |
| ENSG00000070961.15 | ATP2B1 | chr12 89615182 A G b38 | rs2681472 | 1,60E-18 | 0,35 | Artery - Tibial |
| ENSG00000070961.15 | ATP2B1 | chr12 89619312 T C b38 | rs2681492 | 3,40E-18 | 0,35 | Artery - Tibial |
| ENSG00000070961.15 | ATP2B1 | chr12 89632685 G A b38 | rs11105352 | 3,60E-18 | 0,35 | Artery - Tibial |
| ENSG00000070961.15 | ATP2B1 | chr12 89632686 C A b38 | rs11105353 | 3,60E-18 | 0,35 | Artery - Tibial |
| ENSG00000070961.15 | ATP2B1 | chr12 89632746 A G b38 | rs11105354 | 3,60E-18 | 0,35 | Artery - Tibial |
| ENSG00000070961.15 | ATP2B1 | chr12 89656726 A G b38 | rs12579302 | 3,60E-18 | 0,35 | Artery - Tibial |
| ENSG00000070961.15 | ATP2B1 | chr12 89665065 G A b38 | rs111478946 | 3,60E-18 | 0,35 | Artery - Tibial |
| ENSG00000070961.15 | ATP2B1 | chr12 89675499 T G b38 | rs11105364 | 3,60E-18 | 0,35 | Artery - Tibial |
| ENSG00000070961.15 | ATP2B1 | chr12 89680664 G C b38 | rs11105368 | 7,40E-18 | 0,34 | Artery - Tibial |
| ENSG00000070961.15 | ATP2B1 | chr12 89681466 C T b38 | rs4842675 | 7,40E-18 | 0,34 | Artery - Tibial |
| ENSG00000070961.15 | ATP2B1 | chr12 89591515 AAAGT A b38 | rs10580742 | 1,20E-17 | 0,35 | Artery - Tibial |
| ENSG00000070961.15 | ATP2B1 | chr12 89696964 C T b38 | rs11105378 | 1,30E-17 | 0,34 | Artery - Tibial |
| ENSG00000070961.15 | ATP2B1 | chr12 89666809 G A b38 | rs17249754 | 1,50E-17 | 0,34 | Artery - Tibial |
| ENSG00000070961.15 | ATP2B1 | chr12 89668599 G A b38 | rs6538195 | 1,50E-17 | 0,34 | Artery - Tibial |
| ENSG00000070961.15 | ATP2B1 | chr12 89678299 G A b38 | rs73437358 | 1,50E-17 | 0,34 | Artery - Tibial |
| ENSG00000070961.15 | ATP2B1 | chr12 89625401 C G b38 | rs57481061 | 1,50E-17 | 0,34 | Artery - Tibial |
| ENSG00000070961.15 | ATP2B1 | chr12 89548613 A G b38 | rs11105328 | 1,70E-17 | 0,34 | Artery - Tibial |
| ENSG00000070961.15 | ATP2B1 | chr12 89547772 T C b38 | rs4842666 | 2,10E-17 | 0,34 | Artery - Tibial |
| ENSG00000070961.15 | ATP2B1 | chr12 89694803 T A b38 | rs11105375 | 5,70E-17 | 0,34 | Artery - Tibial |
| ENSG00000070961.15 | ATP2B1 | chr12 89697090 A G b38 | rs12230074 | 8,10E-17 | 0,33 | Artery - Tibial |
| ENSG00000070961.15 | ATP2B1 | chr12 89660842 T C b38 | rs73437338 | 1,50E-16 | 0,33 | Artery - Tibial |
| ENSG00000070961.15 | ATP2B1 | chr12 89597536 C T b38 | rs117742247 | 4,20E-16 | 0,34 | Artery - Tibial |
| ENSG00000070961.15 | ATP2B1 | chr12 89599730 A T b38 | rs11105337 | 4,60E-16 | 0,33 | Artery - Tibial |
| ENSG00000070961.15 | ATP2B1 | chr12 89694991 G A b38 | rs11105376 | 7,80E-16 | 0,33 | Artery - Tibial |
| ENSG00000070961.15 | ATP2B1 | chr12 89709925 C G b38 | rs2280715 | 4,30E-14 | 0,31 | Artery - Tibial |
| ENSG00000070961.15 | ATP2B1 | chr12 89713495 T C b38 | rs11105382 | 4,30E-14 | 0,31 | Artery - Tibial |
| ENSG00000070961.15 | ATP2B1 | chr12 89713529 T C b38 | rs11105383 | 4,30E-14 | 0,31 | Artery - Tibial |
| ENSG00000070961.15 | ATP2B1 | chr12 89719293 T G b38 | rs7299436 | 4,30E-14 | 0,31 | Artery - Tibial |
| ENSG00000070961.15 | ATP2B1 | chr12 89709168 C G b38 | rs73437382 | 8,00E-14 | 0,3 | Artery - Tibial |
| ENSG00000070961.15 | ATP2B1 | chr12 89698005 G C b38 | rs4842676 | 6,10E-13 | -0,27 | Artery - Tibial |
| ENSG00000070961.15 | ATP2B1 | chr12 89701396 T C b38 | rs11105379 | 1,40E-12 | 0,3 | Artery - Tibial |
| ENSG00000070961.15 | ATP2B1 | chr12 89695013 G A b38 | rs10858917 | 2,50E-12 | -0,26 | Artery - Tibial |
| ENSG00000070961.15 | ATP2B1 | chr12 89556543 A C b38 | rs7302816 | 4,50E-12 | 0,25 | Artery - Tibial |
| ENSG00000070961.15 | ATP2B1 | chr12 89713058 A G b38 | rs11105381 | 2,90E-11 | -0,25 | Artery - Tibial |
| ENSG00000070961.15 | ATP2B1 | chr12 89715560 T C b38 | rs2408046 | 5,50E-11 | -0,24 | Artery - Tibial |
| ENSG00000070961.15 | ATP2B1 | chr12 89703890 G C b38 | rs10745511 | 5,70E-11 | -0,25 | Artery - Tibial |
| ENSG00000070961.15 | ATP2B1 | chr12 89705620 T A b38 | rs2113894 | 5,70E-11 | -0,25 | Artery - Tibial |
| ENSG00000070961.15 | ATP2B1 | chr12 89696509 CAA C b38 | rs10550903 | 7,60E-11 | -0,23 | Artery - Tibial |
| ENSG00000070961.15 | ATP2B1 | chr12 89738370 G A b38 | rs12579052 | 5,40E-10 | 0,28 | Artery - Tibial |
| ENSG00000070961.15 | ATP2B1 | chr12 90001084 A T b38 | rs4408360 | 3,40E-08 | -0,18 | Esophagus - Mucosa |
| ENSG00000070961.15 | ATP2B1 | chr12_90024395_T_G_b38 | rs149193253 | 4,40E-08 | 1 | Brain - Anterior cingulate cortex (BA24) |
| ENSG00000070961.15 | ATP2B1 | chr12_90049701_T_C_b38 | rs150184539 | 4,40E-08 | 1 | Brain - Anterior cingulate cortex (BA24) |
| ENSG00000070961.15 | ATP2B1 | chr12_90137707_G_A_b38 | rs77187603 | 4,40E-08 | 1 | Brain - Anterior cingulate cortex (BA24) |
| ENSG00000070961.15 | ATP2B1 | chr12_90143905_A_G_b38 | rs150119566 | 4,40E-08 | 1 | Brain - Anterior cingulate cortex (BA24) |
| ENSG00000070961.15 | ATP2B1 | chr12 89677042 CAAAG C b38 | rs143087380 | 5,00E-08 | -0,17 | Artery - Tibial |
| ENSG00000070961.15 | ATP2B1 | chr12 90006794 TA T b38 | rs67882152 | 8,80E-08 | -0,18 | Esophagus - Mucosa |
| ENSG00000070961.15 | ATP2B1 | chr12 90006796 A T b38 | rs114744356 | 8,80E-08 | -0,18 | Esophagus - Mucosa |
| ENSG00000070961.15 | ATP2B1 | chr12 90010274 T C b38 | rs7488223 | 1,10E-07 | -0,17 | Esophagus - Mucosa |
| ENSG00000070961.15 | ATP2B1 | chr12 89973923 G C b38 | rs4842701 | 1,30E-07 | -0,17 | Esophagus - Mucosa |
| ENSG00000070961.15 | ATP2B1 | chr12 89997003 T C b38 | rs7968803 | 1,30E-07 | -0,18 | Esophagus - Mucosa |
| ENSG00000070961.15 | ATP2B1 | chr12 89991517 A G b38 | rs10777217 | 1,60E-07 | -0,17 | Esophagus - Mucosa |
| ENSG00000070961.15 | ATP2B1 | chr12 89595822 G A b38 | rs1401982 | 2,30E-07 | -0,16 | Artery - Tibial |
| ENSG00000070961.15 | ATP2B1 | chr12 89576142 A AACTC b38 | rs10689649 | 2,50E-07 | 0,16 | Artery - Tibial |
| ENSG00000070961.15 | ATP2B1 | chr12 89968275 A G b38 | rs4466894 | 2,70E-07 | -0,17 | Esophagus - Mucosa |
| ENSG00000070961.15 | ATP2B1 | chr12 89584456 C T b38 | rs1689040 | 2,80E-07 | 0,16 | Artery - Tibial |
| ENSG00000070961.15 | ATP2B1 | chr12 89951830 C T b38 | rs4842699 | 3,00E-07 | -0,17 | Esophagus - Mucosa |
| ENSG00000070961.15 | ATP2B1 | chr12 89631845 G A b38 | rs2681485 | 3,30E-07 | -0,16 | Artery - Tibial |
| ENSG00000070961.15 | ATP2B1 | chr12 90140138 C T b38 | rs55849728 | 3,80E-07 | -0,18 | Esophagus - Mucosa |
| ENSG00000070961.15 | ATP2B1 | chr12 89571272 C T b38 | rs7313874 | 3,90E-07 | 0,16 | Artery - Tibial |
| ENSG00000070961.15 | ATP2B1 | chr12 89434814 A G b38 | rs8181784 | 4,20E-07 | 0,17 | Artery - Tibial |
| ENSG00000070961.15 | ATP2B1 | chr12 89435693 C T b38 | rs11105287 | 4,20E-07 | 0,17 | Artery - Tibial |
| ENSG00000070961.15 | ATP2B1 | chr12 89569762 G GAGA b38 | rs35533953 | 4,20E-07 | 0,16 | Artery - Tibial |
| ENSG00000070961.15 | ATP2B1 | chr12 89687411 T C b38 | rs7136259 | 5,00E-07 | -0,16 | Artery - Tibial |
| ENSG00000070961.15 | ATP2B1 | chr12 89702568 T C b38 | rs10858918 | 5,00E-07 | -0,17 | Artery - Tibial |
| ENSG00000070961.15 | ATP2B1 | chr12 89943269 A C b38 | rs4842697 | 5,20E-07 | -0,17 | Esophagus - Mucosa |
| ENSG00000070961.15 | ATP2B1 | chr12 89436943 T C b38 | rs10777163 | 6,60E-07 | 0,17 | Artery - Tibial |
| ENSG00000070961.15 | ATP2B1 | chr12 89643729 T C b38 | rs111337717 | 7,20E-07 | 0,35 | Artery - Tibial |
| ENSG00000070961.15 | ATP2B1 | chr12 89713948 C T b38 | rs1590008 | 7,20E-07 | -0,16 | Artery - Tibial |
| ENSG00000070961.15 | ATP2B1 | chr12 89717005 G A b38 | rs1980235 | 7,20E-07 | -0,16 | Artery - Tibial |
| ENSG00000070961.15 | ATP2B1 | chr12 90156621 AG A b38 | rs369917221 | 7,60E-07 | 0,75 | Artery - Tibial |
| ENSG00000070961.15 | ATP2B1 | chr12 90025415 G A b38 | rs10858944 | 8,30E-07 | -0,16 | Esophagus - Mucosa |
| ENSG00000070961.15 | ATP2B1 | chr12 90034251 A G b38 | rs73208536 | 8,40E-07 | -0,16 | Esophagus - Mucosa |
| ENSG00000070961.15 | ATP2B1 | chr12_90385245_G_A_b38 | rs1594565 | 8,60E-07 | 0,38 | Brain - Spinal cord (cervical c-1) |
| ENSG00000070961.15 | ATP2B1 | chr12 90047438 T C b38 | rs10777221 | 8,70E-07 | -0,16 | Esophagus - Mucosa |
| ENSG00000070961.15 | ATP2B1 | chr12 90048224 G A b38 | rs7488974 | 8,70E-07 | -0,16 | Esophagus - Mucosa |
| ENSG00000070961.15 | ATP2B1 | chr12 89987295 A ATGTT b38 | rs10625779 | 9,80E-07 | -0,16 | Esophagus - Mucosa |
| ENSG00000070961.15 | ATP2B1 | chr12 89708115 T A b38 | rs1358350 | 9,90E-07 | -0,16 | Artery - Tibial |
| ENSG00000070961.15 | ATP2B1 | chr12 89640625 G GC b38 | rs66786558 | 0,0000011 | -0,16 | Artery - Tibial |
| ENSG00000070961.15 | ATP2B1 | chr12 89640630 T C b38 | rs78409878 | 0,0000011 | -0,16 | Artery - Tibial |
| ENSG00000070961.15 | ATP2B1 | chr12 89648365 C G b38 | rs11105358 | 0,0000011 | -0,16 | Artery - Tibial |
| ENSG00000070961.15 | ATP2B1 | chr12_90398938_C_T_b38 | rs7955801 | 0,0000012 | 0,38 | Brain - Spinal cord (cervical c-1) |
| ENSG00000070961.15 | ATP2B1 | chr12_90392860_A_C_b38 | rs2116497 | 0,0000012 | 0,37 | Brain - Spinal cord (cervical c-1) |
| ENSG00000070961.15 | ATP2B1 | chr12 89437093 C T b38 | rs10858851 | 0,0000012 | 0,17 | Artery - Tibial |
| ENSG00000070961.15 | ATP2B1 | chr12 90072266 A G b38 | rs6538223 | 0,0000012 | -0,16 | Esophagus - Mucosa |
| ENSG00000070961.15 | ATP2B1 | chr12 90168240 C T b38 | rs4842509 | 0,0000012 | -0,17 | Esophagus - Mucosa |
| ENSG00000070961.15 | ATP2B1 | chr12 90170518 A G b38 | rs17836934 | 0,0000014 | -0,17 | Esophagus - Mucosa |
| ENSG00000070961.15 | ATP2B1 | chr12 89482366 T C b38 | rs11105298 | 0,0000015 | 0,17 | Artery - Tibial |
| ENSG00000070961.15 | ATP2B1 | chr12_90391772_A_C_b38 | rs7976895 | 0,0000016 | 0,37 | Brain - Spinal cord (cervical c-1) |
| ENSG00000070961.15 | ATP2B1 | chr12_90393455_G_A_b38 | rs12580873 | 0,0000016 | 0,37 | Brain - Spinal cord (cervical c-1) |
| ENSG00000070961.15 | ATP2B1 | chr12 90084874 G A b38 | rs10858948 | 0,0000016 | -0,15 | Esophagus - Mucosa |
| ENSG00000070961.15 | ATP2B1 | chr12 90079719 A G b38 | rs6538224 | 0,0000017 | -0,16 | Esophagus - Mucosa |
| ENSG00000070961.15 | ATP2B1 | chr12_90376827_T_C_b38 | rs11105589 | 0,0000018 | 0,37 | Brain - Spinal cord (cervical c-1) |
| ENSG00000070961.15 | ATP2B1 | chr12_90386667_C_T_b38 | rs6538234 | 0,0000018 | 0,37 | Brain - Spinal cord (cervical c-1) |
| ENSG00000070961.15 | ATP2B1 | chr12_90388244_T_A_b38 | rs7979392 | 0,0000018 | 0,37 | Brain - Spinal cord (cervical c-1) |
| ENSG00000070961.15 | ATP2B1 | chr12_90390285_G_C_b38 | rs2043213 | 0,0000018 | 0,37 | Brain - Spinal cord (cervical c-1) |
| ENSG00000070961.15 | ATP2B1 | chr12 89526956 C T b38 | rs11105312 | 0,0000018 | 0,16 | Artery - Tibial |
| ENSG00000070961.15 | ATP2B1 | chr12 89527498 C T b38 | rs10777185 | 0,0000018 | 0,16 | Artery - Tibial |
| ENSG00000070961.15 | ATP2B1 | chr12 89625452 C T b38 | rs7297206 | 0,0000018 | -0,15 | Artery - Tibial |
| ENSG00000070961.15 | ATP2B1 | chr12 90059201 T C b38 | rs7959983 | 0,0000021 | -0,16 | Esophagus - Mucosa |
| ENSG00000070961.15 | ATP2B1 | chr12 90080698 G A b38 | rs7485419 | 0,0000021 | -0,16 | Esophagus - Mucosa |
| ENSG00000070961.15 | ATP2B1 | chr12_90376071_T_C_b38 | rs12582977 | 0,0000023 | 0,37 | Brain - Spinal cord (cervical c-1) |
| ENSG00000070961.15 | ATP2B1 | chr12 89533400 G A b38 | rs11105319 | 0,0000024 | 0,16 | Artery - Tibial |
| ENSG00000070961.15 | ATP2B1 | chr12 89534012 A G b38 | rs10858902 | 0,0000024 | 0,16 | Artery - Tibial |
| ENSG00000070961.15 | ATP2B1 | chr12 89534195 G A b38 | rs10858903 | 0,0000024 | 0,16 | Artery - Tibial |
| ENSG00000070961.15 | ATP2B1 | chr12 89539669 T C b38 | rs4842662 | 0,0000024 | 0,16 | Artery - Tibial |
| ENSG00000070961.15 | ATP2B1 | chr12 89539986 G C b38 | rs3741898 | 0,0000024 | 0,16 | Artery - Tibial |
| ENSG00000070961.15 | ATP2B1 | chr12 89540697 C T b38 | rs10858906 | 0,0000024 | 0,16 | Artery - Tibial |
| ENSG00000070961.15 | ATP2B1 | chr12 89541074 CAGAG C b38 | rs71897440 | 0,0000024 | 0,16 | Artery - Tibial |
| ENSG00000070961.15 | ATP2B1 | chr12 89541195 G C b38 | rs12369195 | 0,0000024 | 0,16 | Artery - Tibial |
| ENSG00000070961.15 | ATP2B1 | chr12 89528255 C A b38 | rs10858897 | 0,0000025 | 0,16 | Artery - Tibial |
| ENSG00000070961.15 | ATP2B1 | chr12 89535128 A G b38 | rs12369685 | 0,0000026 | 0,17 | Artery - Tibial |
| ENSG00000070961.15 | ATP2B1 | chr12 90068229 T C b38 | rs7310135 | 0,0000026 | -0,15 | Esophagus - Mucosa |
| ENSG00000070961.15 | ATP2B1 | chr12 90079906 G A b38 | rs6419356 | 0,0000026 | -0,15 | Esophagus - Mucosa |
| ENSG00000070961.15 | ATP2B1 | chr12 90081169 G A b38 | rs7962435 | 0,0000026 | -0,15 | Esophagus - Mucosa |
| ENSG00000070961.15 | ATP2B1 | chr12 90084513 C T b38 | rs7132441 | 0,0000026 | -0,15 | Esophagus - Mucosa |
| ENSG00000070961.15 | ATP2B1 | chr12 89530034 G C b38 | rs10858899 | 0,0000027 | 0,16 | Artery - Tibial |
| ENSG00000070961.15 | ATP2B1 | chr12 89530596 A ATCT b38 | rs112698976 | 0,0000027 | 0,16 | Artery - Tibial |
| ENSG00000070961.15 | ATP2B1 | chr12 89530782 C T b38 | rs11105314 | 0,0000027 | 0,16 | Artery - Tibial |
| ENSG00000070961.15 | ATP2B1 | chr12 89531423 T G b38 | rs11105316 | 0,0000027 | 0,16 | Artery - Tibial |
| ENSG00000070961.15 | ATP2B1 | chr12 89933142 G A b38 | rs11105466 | 0,0000028 | -0,16 | Esophagus - Mucosa |
| ENSG00000070961.15 | ATP2B1 | chr12 89432148 T C b38 | rs10858848 | 0,0000029 | 0,16 | Artery - Tibial |
| ENSG00000070961.15 | ATP2B1 | chr12 89497589 G A b38 | rs11105302 | 0,000003 | 0,16 | Artery - Tibial |
| ENSG00000070961.15 | ATP2B1 | chr12 89497697 T G b38 | rs10858881 | 0,000003 | 0,16 | Artery - Tibial |
| ENSG00000070961.15 | ATP2B1 | chr12 89441951 C A b38 | rs10858854 | 0,0000032 | 0,17 | Artery - Tibial |
| ENSG00000070961.15 | ATP2B1 | chr12 89445295 C T b38 | rs11105291 | 0,0000032 | 0,17 | Artery - Tibial |
| ENSG00000070961.15 | ATP2B1 | chr12 89445926 A G b38 | rs10858860 | 0,0000032 | 0,17 | Artery - Tibial |
| ENSG00000070961.15 | ATP2B1 | chr12 90049705 A G b38 | rs10858945 | 0,0000032 | -0,15 | Esophagus - Mucosa |
| ENSG00000070961.15 | ATP2B1 | chr12 90050234 G T b38 | rs7958749 | 0,0000032 | -0,15 | Esophagus - Mucosa |
| ENSG00000070961.15 | ATP2B1 | chr12 89481583 G A b38 | rs75615848 | 0,0000035 | 0,32 | Artery - Tibial |
| ENSG00000070961.15 | ATP2B1 | chr12 89439561 T C b38 | rs10858852 | 0,0000036 | 0,16 | Artery - Tibial |
| ENSG00000070961.15 | ATP2B1 | chr12 89440336 C T b38 | rs4842653 | 0,0000036 | 0,16 | Artery - Tibial |
| ENSG00000070961.15 | ATP2B1 | chr12 89440651 T A b38 | rs4842654 | 0,0000036 | 0,16 | Artery - Tibial |
| ENSG00000070961.15 | ATP2B1 | chr12 89442082 C G b38 | rs10858855 | 0,0000036 | 0,16 | Artery - Tibial |
| ENSG00000070961.15 | ATP2B1 | chr12 89442149 G A b38 | rs9795772 | 0,0000036 | 0,16 | Artery - Tibial |
| ENSG00000070961.15 | ATP2B1 | chr12 89442895 G C b38 | rs111513812 | 0,0000036 | 0,16 | Artery - Tibial |
| ENSG00000070961.15 | ATP2B1 | chr12 89443404 T C b38 | rs11105289 | 0,0000036 | 0,16 | Artery - Tibial |
| ENSG00000070961.15 | ATP2B1 | chr12 89445026 A T b38 | rs11105290 | 0,0000036 | 0,16 | Artery - Tibial |
| ENSG00000070961.15 | ATP2B1 | chr12 89445465 C T b38 | rs10858859 | 0,0000036 | 0,16 | Artery - Tibial |
| ENSG00000070961.15 | ATP2B1 | chr12 89445949 A G b38 | rs10858861 | 0,0000036 | 0,16 | Artery - Tibial |
| ENSG00000070961.15 | ATP2B1 | chr12 89448276 T C b38 | rs10858862 | 0,0000036 | 0,16 | Artery - Tibial |
| ENSG00000070961.15 | ATP2B1 | chr12 89451080 T TG b38 | rs11461256 | 0,0000036 | 0,16 | Artery - Tibial |
| ENSG00000070961.15 | ATP2B1 | chr12 89452453 C T b38 | rs10858864 | 0,0000036 | 0,16 | Artery - Tibial |
| ENSG00000070961.15 | ATP2B1 | chr12 89452699 G A b38 | rs10858865 | 0,0000036 | 0,16 | Artery - Tibial |
| ENSG00000070961.15 | ATP2B1 | chr12 89457389 C T b38 | rs10858867 | 0,0000036 | 0,16 | Artery - Tibial |
| ENSG00000070961.15 | ATP2B1 | chr12 89463834 C T b38 | rs10858869 | 0,0000036 | 0,16 | Artery - Tibial |
| ENSG00000070961.15 | ATP2B1 | chr12 89466542 C A b38 | rs3803128 | 0,0000036 | 0,16 | Artery - Tibial |
| ENSG00000070961.15 | ATP2B1 | chr12 89467752 T C b38 | rs10506972 | 0,0000036 | 0,16 | Artery - Tibial |
| ENSG00000070961.15 | ATP2B1 | chr12 89468253 G T b38 | rs12228177 | 0,0000036 | 0,16 | Artery - Tibial |
| ENSG00000070961.15 | ATP2B1 | chr12 89477836 G A b38 | rs10858871 | 0,0000036 | 0,16 | Artery - Tibial |
| ENSG00000070961.15 | ATP2B1 | chr12 89479155 C T b38 | rs10858872 | 0,0000036 | 0,16 | Artery - Tibial |
| ENSG00000070961.15 | ATP2B1 | chr12 89483146 C T b38 | rs112511312 | 0,0000036 | 0,16 | Artery - Tibial |
| ENSG00000070961.15 | ATP2B1 | chr12 89484166 A G b38 | rs11105299 | 0,0000036 | 0,16 | Artery - Tibial |
| ENSG00000070961.15 | ATP2B1 | chr12 89487787 C G b38 | rs4842488 | 0,0000036 | 0,16 | Artery - Tibial |
| ENSG00000070961.15 | ATP2B1 | chr12 89490501 C T b38 | rs10858877 | 0,0000036 | 0,16 | Artery - Tibial |
| ENSG00000070961.15 | ATP2B1 | chr12 89490517 G A b38 | rs10858878 | 0,0000036 | 0,16 | Artery - Tibial |
| ENSG00000070961.15 | ATP2B1 | chr12 89492146 C A b38 | rs6538189 | 0,0000036 | 0,16 | Artery - Tibial |
| ENSG00000070961.15 | ATP2B1 | chr12 89492165 T C b38 | rs6538190 | 0,0000036 | 0,16 | Artery - Tibial |
| ENSG00000070961.15 | ATP2B1 | chr12 89492801 T A b38 | rs10858879 | 0,0000036 | 0,16 | Artery - Tibial |
| ENSG00000070961.15 | ATP2B1 | chr12 89493872 T C b38 | rs1008764 | 0,0000036 | 0,16 | Artery - Tibial |
| ENSG00000070961.15 | ATP2B1 | chr12 89503120 C T b38 | rs10858885 | 0,0000036 | 0,16 | Artery - Tibial |
| ENSG00000070961.15 | ATP2B1 | chr12 89503384 C G b38 | rs10858886 | 0,0000036 | 0,16 | Artery - Tibial |
| ENSG00000070961.15 | ATP2B1 | chr12 89503557 C T b38 | rs11105305 | 0,0000036 | 0,16 | Artery - Tibial |
| ENSG00000070961.15 | ATP2B1 | chr12 89503611 C T b38 | rs11105306 | 0,0000036 | 0,16 | Artery - Tibial |
| ENSG00000070961.15 | ATP2B1 | chr12 89508827 G A b38 | rs10858889 | 0,0000036 | 0,16 | Artery - Tibial |
| ENSG00000070961.15 | ATP2B1 | chr12 89513389 T C b38 | rs10858890 | 0,0000036 | 0,16 | Artery - Tibial |
| ENSG00000070961.15 | ATP2B1 | chr12 89516073 T C b38 | rs11105309 | 0,0000036 | 0,16 | Artery - Tibial |
| ENSG00000070961.15 | ATP2B1 | chr12 89519642 C T b38 | rs1054807 | 0,0000036 | 0,16 | Artery - Tibial |
| ENSG00000070961.15 | ATP2B1 | chr12 89520484 T C b38 | rs10858894 | 0,0000036 | 0,16 | Artery - Tibial |
| ENSG00000070961.15 | ATP2B1 | chr12 89529599 A T b38 | rs11105313 | 0,0000036 | 0,16 | Artery - Tibial |
| ENSG00000070961.15 | ATP2B1 | chr12 90056289 A G b38 | rs12227543 | 0,0000036 | -0,15 | Esophagus - Mucosa |
| ENSG00000070961.15 | ATP2B1 | chr12 89524900 C A b38 | rs11105310 | 0,0000037 | 0,16 | Artery - Tibial |
| ENSG00000070961.15 | ATP2B1 | chr12 89525791 C T b38 | rs10777184 | 0,0000037 | 0,16 | Artery - Tibial |
| ENSG00000070961.15 | ATP2B1 | chr12 89526407 C T b38 | rs10858896 | 0,0000037 | 0,16 | Artery - Tibial |
| ENSG00000070961.15 | ATP2B1 | chr12 89527500 G A b38 | rs10777186 | 0,0000037 | 0,16 | Artery - Tibial |
| ENSG00000070961.15 | ATP2B1 | chr12 90052157 T C b38 | rs4842710 | 0,0000037 | -0,15 | Esophagus - Mucosa |
| ENSG00000070961.15 | ATP2B1 | chr12 90057274 A G b38 | rs10858947 | 0,0000037 | -0,15 | Esophagus - Mucosa |
| ENSG00000070961.15 | ATP2B1 | chr12 90068211 G A b38 | rs7139279 | 0,0000037 | -0,15 | Esophagus - Mucosa |
| ENSG00000070961.15 | ATP2B1 | chr12 90077187 C T b38 | rs7488228 | 0,0000037 | -0,15 | Esophagus - Mucosa |
| ENSG00000070961.15 | ATP2B1 | chr12 90083730 C T b38 | rs4842505 | 0,0000037 | -0,15 | Esophagus - Mucosa |
| ENSG00000070961.15 | ATP2B1 | chr12 90083900 C T b38 | rs7306280 | 0,0000037 | -0,15 | Esophagus - Mucosa |
| ENSG00000070961.15 | ATP2B1 | chr12 89996947 A G b38 | rs7953742 | 0,0000038 | -0,15 | Esophagus - Mucosa |
| ENSG00000070961.15 | ATP2B1 | chr12 89996827 CA C b38 | rs112780917 | 0,0000039 | -0,15 | Esophagus - Mucosa |
| ENSG00000070961.15 | ATP2B1 | chr12 90007154 C CA b38 | rs11396475 | 0,0000039 | -0,15 | Esophagus - Mucosa |
| ENSG00000070961.15 | ATP2B1 | chr12 90018978 A C b38 | rs4842705 | 0,0000039 | -0,15 | Esophagus - Mucosa |
| ENSG00000070961.15 | ATP2B1 | chr12 89531304 C A b38 | rs10858900 | 0,0000041 | 0,16 | Artery - Tibial |
| ENSG00000070961.15 | ATP2B1 | chr12 89536622 A G b38 | rs4842661 | 0,0000047 | 0,17 | Artery - Tibial |
| ENSG00000070961.15 | ATP2B1 | chr12_89539933_C_T_b38 | rs3825330 | 0,000005 | 0,16 | Artery - Tibial |
| ENSG00000070961.15 | ATP2B1 | chr12_89541610_C_T_b38 | rs10858909 | 0,000005 | 0,16 | Artery - Tibial |
| ENSG00000070961.15 | ATP2B1 | chr12_89544813_G_A_b38 | rs11105325 | 0,000005 | 0,16 | Artery - Tibial |
| ENSG00000070961.15 | ATP2B1 | chr12_90050964_A_G_b38 | rs10777222 | 0,000005 | -0,15 | Esophagus - Mucosa |
| ENSG00000070961.15 | ATP2B1 | chr12_89540154_G_A_b38 | rs3741899 | 0,0000051 | 0,16 | Artery - Tibial |
| ENSG00000070961.15 | ATP2B1 | chr12_90389431_C_T_b38 | rs1228496 | 0,0000052 | 0,49 | Brain - Substantia nigra |
| ENSG00000070961.15 | ATP2B1 | chr12_89528334_A_G_b38 | rs10858898 | 0,0000052 | 0,16 | Artery - Tibial |
| ENSG00000070961.15 | ATP2B1 | chr12_89544253_G_A_b38 | rs4842664 | 0,0000052 | 0,16 | Artery - Tibial |
| ENSG00000070961.15 | ATP2B1 | chr12_90510721_G_T_b38 | rs12578110 | 0,0000055 | 0,46 | Brain - Substantia nigra |
| ENSG00000070961.15 | ATP2B1 | chr12_89532881_G_A_b38 | rs11105318 | 0,0000056 | 0,16 | Artery - Tibial |
| ENSG00000070961.15 | ATP2B1 | chr12_89931928_T_C_b38 | rs7970120 | 0,0000063 | -0,21 | Artery - Tibial |
| ENSG00000070961.15 | ATP2B1 | chr12_89432335_C_G_b38 | rs11105286 | 0,0000064 | 0,15 | Artery - Tibial |
| ENSG00000070961.15 | ATP2B1 | chr12_90386667_C_T_b38 | rs6538234 | 0,0000083 | 0,51 | Brain - Substantia nigra |
| ENSG00000070961.15 | ATP2B1 | chr12_90388244_T_A_b38 | rs7979392 | 0,0000083 | 0,51 | Brain - Substantia nigra |
| ENSG00000070961.15 | ATP2B1 | chr12_90390285_G_C_b38 | rs2043213 | 0,0000083 | 0,51 | Brain - Substantia nigra |
| ENSG00000070961.15 | ATP2B1 | chr12_90398938_C_T_b38 | rs7955801 | 0,0000083 | 0,51 | Brain - Substantia nigra |
| ENSG00000070961.15 | ATP2B1 | chr12_90063091_G_A_b38 | rs6538220 | 0,0000084 | -0,15 | Esophagus - Mucosa |
| ENSG00000070961.15 | ATP2B1 | chr12_90516738_A_G_b38 | rs1347015 | 0,0000088 | 0,47 | Brain - Substantia nigra |
| ENSG00000070961.15 | ATP2B1 | chr12_90523189_C_T_b38 | rs1579534 | 0,0000088 | 0,47 | Brain - Substantia nigra |
| ENSG00000070961.15 | ATP2B1 | chr12_90510105_C_T_b38 | rs10858992 | 0,0000088 | 0,46 | Brain - Substantia nigra |
| ENSG00000070961.15 | ATP2B1 | chr12_90519613_A_T_b38 | rs61655253 | 0,0000088 | 0,46 | Brain - Substantia nigra |
| ENSG00000070961.15 | ATP2B1 | chr12_90528232_A_G_b38 | rs4274255 | 0,0000088 | 0,46 | Brain - Substantia nigra |
| ENSG00000070961.15 | ATP2B1 | chr12_90528837_C_T_b38 | rs11105663 | 0,0000088 | 0,46 | Brain - Substantia nigra |
| ENSG00000070961.15 | ATP2B1 | chr12_90532822_TTCTTAAGGACAA_T_b38 | rs6144807 | 0,0000088 | 0,46 | Brain - Substantia nigra |
| ENSG00000070961.15 | ATP2B1 | chr12_90533124_TA_T_b38 | rs36032096 | 0,0000088 | 0,46 | Brain - Substantia nigra |
| ENSG00000070961.15 | ATP2B1 | chr12_90535560_T_C_b38 | rs12580833 | 0,0000088 | 0,46 | Brain - Substantia nigra |
| ENSG00000070961.15 | ATP2B1 | chr12_90543285_C_T_b38 | rs11105668 | 0,0000088 | 0,46 | Brain - Substantia nigra |
| ENSG00000070961.15 | ATP2B1 | chr12_89780365_G_T_b38 | rs116990534 | 0,0000092 | 0,35 | Artery - Tibial |
| ENSG00000070961.15 | ATP2B1 | chr12_89719404_G_A_b38 | rs4842679 | 0,0000094 | -0,15 | Artery - Tibial |
| ENSG00000070961.15 | ATP2B1 | chr12_89504370_G_A_b38 | rs138808993 | 0,0000095 | 0,65 | Brain - Anterior cingulate cortex (BA24) |
| ENSG00000070961.15 | ATP2B1 | chr12_89538657_G_A_b38 | rs143285018 | 0,0000095 | 0,65 | Brain - Anterior cingulate cortex (BA24) |
| ENSG00000070961.15 | ATP2B1 | chr12_90129715_G_A_b38 | rs59313167 | 0,0000097 | -0,15 | Esophagus - Mucosa |
| ENSG00000070961.15 | ATP2B1 | chr12_90134908_C_T_b38 | rs7970711 | 0,0000097 | -0,15 | Esophagus - Mucosa |
| ENSG00000070961.15 | ATP2B1 | chr12_90136962_G_A_b38 | rs7979050 | 0,0000097 | -0,15 | Esophagus - Mucosa |
| ENSG00000070961.15 | ATP2B1 | chr12_89959608_C_T_b38 | rs11105474 | 0,00001 | -0,14 | Esophagus - Mucosa |
| ENSG00000070961.15 | ATP2B1 | chr12_89622750_A_G_b38 | rs147471906 | 0,000011 | 0,59 | Brain - Anterior cingulate cortex (BA24) |
| ENSG00000070961.15 | ATP2B1 | chr12_89643621_G_A_b38 | rs149731397 | 0,000011 | 0,59 | Brain - Anterior cingulate cortex (BA24) |
| ENSG00000070961.15 | ATP2B1 | chr12_90394122_TTTC_T_b38 | rs10543504 | 0,000011 | 0,52 | Brain - Substantia nigra |
| ENSG00000070961.15 | ATP2B1 | chr12_90385245_G_A_b38 | rs1594565 | 0,000011 | 0,5 | Brain - Substantia nigra |
| ENSG00000070961.15 | ATP2B1 | chr12_90088859_T_C_b38 | rs11105501 | 0,000011 | -0,15 | Esophagus - Mucosa |
| ENSG00000070961.15 | ATP2B1 | chr12_90089663_G_A_b38 | rs11105503 | 0,000011 | -0,15 | Esophagus - Mucosa |
| ENSG00000070961.15 | ATP2B1 | chr12_90089938_C_T_b38 | rs11105504 | 0,000011 | -0,15 | Esophagus - Mucosa |
| ENSG00000070961.15 | ATP2B1 | chr12_89926004_G_A_b38 | rs111342256 | 0,000012 | 0,65 | Artery - Tibial |
| ENSG00000070961.15 | ATP2B1 | chr12_90195090_A_G_b38 | rs10858967 | 0,000012 | -0,34 | Cells - EBV-transformed lymphocytes |
| ENSG00000070961.15 | ATP2B1 | chr12_90407150_G_A_b38 | rs7969733 | 0,000013 | 0,34 | Brain - Spinal cord (cervical c-1) |
| ENSG00000070961.15 | ATP2B1 | chr12_90411424_C_G_b38 | rs10777244 | 0,000013 | 0,34 | Brain - Spinal cord (cervical c-1) |
| ENSG00000070961.15 | ATP2B1 | chr12_90411690_G_A_b38 | rs7958205 | 0,000013 | 0,34 | Brain - Spinal cord (cervical c-1) |
| ENSG00000070961.15 | ATP2B1 | chr12_90411780_G_A_b38 | rs7958308 | 0,000013 | 0,34 | Brain - Spinal cord (cervical c-1) |
| ENSG00000070961.15 | ATP2B1 | chr12_90414020_G_T_b38 | rs7966452 | 0,000013 | 0,34 | Brain - Spinal cord (cervical c-1) |
| ENSG00000070961.15 | ATP2B1 | chr12_90414588_CT_C_b38 | rs35829324 | 0,000013 | 0,34 | Brain - Spinal cord (cervical c-1) |
| ENSG00000070961.15 | ATP2B1 | chr12_90416671_A_G_b38 | rs2897862 | 0,000013 | 0,34 | Brain - Spinal cord (cervical c-1) |
| ENSG00000070961.15 | ATP2B1 | chr12_90416672_A_T_b38 | rs2408160 | 0,000013 | 0,34 | Brain - Spinal cord (cervical c-1) |
| ENSG00000070961.15 | ATP2B1 | chr12_90422109_C_T_b38 | rs7315821 | 0,000013 | 0,34 | Brain - Spinal cord (cervical c-1) |
| ENSG00000070961.15 | ATP2B1 | chr12_90425535_T_A_b38 | rs6538235 | 0,000013 | 0,34 | Brain - Spinal cord (cervical c-1) |
| ENSG00000070961.15 | ATP2B1 | chr12_90427858_G_A_b38 | rs10777245 | 0,000013 | 0,34 | Brain - Spinal cord (cervical c-1) |
| ENSG00000070961.15 | ATP2B1 | chr12_90430064_C_G_b38 | rs6538236 | 0,000013 | 0,34 | Brain - Spinal cord (cervical c-1) |
| ENSG00000070961.15 | ATP2B1 | chr12_90436231_T_C_b38 | rs1835768 | 0,000013 | 0,34 | Brain - Spinal cord (cervical c-1) |
| ENSG00000070961.15 | ATP2B1 | chr12_90436298_G_A_b38 | rs1835767 | 0,000013 | 0,34 | Brain - Spinal cord (cervical c-1) |
| ENSG00000070961.15 | ATP2B1 | chr12_90439201_G_A_b38 | rs34234695 | 0,000013 | 0,34 | Brain - Spinal cord (cervical c-1) |
| ENSG00000070961.15 | ATP2B1 | chr12_90439855_C_T_b38 | rs11105617 | 0,000013 | 0,34 | Brain - Spinal cord (cervical c-1) |
| ENSG00000070961.15 | ATP2B1 | chr12_90443686_G_A_b38 | rs66955893 | 0,000013 | 0,34 | Brain - Spinal cord (cervical c-1) |
| ENSG00000070961.15 | ATP2B1 | chr12_90451488_G_A_b38 | rs11105622 | 0,000013 | 0,34 | Brain - Spinal cord (cervical c-1) |
| ENSG00000070961.15 | ATP2B1 | chr12_90452503_C_T_b38 | rs7960740 | 0,000013 | 0,34 | Brain - Spinal cord (cervical c-1) |
| ENSG00000070961.15 | ATP2B1 | chr12_90457546_A_T_b38 | rs11105623 | 0,000013 | 0,34 | Brain - Spinal cord (cervical c-1) |
| ENSG00000070961.15 | ATP2B1 | chr12_90468635_G_A_b38 | rs7954409 | 0,000013 | 0,34 | Brain - Spinal cord (cervical c-1) |
| ENSG00000070961.15 | ATP2B1 | chr12_90474639_G_A_b38 | rs1864782 | 0,000013 | 0,34 | Brain - Spinal cord (cervical c-1) |
| ENSG00000070961.15 | ATP2B1 | chr12_90476135_C_T_b38 | rs7298940 | 0,000013 | 0,34 | Brain - Spinal cord (cervical c-1) |
| ENSG00000070961.15 | ATP2B1 | chr12_90478226_G_A_b38 | rs12818545 | 0,000013 | 0,34 | Brain - Spinal cord (cervical c-1) |
| ENSG00000070961.15 | ATP2B1 | chr12_90482472_AAC_A_b38 | rs67668093 | 0,000013 | 0,34 | Brain - Spinal cord (cervical c-1) |
| ENSG00000070961.15 | ATP2B1 | chr12_90484858_C_T_b38 | rs7311599 | 0,000013 | 0,34 | Brain - Spinal cord (cervical c-1) |
| ENSG00000070961.15 | ATP2B1 | chr12_90559065_T_A_b38 | rs2520561 | 0,000013 | 0,31 | Brain - Spinal cord (cervical c-1) |
| ENSG00000070961.15 | ATP2B1 | chr12_90391772_A_C_b38 | rs7976895 | 0,000014 | 0,48 | Brain - Substantia nigra |
| ENSG00000070961.15 | ATP2B1 | chr12_90392860_A_C_b38 | rs2116497 | 0,000014 | 0,48 | Brain - Substantia nigra |
| ENSG00000070961.15 | ATP2B1 | chr12_90393455_G_A_b38 | rs12580873 | 0,000014 | 0,48 | Brain - Substantia nigra |
| ENSG00000070961.15 | ATP2B1 | chr12_90552916_G_A_b38 | rs2520567 | 0,000014 | 0,44 | Brain - Substantia nigra |
| ENSG00000070961.15 | ATP2B1 | chr12_90001825_T_C_b38 | rs17192565 | 0,000014 | 0,43 | Artery - Tibial |
| ENSG00000070961.15 | ATP2B1 | chr12_90459042_T_A_b38 | rs7132660 | 0,000014 | 0,34 | Brain - Spinal cord (cervical c-1) |
| ENSG00000070961.15 | ATP2B1 | chr12_90106317_C CAT_b38 | rs148143712 | 0,000015 | -0,15 | Esophagus - Mucosa |
| ENSG00000070961.15 | ATP2B1 | chr12_90112140_G_A_b38 | rs7966278 | 0,000015 | -0,15 | Esophagus - Mucosa |
| ENSG00000070961.15 | ATP2B1 | chr12_89556543_A_C_b38 | rs7302816 | 0,000016 | 0,19 | Artery - Aorta |
| ENSG00000070961.15 | ATP2B1 | chr12_89422110_G_A_b38 | rs80277599 | 0,000017 | 1,4 | Brain - Hypothalamus |
| ENSG00000070961.15 | ATP2B1 | chr12_89428758_A_G_b38 | rs112370308 | 0,000017 | 1,4 | Brain - Hypothalamus |
| ENSG00000070961.15 | ATP2B1 | chr12_89441431_T_C_b38 | rs113025676 | 0,000017 | 1,4 | Brain - Hypothalamus |
| ENSG00000070961.15 | ATP2B1 | chr12_89457067_C_T_b38 | rs61435009 | 0,000017 | 1,4 | Brain - Hypothalamus |
| ENSG00000070961.15 | ATP2B1 | chr12_89471149_A_G_b38 | rs74754458 | 0,000017 | 1,4 | Brain - Hypothalamus |
| ENSG00000070961.15 | ATP2B1 | chr12_89478671_T_C_b38 | rs60772618 | 0,000017 | 1,4 | Brain - Hypothalamus |
| ENSG00000070961.15 | ATP2B1 | chr12_89484136_C_G_b38 | rs74543833 | 0,000017 | 1,4 | Brain - Hypothalamus |
| ENSG00000070961.15 | ATP2B1 | chr12_89487042_C_G_b38 | rs58117070 | 0,000017 | 1,4 | Brain - Hypothalamus |
| ENSG00000070961.15 | ATP2B1 | chr12_89488169_C_T_b38 | rs185300284 | 0,000017 | 1,4 | Brain - Hypothalamus |
| ENSG00000070961.15 | ATP2B1 | chr12_89488792_C_T_b38 | rs76672752 | 0,000017 | 1,4 | Brain - Hypothalamus |
| ENSG00000070961.15 | ATP2B1 | chr12_89490053_G_T_b38 | rs78332000 | 0,000017 | 1,4 | Brain - Hypothalamus |
| ENSG00000070961.15 | ATP2B1 | chr12_90581088_T TA_b38 | rs143922775 | 0,000017 | 1,4 | Brain - Hypothalamus |
| ENSG00000070961.15 | ATP2B1 | chr12_90583461_C_T_b38 | rs17018043 | 0,000017 | 1,4 | Brain - Hypothalamus |
| ENSG00000070961.15 | ATP2B1 | chr12_90586839_A_G_b38 | rs73358180 | 0,000017 | 1,4 | Brain - Hypothalamus |
| ENSG00000070961.15 | ATP2B1 | chr12_90589431_T_C_b38 | rs73358183 | 0,000017 | 1,4 | Brain - Hypothalamus |
| ENSG00000070961.15 | ATP2B1 | chr12_90593192_C_T_b38 | rs73358194 | 0,000017 | 1,4 | Brain - Hypothalamus |
| ENSG00000070961.15 | ATP2B1 | chr12_90594592_G_A_b38 | rs2106528 | 0,000017 | 1,4 | Brain - Hypothalamus |
| ENSG00000070961.15 | ATP2B1 | chr12_90599502_C_T_b38 | rs76134788 | 0,000017 | 1,4 | Brain - Hypothalamus |
| ENSG00000070961.15 | ATP2B1 | chr12_90602009_G_C_b38 | rs73360109 | 0,000017 | 1,4 | Brain - Hypothalamus |
| ENSG00000070961.15 | ATP2B1 | chr12_90602975_T_C_b38 | rs73360112 | 0,000017 | 1,4 | Brain - Hypothalamus |
| ENSG00000070961.15 | ATP2B1 | chr12_90603192_A_G_b38 | rs17018088 | 0,000017 | 1,4 | Brain - Hypothalamus |
| ENSG00000070961.15 | ATP2B1 | chr12_90606864_CAG_C_b38 | rs34639687 | 0,000017 | 1,4 | Brain - Hypothalamus |
| ENSG00000070961.15 | ATP2B1 | chr12_90608792_A_G_b38 | rs7306918 | 0,000017 | 1,4 | Brain - Hypothalamus |
| ENSG00000070961.15 | ATP2B1 | chr12_90609936_C_T_b38 | rs73360125 | 0,000017 | 1,4 | Brain - Hypothalamus |
| ENSG00000070961.15 | ATP2B1 | chr12_90614576_G_T_b38 | rs73360133 | 0,000017 | 1,4 | Brain - Hypothalamus |
| ENSG00000070961.15 | ATP2B1 | chr12_90626419_G_T_b38 | rs11829105 | 0,000017 | 1,4 | Brain - Hypothalamus |
| ENSG00000070961.15 | ATP2B1 | chr12_90626488_A_T_b38 | rs11833692 | 0,000017 | 1,4 | Brain - Hypothalamus |
| ENSG00000070961.15 | ATP2B1 | chr12_90628581_A_G_b38 | rs73360170 | 0,000017 | 1,4 | Brain - Hypothalamus |
| ENSG00000070961.15 | ATP2B1 | chr12_90646115_A_G_b38 | rs11830515 | 0,000017 | 1,4 | Brain - Hypothalamus |
| ENSG00000070961.15 | ATP2B1 | chr12_90650906_T_C_b38 | rs11105723 | 0,000017 | 1,4 | Brain - Hypothalamus |
| ENSG00000070961.15 | ATP2B1 | chr12_90661217_T_C_b38 | rs11105730 | 0,000017 | 1,4 | Brain - Hypothalamus |
| ENSG00000070961.15 | ATP2B1 | chr12_89525853_ACC_A_b38 | rs67346689 | 0,000017 | 0,16 | Artery - Tibial |
| ENSG00000070961.15 | ATP2B1 | chr12_90637101_TTG_T_b38 | rs1491580271 | 0,000018 | 1,4 | Brain - Hypothalamus |
| ENSG00000070961.15 | ATP2B1 | chr12_90136354_G_A_b38 | rs143689254 | 0,000018 | 0,68 | Brain - Anterior cingulate cortex (BA24) |
| ENSG00000070961.15 | ATP2B1 | chr12_90241276_A_G_b38 | rs73196463 | 0,000018 | 0,62 | Artery - Tibial |
| ENSG00000070961.15 | ATP2B1 | chr12_90554733_G_A_b38 | rs2520565 | 0,000018 | 0,44 | Brain - Substantia nigra |
| ENSG00000070961.15 | ATP2B1 | chr12_90556726_A_G_b38 | rs2520563 | 0,000018 | 0,44 | Brain - Substantia nigra |
| ENSG00000070961.15 | ATP2B1 | chr12_90557663_C_T_b38 | rs2520562 | 0,000018 | 0,44 | Brain - Substantia nigra |
| ENSG00000070961.15 | ATP2B1 | chr12_90558072_T_C_b38 | rs2723917 | 0,000018 | 0,44 | Brain - Substantia nigra |
| ENSG00000070961.15 | ATP2B1 | chr12_89569279_G_A_b38 | rs372643302 | 0,000018 | 0,17 | Artery - Tibial |
| ENSG00000070961.15 | ATP2B1 | chr12_89950918_A_G_b38 | rs7294671 | 0,000018 | -0,13 | Esophagus - Mucosa |
| ENSG00000070961.15 | ATP2B1 | chr12_89956338_C_G_b38 | rs11105473 | 0,000018 | -0,14 | Esophagus - Mucosa |
| ENSG00000070961.15 | ATP2B1 | chr12_89957460_T_G_b38 | rs7132526 | 0,000018 | -0,14 | Esophagus - Mucosa |
| ENSG00000070961.15 | ATP2B1 | chr12_89704243_G_A_b38 | rs61926784 | 0,000019 | 0,19 | Brain - Frontal Cortex (BA9) |
| ENSG00000070961.15 | ATP2B1 | chr12_90326873_G_T_b38 | rs825954 | 0,000023 | -0,15 | Esophagus - Muscularis |
| ENSG00000070961.15 | ATP2B1 | chr12_90329398_C_T_b38 | rs10858974 | 0,000024 | -0,13 | Thyroid |
| ENSG00000070961.15 | ATP2B1 | chr12_90299326_G_A_b38 | rs11105563 | 0,000024 | -0,35 | Cells - EBV-transformed lymphocytes |
| ENSG00000070961.15 | ATP2B1 | chr12_89876539_G_A_b38 | rs74851539 | 0,000025 | 0,41 | Artery - Tibial |
| ENSG00000070961.15 | ATP2B1 | chr12_89940862_G_A_b38 | rs12368259 | 0,000027 | 0,22 | Esophagus - Mucosa |
| ENSG00000070961.15 | ATP2B1 | chr12_90299488_G_C_b38 | rs10506984 | 0,000027 | -0,34 | Cells - EBV-transformed lymphocytes |
| ENSG00000070961.15 | ATP2B1 | chr12_90303890_G_C_b38 | rs2056699 | 0,000027 | -0,34 | Cells - EBV-transformed lymphocytes |
| ENSG00000070961.15 | ATP2B1 | chr12_90306554_G_A_b38 | rs1587440 | 0,000027 | -0,34 | Cells - EBV-transformed lymphocytes |
| ENSG00000070961.15 | ATP2B1 | chr12_90311197_C_T_b38 | rs10858971 | 0,000027 | -0,34 | Cells - EBV-transformed lymphocytes |
| ENSG00000070961.15 | ATP2B1 | chr12_90631370_A_T_b38 | rs11105706 | 0,000028 | -0,28 | Colon - Transverse |
| ENSG00000070961.15 | ATP2B1 | chr12_90631458_A_G_b38 | rs11105707 | 0,000028 | -0,28 | Colon - Transverse |
| ENSG00000070961.15 | ATP2B1 | chr12_90632378_G_A_b38 | rs11105709 | 0,000028 | -0,28 | Colon - Transverse |
| ENSG00000070961.15 | ATP2B1 | chr12_90636015_C_A_b38 | rs11105712 | 0,000028 | -0,28 | Colon - Transverse |
| ENSG00000070961.15 | ATP2B1 | chr12_90638508_T_A_b38 | rs112784216 | 0,000028 | -0,28 | Colon - Transverse |
| ENSG00000070961.15 | ATP2B1 | chr12_90451210_G_A_b38 | rs1148676 | 0,000029 | 0,3 | Brain - Spinal cord (cervical c-1) |
| ENSG00000070961.15 | ATP2B1 | chr12_89973104_A_G_b38 | rs10777215 | 0,000029 | -0,13 | Esophagus - Mucosa |
| ENSG00000070961.15 | ATP2B1 | chr12_89999628_A_ATTC_b38 | rs10671258 | 0,000029 | -0,13 | Esophagus - Mucosa |
| ENSG00000070961.15 | ATP2B1 | chr12_90000940_A_G_b38 | rs4406875 | 0,000029 | -0,13 | Esophagus - Mucosa |
| ENSG00000070961.15 | ATP2B1 | chr12_90005097_T_A_b38 | rs4240746 | 0,000029 | -0,13 | Esophagus - Mucosa |
| ENSG00000070961.15 | ATP2B1 | chr12_89559134_C_T_b38 | rs112417857 | 0,000031 | 0,54 | Artery - Tibial |
| ENSG00000070961.15 | ATP2B1 | chr12_89394010_T_C_b38 | rs11105273 | 0,000031 | 0,18 | Artery - Tibial |
| ENSG00000070961.15 | ATP2B1 | chr12_89975574_A_G_b38 | rs7303123 | 0,000031 | -0,13 | Esophagus - Mucosa |
| ENSG00000070961.15 | ATP2B1 | chr12_89976239_C_T_b38 | rs10777216 | 0,000031 | -0,13 | Esophagus - Mucosa |
| ENSG00000070961.15 | ATP2B1 | chr12_89957885_A_G_b38 | rs4842700 | 0,000032 | -0,18 | Artery - Tibial |
| ENSG00000070961.15 | ATP2B1 | chr12_90287401_C_T_b38 | rs11105559 | 0,000032 | -0,34 | Cells - EBV-transformed lymphocytes |
| ENSG00000070961.15 | ATP2B1 | chr12_90295570_G_A_b38 | rs2165035 | 0,000032 | -0,34 | Cells - EBV-transformed lymphocytes |
| ENSG00000070961.15 | ATP2B1 | chr12_90273379_C_A_b38 | rs12320693 | 0,000033 | -0,37 | Cells - EBV-transformed lymphocytes |
| ENSG00000070961.15 | ATP2B1 | chr12_89995520_A_C_b38 | rs6538214 | 0,000034 | -0,13 | Esophagus - Mucosa |
| ENSG00000070961.15 | ATP2B1 | chr12_90007782_T_TATAAA_b38 | rs57291885 | 0,000034 | -0,13 | Esophagus - Mucosa |
| ENSG00000070961.15 | ATP2B1 | chr12_90008924_A_C_b38 | rs4842504 | 0,000034 | -0,13 | Esophagus - Mucosa |
| ENSG00000070961.15 | ATP2B1 | chr12_90018772_T_C_b38 | rs10858942 | 0,000034 | -0,13 | Esophagus - Mucosa |
| ENSG00000070961.15 | ATP2B1 | chr12_90061403_T_C_b38 | rs7303035 | 0,000037 | -0,13 | Esophagus - Mucosa |
| ENSG00000070961.15 | ATP2B1 | chr12_89042639_G_A_b38 | rs76671707 | 0,000037 | -0,57 | Lung |
| ENSG00000070961.15 | ATP2B1 | chr12_90266564_G_A_b38 | rs1347846 | 0,000039 | -0,33 | Cells - EBV-transformed lymphocytes |
| ENSG00000070961.15 | ATP2B1 | chr12_90259816_T_C_b38 | rs4381424 | 0,000039 | -0,34 | Cells - EBV-transformed lymphocytes |
| ENSG00000070961.15 | ATP2B1 | chr12_90303378_AT_A_b38 | rs141511768 | 0,000039 | -0,36 | Cells - EBV-transformed lymphocytes |
| ENSG00000070961.15 | ATP2B1 | chr12_90275487_CAA_C_b38 | rs3042884 | 0,000041 | -0,35 | Cells - EBV-transformed lymphocytes |
| ENSG00000070961.15 | ATP2B1 | chr12_90328476_T_C_b38 | rs1405527 | 0,000042 | -0,13 | Thyroid |
| ENSG00000070961.15 | ATP2B1 | chr12_90330109_G_A_b38 | rs12818395 | 0,000042 | -0,13 | Thyroid |
| ENSG00000070961.15 | ATP2B1 | chr12_90150155_C_T_b38 | rs7135166 | 0,000042 | -0,14 | Esophagus - Mucosa |
| ENSG00000070961.15 | ATP2B1 | chr12_89709017_C_A_b38 | rs12369944 | 0,000044 | 0,18 | Brain - Frontal Cortex (BA9) |
| ENSG00000070961.15 | ATP2B1 | chr12_89966286_A_G_b38 | rs10745520 | 0,000044 | -0,13 | Esophagus - Mucosa |
| ENSG00000070961.15 | ATP2B1 | chr12 89702020 T A b38 | rs11105380 | 0,000045 | -0,14 | Artery - Tibial |
| ENSG00000070961.15 | ATP2B1 | chr12 90153750 C G b38 | rs2118803 | 0,000047 | -0,14 | Esophagus - Mucosa |
| ENSG00000070961.15 | ATP2B1 | chr12 90568565 A G b38 | rs35520023 | 0,000048 | 0,13 | Adipose - Subcutaneous |
| ENSG00000070961.15 | ATP2B1 | chr12 90629207 G A b38 | rs11105704 | 0,000048 | -0,27 | Colon - Transverse |
| ENSG00000070961.15 | ATP2B1 | chr12 89666809 G A b38 | rs17249754 | 0,000051 | 0,19 | Artery - Aorta |
| ENSG00000070961.15 | ATP2B1 | chr12 89668599 G A b38 | rs6538195 | 0,000051 | 0,19 | Artery - Aorta |
| ENSG00000070961.15 | ATP2B1 | chr12 89678299 G A b38 | rs73437358 | 0,000051 | 0,19 | Artery - Aorta |
| ENSG00000070961.15 | ATP2B1 | chr12 89913265 C T b38 | rs7972820 | 0,000053 | -0,13 | Esophagus - Mucosa |
| ENSG00000070961.15 | ATP2B1 | chr12 89784106 A T b38 | rs113133867 | 0,000054 | 0,55 | Artery - Tibial |
| ENSG00000070961.15 | ATP2B1 | chr12 89632685 G A b38 | rs11105352 | 0,000054 | 0,19 | Artery - Aorta |
| ENSG00000070961.15 | ATP2B1 | chr12 89632686 C A b38 | rs11105353 | 0,000054 | 0,19 | Artery - Aorta |
| ENSG00000070961.15 | ATP2B1 | chr12 89632746 A G b38 | rs11105354 | 0,000054 | 0,19 | Artery - Aorta |
| ENSG00000070961.15 | ATP2B1 | chr12 89656726 A G b38 | rs12579302 | 0,000054 | 0,19 | Artery - Aorta |
| ENSG00000070961.15 | ATP2B1 | chr12 89665065 G A b38 | rs111478946 | 0,000054 | 0,19 | Artery - Aorta |
| ENSG00000070961.15 | ATP2B1 | chr12 89675499 T G b38 | rs11105364 | 0,000054 | 0,19 | Artery - Aorta |
| ENSG00000070961.15 | ATP2B1 | chr12 90158457 G GT b38 | rs79468373 | 0,000054 | -0,14 | Esophagus - Mucosa |
| ENSG00000070961.15 | ATP2B1 | chr12 89909790 G C b38 | rs7957657 | 0,000057 | -0,13 | Esophagus - Mucosa |
| ENSG00000070961.15 | ATP2B1 | chr12 90328476 T C b38 | rs1405527 | 0,000057 | -0,15 | Esophagus - Muscularis |
| ENSG00000070961.15 | ATP2B1 | chr12 90329398 C T b38 | rs10858974 | 0,000057 | -0,15 | Esophagus - Muscularis |
| ENSG00000070961.15 | ATP2B1 | chr12 90330109 G A b38 | rs12818395 | 0,000057 | -0,15 | Esophagus - Muscularis |
| ENSG00000070961.15 | ATP2B1 | chr12 90026116 GT G b38 | rs11324040 | 0,000058 | -0,13 | Esophagus - Mucosa |
| ENSG00000070961.15 | ATP2B1 | chr12 89629273 T C b38 | rs116858620 | 0,00006 | 0,43 | Lung |
| ENSG00000070961.15 | ATP2B1 | chr12 90035251 A G b38 | rs7311631 | 0,00006 | -0,13 | Esophagus - Mucosa |
| ENSG00000070961.15 | ATP2B1 | chr12 89879521 G A b38 | rs2553100 | 0,00006 | -0,13 | Esophagus - Mucosa |
| ENSG00000070961.15 | ATP2B1 | chr12 89944741 G A b38 | rs4842698 | 0,000062 | -0,18 | Artery - Tibial |
| ENSG00000070961.15 | ATP2B1 | chr12 89680664 G C b38 | rs11105368 | 0,000063 | 0,19 | Artery - Aorta |
| ENSG00000070961.15 | ATP2B1 | chr12 90577787 T C b38 | rs11105685 | 0,000063 | 0,13 | Adipose - Subcutaneous |
| ENSG00000070961.15 | ATP2B1 | chr12 89681466 C T b38 | rs4842675 | 0,000065 | 0,19 | Artery - Aorta |
| ENSG00000070961.15 | ATP2B1 | chr12 89619312 T C b38 | rs2681492 | 0,000066 | 0,19 | Artery - Aorta |
| ENSG00000070961.15 | ATP2B1 | chr12 89615182 A G b38 | rs2681472 | 0,000066 | 0,19 | Artery - Aorta |
| ENSG00000070961.15 | ATP2B1 | chr12 89956222 G A b38 | rs10777213 | 0,000066 | -0,13 | Esophagus - Mucosa |
| ENSG00000070961.15 | ATP2B1 | chr12 89958659 C T b38 | rs7306228 | 0,000066 | -0,13 | Esophagus - Mucosa |
| ENSG00000070961.15 | ATP2B1 | chr12 90039790 A G b38 | rs6538218 | 0,000066 | -0,13 | Esophagus - Mucosa |
| ENSG00000070961.15 | ATP2B1 | chr12 89912752 T C b38 | rs7135197 | 0,000067 | -0,13 | Esophagus - Mucosa |
| ENSG00000070961.15 | ATP2B1 | chr12 89915156 G A b38 | rs7959788 | 0,000067 | -0,13 | Esophagus - Mucosa |
| ENSG00000070961.15 | ATP2B1 | chr12 90152680 A G b38 | rs10745525 | 0,000068 | -0,18 | Artery - Tibial |
| ENSG00000070961.15 | ATP2B1 | chr12 89837533 A G b38 | rs2722224 | 0,000079 | -0,13 | Esophagus - Mucosa |
| ENSG00000070961.15 | ATP2B1 | chr12 89696964 C T b38 | rs11105378 | 0,00008 | 0,19 | Artery - Aorta |
| ENSG00000070961.15 | ATP2B1 | chr12 89695013 G A b38 | rs10858917 | 0,00008 | -0,17 | Artery - Aorta |
| ENSG00000070961.15 | ATP2B1 | chr12 90050278 C T b38 | rs55846104 | 0,000081 | -0,19 | Esophagus - Mucosa |
| ENSG00000070961.15 | ATP2B1 | chr12 90147932 C T b38 | rs4608145 | 0,000083 | -0,13 | Lung |
| ENSG00000070961.15 | ATP2B1 | chr12 90087601 T C b38 | rs7316395 | 0,000087 | -0,13 | Lung |
| ENSG00000070961.15 | ATP2B1 | chr12 90087981 A G b38 | rs7300611 | 0,000087 | -0,13 | Lung |
| ENSG00000070961.15 | ATP2B1 | chr12 90089145 A G b38 | rs10735278 | 0,000087 | -0,13 | Lung |
| ENSG00000070961.15 | ATP2B1 | chr12 90089249 T C b38 | rs10735279 | 0,000087 | -0,13 | Lung |
| ENSG00000070961.15 | ATP2B1 | chr12 90089939 A G b38 | rs10858950 | 0,000087 | -0,13 | Lung |
| ENSG00000070961.15 | ATP2B1 | chr12 90090107 T C b38 | rs11105505 | 0,000087 | -0,13 | Lung |
| ENSG00000070961.15 | ATP2B1 | chr12 89966684 C T b38 | rs4584633 | 0,00009 | -0,17 | Artery - Tibial |
| ENSG00000070961.15 | ATP2B1 | chr12 89921101 C G b38 | rs4842693 | 0,000093 | -0,13 | Esophagus - Mucosa |
| ENSG00000070961.15 | ATP2B1 | chr12 90133970 C T b38 | rs11105516 | 0,000094 | -0,13 | Lung |
| ENSG00000070961.15 | ATP2B1 | chr12 90022580 A C b38 | rs10777219 | 0,000094 | -0,14 | Artery - Tibial |
| ENSG00000070961.15 | ATP2B1 | chr12 90024345 G A b38 | rs10858943 | 0,000094 | -0,14 | Artery - Tibial |
| ENSG00000070961.15 | ATP2B1 | chr12 90157130 AT A b38 | rs34734819 | 0,000099 | -0,13 | Esophagus - Mucosa |
| ENSG00000070961.15 | ATP2B1 | chr12 89694803 T A b38 | rs11105375 | 0,0001 | 0,18 | Artery - Aorta |
| ENSG00000070961.15 | ATP2B1 | chr12 89697090 A G b38 | rs12230074 | 0,0001 | 0,18 | Artery - Aorta |
| ENSG00000070961.15 | ATP2B1 | chr12 90148406 A C b38 | rs1438984 | 0,0001 | -0,13 | Lung |
| ENSG00000070961.15 | ATP2B1 | chr12 90148426 G A b38 | rs1438983 | 0,0001 | -0,13 | Lung |
| ENSG00000070961.15 | ATP2B1 | chr12 90042305 C T b38 | rs7975953 | 0,0001 | -0,14 | Artery - Tibial |
| ENSG00000070961.15 | ATP2B1 | chr12 90042794 T C b38 | rs7963304 | 0,0001 | -0,14 | Artery - Tibial |
| ENSG00000070961.15 | ATP2B1 | chr12 90110385 A C b38 | rs10858955 | 0,00011 | -0,13 | Lung |
| ENSG00000070961.15 | ATP2B1 | chr12 90111572 G A b38 | rs10745523 | 0,00011 | -0,13 | Lung |
| ENSG00000070961.15 | ATP2B1 | chr12 90114532 G T b38 | rs10777224 | 0,00011 | -0,13 | Lung |
| ENSG00000070961.15 | ATP2B1 | chr12 90120301 C G b38 | rs10858956 | 0,00011 | -0,13 | Lung |
| ENSG00000070961.15 | ATP2B1 | chr12 89336143 A C b38 | rs10161308 | 0,00012 | 0,22 | Artery - Tibial |
| ENSG00000070961.15 | ATP2B1 | chr12 89660842 T C b38 | rs73437338 | 0,00012 | 0,19 | Artery - Aorta |
| ENSG00000070961.15 | ATP2B1 | chr12 89591515 AAAGT A b38 | rs10580742 | 0,00012 | 0,19 | Artery - Aorta |
| ENSG00000070961.15 | ATP2B1 | chr12 89677042 CAAAG C b38 | rs143087380 | 0,00012 | -0,11 | Thyroid |
| ENSG00000070961.15 | ATP2B1 | chr12 90099780 G A b38 | rs7486390 | 0,00012 | -0,13 | Lung |
| ENSG00000070961.15 | ATP2B1 | chr12 90100728 G C b38 | rs7303398 | 0,00012 | -0,13 | Lung |
| ENSG00000070961.15 | ATP2B1 | chr12 89694991 G A b38 | rs11105376 | 0,00013 | 0,19 | Artery - Aorta |
| ENSG00000070961.15 | ATP2B1 | chr12 90117803 G C b38 | rs7486630 | 0,00013 | -0,13 | Lung |
| ENSG00000070961.15 | ATP2B1 | chr12 90119438 T A b38 | rs7139353 | 0,00013 | -0,13 | Lung |
| ENSG00000070961.15 | ATP2B1 | chr12 90121102 T C b38 | rs10777226 | 0,00013 | -0,13 | Lung |
| ENSG00000070961.15 | ATP2B1 | chr12 90122191 G C b38 | rs6538227 | 0,00013 | -0,13 | Lung |
| ENSG00000070961.15 | ATP2B1 | chr12 90123327 G T b38 | rs10858957 | 0,00013 | -0,13 | Lung |
| ENSG00000070961.15 | ATP2B1 | chr12 90123338 T C b38 | rs10858958 | 0,00013 | -0,13 | Lung |
| ENSG00000070961.15 | ATP2B1 | chr12 90124075 G A b38 | rs7315166 | 0,00013 | -0,13 | Lung |
| ENSG00000070961.15 | ATP2B1 | chr12 90124724 G A b38 | rs4842713 | 0,00013 | -0,13 | Lung |
| ENSG00000070961.15 | ATP2B1 | chr12 90126369 C T b38 | rs7138547 | 0,00013 | -0,13 | Lung |
| ENSG00000070961.15 | ATP2B1 | chr12 90127423 A G b38 | rs10777227 | 0,00013 | -0,13 | Lung |
| ENSG00000070961.15 | ATP2B1 | chr12 90127443 A G b38 | rs10777228 | 0,00013 | -0,13 | Lung |
| ENSG00000070961.15 | ATP2B1 | chr12 90128789 G A b38 | rs12303180 | 0,00013 | -0,13 | Lung |
| ENSG00000070961.15 | ATP2B1 | chr12 90129235 T C b38 | rs4842714 | 0,00013 | -0,13 | Lung |
| ENSG00000070961.15 | ATP2B1 | chr12 90130357 GCTC G b38 | rs35647353 | 0,00013 | -0,13 | Lung |
| ENSG00000070961.15 | ATP2B1 | chr12 90132074 G T b38 | rs10858961 | 0,00013 | -0,13 | Lung |
| ENSG00000070961.15 | ATP2B1 | chr12 90133198 A AG b38 | rs79553876 | 0,00013 | -0,13 | Lung |
| ENSG00000070961.15 | ATP2B1 | chr12 90134620 G C b38 | rs6538228 | 0,00013 | -0,13 | Lung |
| ENSG00000070961.15 | ATP2B1 | chr12 90137321 T C b38 | rs1595599 | 0,00013 | -0,13 | Lung |
| ENSG00000070961.15 | ATP2B1 | chr12 90137388 G A b38 | rs1595598 | 0,00013 | -0,13 | Lung |
| ENSG00000070961.15 | ATP2B1 | chr12 90137720 T G b38 | rs7304226 | 0,00013 | -0,13 | Lung |
| ENSG00000070961.15 | ATP2B1 | chr12 90139086 C T b38 | rs7295818 | 0,00013 | -0,13 | Lung |
| ENSG00000070961.15 | ATP2B1 | chr12 90141861 G T b38 | rs1438986 | 0,00013 | -0,13 | Lung |
| ENSG00000070961.15 | ATP2B1 | chr12 90142076 T G b38 | rs1438985 | 0,00013 | -0,13 | Lung |
| ENSG00000070961.15 | ATP2B1 | chr12 90142537 C A b38 | rs1470031 | 0,00013 | -0,13 | Lung |
| ENSG00000070961.15 | ATP2B1 | chr12 90142786 G A b38 | rs1470030 | 0,00013 | -0,13 | Lung |
| ENSG00000070961.15 | ATP2B1 | chr12 90144066 A C b38 | rs10745524 | 0,00013 | -0,13 | Lung |
| ENSG00000070961.15 | ATP2B1 | chr12 90144234 A G b38 | rs1470029 | 0,00013 | -0,13 | Lung |
| ENSG00000070961.15 | ATP2B1 | chr12 90144899 A T b38 | rs4842715 | 0,00013 | -0,13 | Lung |
| ENSG00000070961.15 | ATP2B1 | chr12 90145400 C A b38 | rs4842716 | 0,00013 | -0,13 | Lung |
| ENSG00000070961.15 | ATP2B1 | chr12 90145401 C T b38 | rs4842717 | 0,00013 | -0,13 | Lung |
| ENSG00000070961.15 | ATP2B1 | chr12 90145987 C A b38 | rs1344962 | 0,00013 | -0,13 | Lung |
| ENSG00000070961.15 | ATP2B1 | chr12 90146644 T C b38 | rs11105517 | 0,00013 | -0,13 | Lung |
| ENSG00000070961.15 | ATP2B1 | chr12 90147279 C G b38 | rs1344961 | 0,00013 | -0,13 | Lung |
| ENSG00000070961.15 | ATP2B1 | chr12 90147385 C G b38 | rs2218085 | 0,00013 | -0,13 | Lung |
| ENSG00000070961.15 | ATP2B1 | chr12 90147644 A C b38 | rs2196783 | 0,00013 | -0,13 | Lung |
| ENSG00000070961.15 | ATP2B1 | chr12 90002703 T C b38 | rs7964216 | 0,00013 | -0,14 | Artery - Tibial |
| ENSG00000070961.15 | ATP2B1 | chr12 90004485 T C b38 | rs10777218 | 0,00013 | -0,14 | Artery - Tibial |
| ENSG00000070961.15 | ATP2B1 | chr12 90005574 C T b38 | rs11105483 | 0,00013 | -0,14 | Artery - Tibial |
| ENSG00000070961.15 | ATP2B1 | chr12 90015717 C T b38 | rs61926987 | 0,00013 | -0,14 | Artery - Tibial |
| ENSG00000070961.15 | ATP2B1 | chr12 90159905 G A b38 | rs6538230 | 0,00014 | -0,16 | Artery - Tibial |
| ENSG00000070961.15 | ATP2B1 | chr12 89547597 G A b38 | rs4842665 | 0,00015 | 0,12 | Artery - Tibial |
| ENSG00000070961.15 | ATP2B1 | chr12 89865713 T C b38 | rs2722213 | 0,00015 | -0,12 | Esophagus - Mucosa |
| ENSG00000070961.15 | ATP2B1 | chr12 89866278 G A b38 | rs2553097 | 0,00015 | -0,12 | Esophagus - Mucosa |
| ENSG00000070961.15 | ATP2B1 | chr12 89875126 G A b38 | rs2553104 | 0,00015 | -0,12 | Esophagus - Mucosa |
| ENSG00000070961.15 | ATP2B1 | chr12 90071866 G A b38 | rs6538222 | 0,00015 | -0,13 | Lung |
| ENSG00000070961.15 | ATP2B1 | chr12 89851601 T C b38 | rs2553101 | 0,00015 | -0,13 | Esophagus - Mucosa |
| ENSG00000070961.15 | ATP2B1 | chr12 89637618 C T b38 | rs10858914 | 0,00017 | -0,12 | Artery - Tibial |
| ENSG00000070961.15 | ATP2B1 | chr12 90010069 T C b38 | rs4393376 | 0,00017 | -0,15 | Artery - Tibial |
| ENSG00000070961.15 | ATP2B1 | chr12 90016872 A C b38 | rs6538217 | 0,00017 | -0,15 | Artery - Tibial |
| ENSG00000070961.15 | ATP2B1 | chr12 89904501 A C b38 | rs7965284 | 0,00017 | -0,18 | Artery - Tibial |
| ENSG00000070961.15 | ATP2B1 | chr12 90128372 C T b38 | rs10858959 | 0,00019 | -0,13 | Artery - Tibial |
| ENSG00000070961.15 | ATP2B1 | chr12 90129926 G A b38 | rs10858960 | 0,00019 | -0,13 | Artery - Tibial |
| ENSG00000070961.15 | ATP2B1 | chr12 89943230 A C b38 | rs4842696 | 0,0002 | -0,14 | Esophagus - Mucosa |
| ENSG00000070961.15 | ATP2B1 | chr12 89345678 C G b38 | rs11105262 | 0,00022 | 0,21 | Artery - Tibial |
| ENSG00000070961.15 | ATP2B1 | chr12 89861997 C A b38 | rs1946327 | 0,00023 | -0,12 | Esophagus - Mucosa |
| ENSG00000070961.15 | ATP2B1 | chr12 90003409 T C b38 | rs4453292 | 0,00023 | -0,15 | Artery - Tibial |
| ENSG00000070961.15 | ATP2B1 | chr12 90162353 T C b38 | rs2731241 | 0,00023 | -0,16 | Artery - Tibial |
| ENSG00000070961.15 | ATP2B1 | chr12 90164205 A G b38 | rs1438980 | 0,00023 | -0,16 | Artery - Tibial |
| ENSG00000070961.15 | ATP2B1 | chr12 89434120 G A b38 | rs118130015 | 0,00027 | 0,33 | Artery - Tibial |
| ENSG00000070961.15 | ATP2B1 | chr12 90173201 C T b38 | rs1347851 | 0,00027 | -0,15 | Artery - Tibial |
| ENSG00000070961.15 | ATP2B1 | chr12 89623959 G T b38 | rs2070759 | 0,00028 | -0,11 | Artery - Tibial |
| ENSG00000070961.15 | ATP2B1 | chr12 89646107 G A b38 | rs939329 | 0,00028 | -0,11 | Artery - Tibial |
| ENSG00000070961.15 | ATP2B1 | chr12 90149730 G A b38 | rs10777230 | 0,00028 | -0,12 | Artery - Tibial |
| ENSG00000070961.15 | ATP2B1 | chr12 89857319 C T b38 | rs12371308 | 0,00028 | -0,13 | Artery - Tibial |
| ENSG00000070961.15 | ATP2B1 | chr12 89857983 T C b38 | rs11105439 | 0,00028 | -0,13 | Artery - Tibial |

**Table S4:**
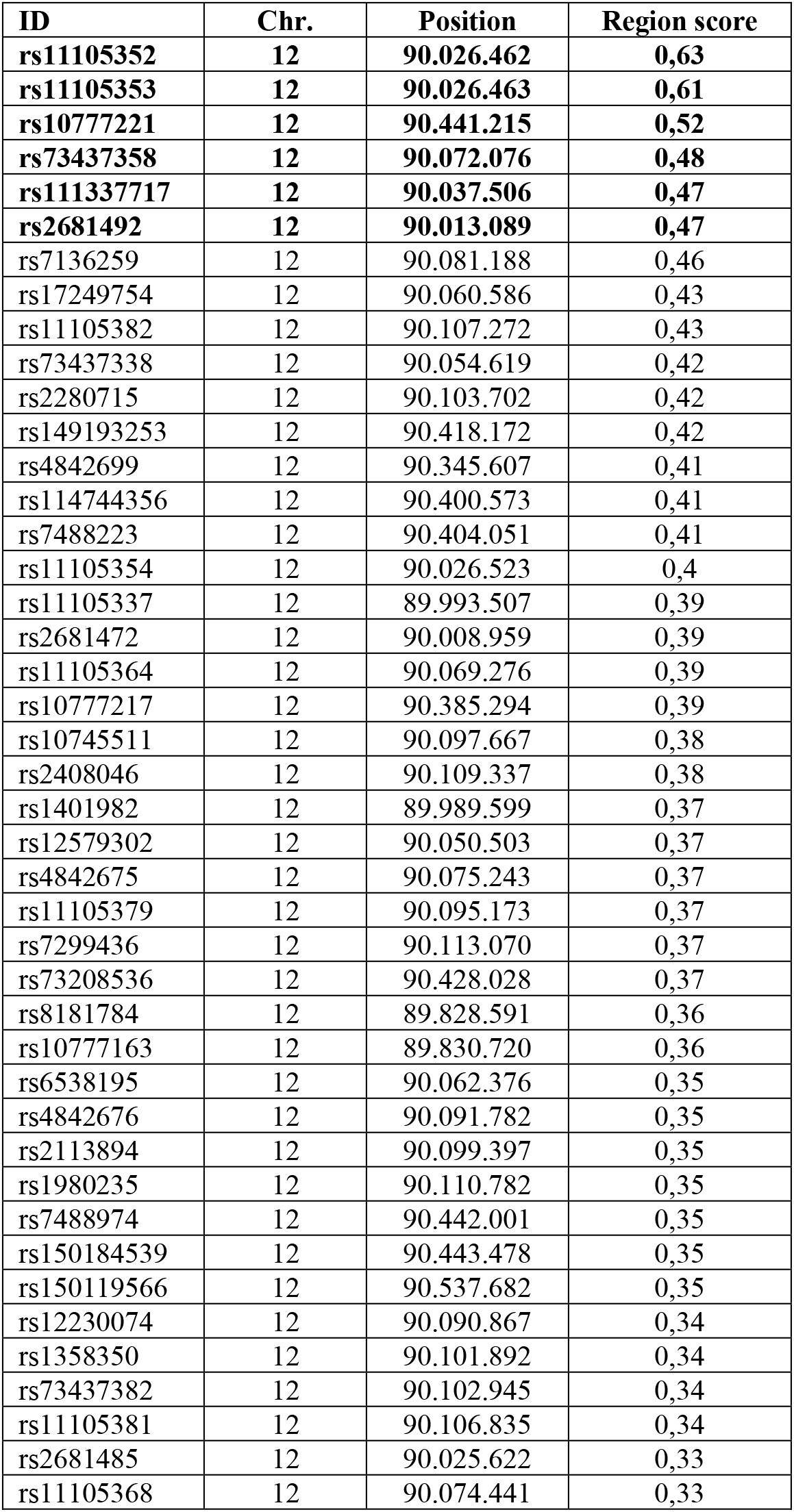
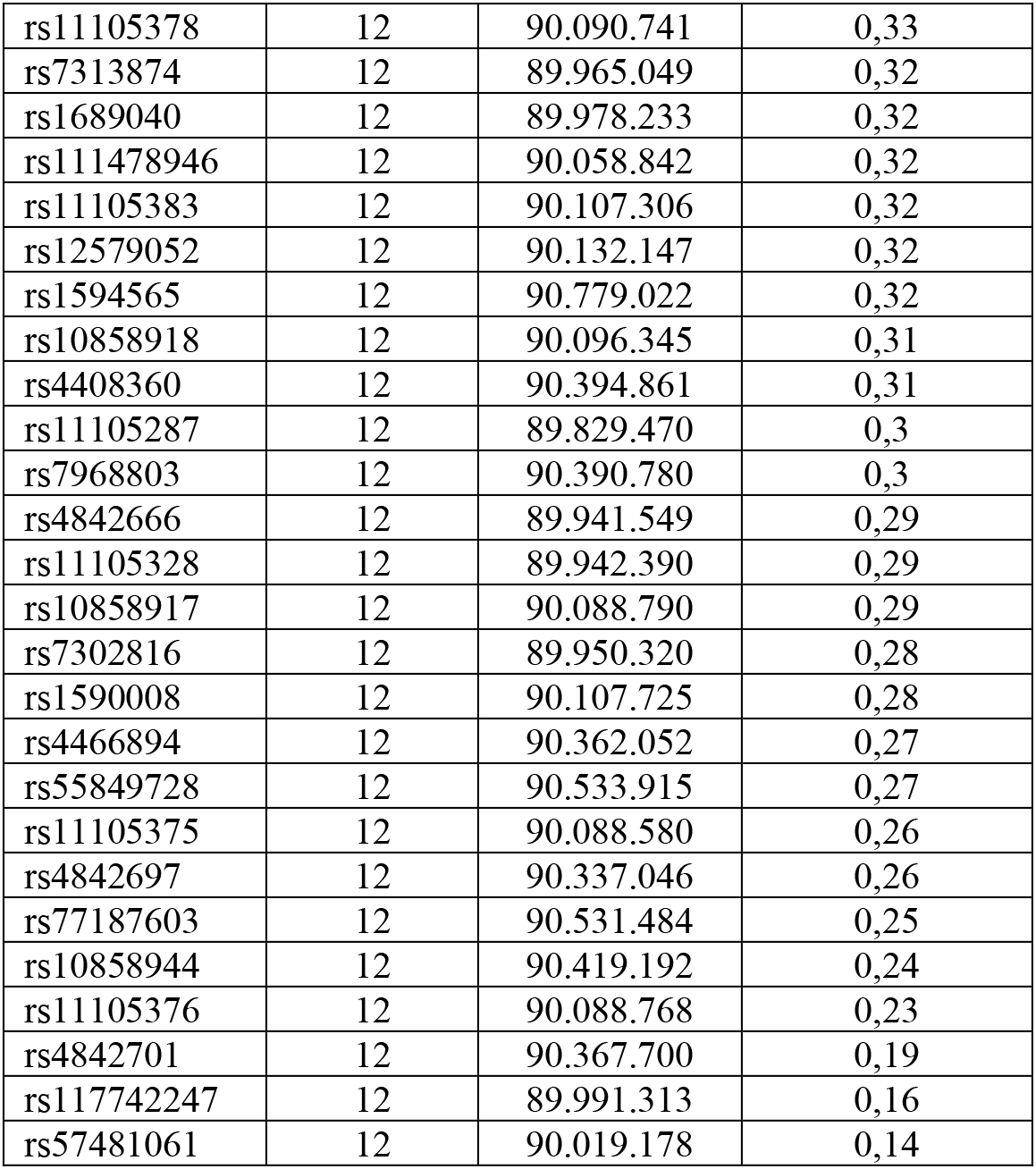
eQTLs SNPs for ATP2B1 gene by using “Genotype-Tissue Expression” GTex database (P<1×10^-6^). N.77 SNPs acting as eQTLs for ATP2B1 gene annotated with prediction functional scores calculated by the “Genome-Wide Annotation of Variants” GWAVA tool are shown. N.8 SNPs are not reported in GWAVA are thus excluded. The ID, the chromosome, the position and the region score are listed. In bold, the top six are selected (rs11105352, rs11105353, rs10777221, rs73437358, rs111337717 and rs2681492).

| ID | Chr. | Position | Region score |
| --- | --- | --- | --- |
| **rs11105352** | **12** | **90.026.462** | **0,63** |
| **rs11105353** | **12** | **90.026.463** | **0,61** |
| **rs10777221** | **12** | **90.441.215** | **0,52** |
| **rs73437358** | **12** | **90.072.076** | **0,48** |
| **rs111337717** | **12** | **90.037.506** | **0,47** |
| **rs2681492** | **12** | **90.013.089** | **0,47** |
| rs7136259 | 12 | 90.081.188 | 0,46 |
| rs17249754 | 12 | 90.060.586 | 0,43 |
| rs11105382 | 12 | 90.107.272 | 0,43 |
| rs73437338 | 12 | 90.054.619 | 0,42 |
| rs2280715 | 12 | 90.103.702 | 0,42 |
| rs149193253 | 12 | 90.418.172 | 0,42 |
| rs4842699 | 12 | 90.345.607 | 0,41 |
| rs114744356 | 12 | 90.400.573 | 0,41 |
| rs7488223 | 12 | 90.404.051 | 0,41 |
| rs11105354 | 12 | 90.026.523 | 0,4 |
| rs11105337 | 12 | 89.993.507 | 0,39 |
| rs2681472 | 12 | 90.008.959 | 0,39 |
| rs11105364 | 12 | 90.069.276 | 0,39 |
| rs10777217 | 12 | 90.385.294 | 0,39 |
| rs10745511 | 12 | 90.097.667 | 0,38 |
| rs2408046 | 12 | 90.109.337 | 0,38 |
| rs1401982 | 12 | 89.989.599 | 0,37 |
| rs12579302 | 12 | 90.050.503 | 0,37 |
| rs4842675 | 12 | 90.075.243 | 0,37 |
| rs11105379 | 12 | 90.095.173 | 0,37 |
| rs7299436 | 12 | 90.113.070 | 0,37 |
| rs73208536 | 12 | 90.428.028 | 0,37 |
| rs8181784 | 12 | 89.828.591 | 0,36 |
| rs10777163 | 12 | 89.830.720 | 0,36 |
| rs6538195 | 12 | 90.062.376 | 0,35 |
| rs4842676 | 12 | 90.091.782 | 0,35 |
| rs2113894 | 12 | 90.099.397 | 0,35 |
| rs1980235 | 12 | 90.110.782 | 0,35 |
| rs7488974 | 12 | 90.442.001 | 0,35 |
| rs150184539 | 12 | 90.443.478 | 0,35 |
| rs150119566 | 12 | 90.537.682 | 0,35 |
| rs12230074 | 12 | 90.090.867 | 0,34 |
| rs1358350 | 12 | 90.101.892 | 0,34 |
| rs73437382 | 12 | 90.102.945 | 0,34 |
| rs11105381 | 12 | 90.106.835 | 0,34 |
| rs2681485 | 12 | 90.025.622 | 0,33 |
| rs11105368 | 12 | 90.074.441 | 0,33 |
| rs11105378 | 12 | 90.090.741 | 0,33 |
| rs7313874 | 12 | 89.965.049 | 0,32 |
| rs1689040 | 12 | 89.978.233 | 0,32 |
| rs111478946 | 12 | 90.058.842 | 0,32 |
| rs11105383 | 12 | 90.107.306 | 0,32 |
| rs12579052 | 12 | 90.132.147 | 0,32 |
| rs1594565 | 12 | 90.779.022 | 0,32 |
| rs10858918 | 12 | 90.096.345 | 0,31 |
| rs4408360 | 12 | 90.394.861 | 0,31 |
| rs11105287 | 12 | 89.829.470 | 0,3 |
| rs7968803 | 12 | 90.390.780 | 0,3 |
| rs4842666 | 12 | 89.941.549 | 0,29 |
| rs11105328 | 12 | 89.942.390 | 0,29 |
| rs10858917 | 12 | 90.088.790 | 0,29 |
| rs7302816 | 12 | 89.950.320 | 0,28 |
| rs1590008 | 12 | 90.107.725 | 0,28 |
| rs4466894 | 12 | 90.362.052 | 0,27 |
| rs55849728 | 12 | 90.533.915 | 0,27 |
| rs11105375 | 12 | 90.088.580 | 0,26 |
| rs4842697 | 12 | 90.337.046 | 0,26 |
| rs77187603 | 12 | 90.531.484 | 0,25 |
| rs10858944 | 12 | 90.419.192 | 0,24 |
| rs11105376 | 12 | 90.088.768 | 0,23 |
| rs4842701 | 12 | 90.367.700 | 0,19 |
| rs117742247 | 12 | 89.991.313 | 0,16 |
| rs57481061 | 12 | 90.019.178 | 0,14 |

**Table S5:** Differentially expressed proteins in HEK293T-ACE2 upon treatment with 1 μM PI-7 molecule for 24 hours. Proteomic analysis in HEK293T-ACE2 upon treatment with 1 μM PI-7 molecule for 24 hours In the treated cells we found n.17 downregulated and n.66 upregulated proteins. The protein entry, the protein names, the gene names, the log2 fold changes (log2FC), the fold change (FC), the Student’s T-test FDR, the number of pepdides (Razor + unique peptides) and the percentage of the sequence coverage (Unique + razon sequence coverage) are shown.

| Entry | Protein name | Gene name | log2FC | FC | Student's *t*-test FDR | Razor + unique peptides | Unique + razor sequence coverage (%) |
| --- | --- | --- | --- | --- | --- | --- | --- |
| Q6FI81 | Anamorsin | CIAPIN1 | -0,92 | 0,53 | 3,71E-02 | 4 | 24,4 |
| P55145 | Mesencephalic astrocyte-derived neurotrophic factor | MANF | -0,87 | 0,55 | 4,88E-02 | 6 | 33 |
| P10412 | Histone H1.4 | HIST1H1E | -0,73 | 0,60 | 5,44E-04 | 12 | 32,4 |
| P51572 | B-cell receptor-associated protein 31 | BCAP31 | -0,72 | 0,61 | 6,31E-03 | 7 | 26 |
| P62280 | 40S ribosomal protein S11 | RPS11 | -0,70 | 0,61 | 5,76E-03 | 8 | 41,1 |
| P47813 | Eukaryotic translation initiation factor 1A, X-chromosomal | EIF1AX | -0,70 | 0,62 | 3,17E-03 | 6 | 34 |
| P62750 | 60S ribosomal protein L23a | RPL23A | -0,62 | 0,65 | 3,54E-03 | 9 | 37,2 |
| P62979 | Ubiquitin-40S ribosomal protein S27a | RPS27A | -0,62 | 0,65 | 5,82E-05 | 12 | 55,8 |
| P84090 | Enhancer of rudimentary homolog | ERH | -0,58 | 0,67 | 1,32E-02 | 4 | 37,5 |
| P60866 | 40S ribosomal protein S20 | RPS20 | -0,58 | 0,67 | 2,10E-03 | 4 | 28,6 |
| P35613 | Basigin | BSG | -0,57 | 0,67 | 2,79E-03 | 6 | 22,9 |
| Q9H444 | Charged multivesicular body protein 4b | CHMP4B | -0,57 | 0,67 | 5,54E-04 | 6 | 27,7 |
| Q07020 | 60S ribosomal protein L18 | RPL18 | -0,56 | 0,68 | 9,09E-03 | 7 | 35,6 |
| P42677 | 40S ribosomal protein S27 | RPS27 | -0,55 | 0,68 | 2,24E-04 | 5 | 40,5 |
| P25788 | Proteasome subunit alpha type-3 | PSMA3 | -0,52 | 0,70 | 5,44E-04 | 6 | 22,4 |
| P13073 | Cytochrome c oxidase subunit 4 isoform 1, mitochondrial | COX4I1 | -0,50 | 0,71 | 7,07E-03 | 6 | 31,4 |
| Q9BRA2 | Thioredoxin domain-containing protein 17 | TXNDC17 | -0,50 | 0,71 | 3,68E-03 | 4 | 46,3 |
| P38159 | RNA-binding motif protein, X chromosome | RBMX | -0,50 | 0,71 | 2,35E-04 | 10 | 26,1 |
| P48643 | T-complex protein 1 subunit epsilon | CCT5 | 0,50 | 1,41 | 6,28E-04 | 28 | 68,6 |
| P12532 | Creatine kinase U-type, mitochondrial | CKMT1A | 0,50 | 1,41 | 6,28E-04 | 7 | 35,5 |
| P04844 | Dolichyl-diphosphooligosaccharide--protein glycosyltransferase subunit 2 | RPN2 | 0,50 | 1,41 | 1,71E-04 | 16 | 39,9 |
| Q9Y5L0 | Transportin-3 | TNPO3 | 0,50 | 1,41 | 4,25E-02 | 6 | 11,1 |
| P63208 | S-phase kinase-associated protein 1 | SKP1 | 0,50 | 1,41 | 7,76E-03 | 5 | 33,7 |
| P06748 | Nucleophosmin | NPM1 | 0,51 | 1,42 | 3,58E-03 | 11 | 36,4 |
| O15160 | DNA-directed RNA polymerases I and III subunit RPAC1 | POLR1C | 0,52 | 1,43 | 3,37E-03 | 5 | 21,7 |
| Q9Y285 | Phenylalanine--tRNA ligase alpha subunit | FARSA | 0,52 | 1,44 | 1,69E-02 | 6 | 19,5 |
| P23921 | Ribonucleoside-diphosphate reductase large subunit | RRM1 | 0,53 | 1,44 | 8,05E-03 | 8 | 19,4 |
| Q13126 | S-methyl-5-thioadenosine phosphorylase | MTAP | 0,55 | 1,46 | 3,11E-03 | 8 | 48,1 |
| Q9BSJ8 | Extended synaptotagmin-1 | ESYT1 | 0,55 | 1,47 | 1,37E-02 | 5 | 6,4 |
| Q15435 | Protein phosphatase 1 regulatory subunit 7 | PPP1R7 | 0,55 | 1,47 | 4,65E-03 | 6 | 23,9 |
| Q92616 | eIF-2-alpha kinase activator GCN1 | GCN1 | 0,55 | 1,47 | 1,74E-04 | 35 | 21 |
| P24534 | Elongation factor 1-beta | EEF1B2 | 0,56 | 1,47 | 2,75E-03 | 5 | 37,3 |
| P09104 | Gamma-enolase | ENO2 | 0,56 | 1,47 | 2,35E-04 | 13 | 48,6 |
| Q14257 | Reticulocalbin-2 | RCN2 | 0,57 | 1,49 | 6,68E-03 | 4 | 20,8 |
| P62917 | 60S ribosomal protein L8 | RPL8 | 0,58 | 1,50 | 9,79E-03 | 7 | 33,9 |
| Q8IY67 | Ribonucleoprotein PTB-binding 1 | RAVER1 | 0,58 | 1,50 | 2,35E-04 | 5 | 17,3 |
| Q8IZL8 | Proline-, glutamic acid- and leucine-rich protein 1 | PELP1 | 0,59 | 1,51 | 3,75E-03 | 6 | 9,4 |
| P61011 | Signal recognition particle 54 kDa protein | SRP54 | 0,60 | 1,51 | 2,37E-02 | 7 | 21 |
| Q9UBE0 | SUMO-activating enzyme subunit 1 | SAE1 | 0,60 | 1,52 | 4,37E-04 | 11 | 47,1 |
| Q6P1J9 | Parafibromin | CDC73 | 0,60 | 1,52 | 2,70E-02 | 4 | 9,6 |
| Q9ULC4 | Malignant T-cell-amplified sequence 1 | MCTS1 | 0,62 | 1,54 | 3,58E-03 | 7 | 57,5 |
| P47985 | Cytochrome b-c1 complex subunit Rieske, mitochondrial | UQCRFS1 | 0,62 | 1,54 | 3,37E-03 | 4 | 22,6 |
| Q8N3U4 | Cohesin subunit SA-2 | STAG2 | 0,63 | 1,55 | 1,48E-02 | 7 | 8,1 |
| P26038 | Moesin | MSN | 0,63 | 1,55 | 5,64E-03 | 7 | 15,8 |
| P60953 | Cell division control protein 42 homolog | CDC42 | 0,63 | 1,55 | 7,15E-03 | 8 | 48,2 |
| Q15021 | Condensin complex subunit 1 | NCAPD2 | 0,64 | 1,55 | 3,53E-03 | 14 | 16,2 |
| Q96P70 | Importin-9 | IPO9 | 0,66 | 1,58 | 1,03E-02 | 5 | 9,1 |
| P26599 | Polypyrimidine tract-binding protein 1 | PTBP1 | 0,66 | 1,59 | 2,24E-04 | 16 | 52 |
| Q9C0B1 | Alpha-ketoglutarate-dependent dioxygenase FTO | FTO | 0,67 | 1,59 | 6,56E-03 | 7 | 20,4 |
| Q92896 | Golgi apparatus protein 1 | GLG1 | 0,69 | 1,61 | 2,52E-02 | 4 | 4,8 |
| Q96CS3 | FAS-associated factor 2 | FAF2 | 0,69 | 1,62 | 2,35E-04 | 4 | 16,2 |
| P61970 | Nuclear transport factor 2 | NUTF2 | 0,70 | 1,62 | 3,58E-03 | 5 | 69,3 |
| P63010 | AP-2 complex subunit beta | AP2B1 | 0,70 | 1,63 | 2,39E-04 | 10 | 20,4 |
| P04632 | Calpain small subunit 1 | CAPNS1 | 0,71 | 1,63 | 3,81E-02 | 5 | 31 |
| Q9Y6G9 | Cytoplasmic dynein 1 light intermediate chain 1 | DYNC1LI1 | 0,73 | 1,65 | 2,07E-03 | 5 | 18,4 |
| Q9HAV4 | Exportin-5 | XPO5 | 0,73 | 1,65 | 2,21E-03 | 13 | 15,9 |
| O75947 | ATP synthase subunit d, mitochondrial | ATP5PD | 0,73 | 1,66 | 1,71E-03 | 8 | 64 |
| P08708 | 40S ribosomal protein S17 | RPS17 | 0,74 | 1,67 | 9,90E-04 | 8 | 60 |
| P20290 | Transcription factor BTF3 | BTF3 | 0,75 | 1,68 | 1,20E-03 | 6 | 48,1 |
| Q96AC1 | Fermitin family homolog 2 | FERMT2 | 0,75 | 1,68 | 1,48E-02 | 8 | 18,4 |
| Q5JTH9 | RRP12-like protein | RRP12 | 0,79 | 1,72 | 4,43E-02 | 11 | 12,8 |
| Q9H0A0 | RNA cytidine acetyltransferase | NAT10 | 0,79 | 1,73 | 5,44E-04 | 12 | 17,2 |
| Q96T76 | MMS19 nucleotide excision repair protein homolog | MMS19 | 0,80 | 1,75 | 5,44E-04 | 6 | 10,2 |
| P62081 | 40S ribosomal protein S7 | RPS7 | 0,82 | 1,76 | 2,31E-03 | 12 | 55,7 |
| O00429 | Dynamin-1-like protein | DNM1L | 0,82 | 1,77 | 1,42E-03 | 12 | 27,3 |
| P35270 | Sepiapterin reductase | SPR | 0,87 | 1,83 | 1,81E-02 | 4 | 24,5 |
| O00764 | Pyridoxal kinase | PDXK | 0,87 | 1,83 | 1,97E-02 | 4 | 21,2 |
| O15260 | Surfeit locus protein 4 | SURF4 | 0,88 | 1,83 | 1,82E-03 | 4 | 18,2 |
| Q9BXW7 | Haloacid dehalogenase-like hydrolase domain-containing 5 | HDHD5 | 0,92 | 1,89 | 2,99E-03 | 6 | 25,1 |
| P61086 | Ubiquitin-conjugating enzyme E2 K | UBE2K | 0,92 | 1,90 | 3,36E-02 | 4 | 33 |
| P99999 | Cytochrome c | CYCS | 0,97 | 1,95 | 6,59E-04 | 4 | 41 |
| Q14978 | Nucleolar and coiled-body phosphoprotein 1 | NOLC1 | 1,01 | 2,01 | 2,14E-02 | 8 | 15,2 |
| Q99436 | Proteasome subunit beta type-7 | PSMB7 | 1,01 | 2,02 | 7,16E-03 | 6 | 34,3 |
| Q9GZS3 | WD repeat-containing protein 61 | WDR61 | 1,02 | 2,02 | 2,21E-03 | 4 | 26,9 |
| Q9BWD1 | Acetyl-CoA acetyltransferase, cytosolic | ACAT2 | 1,03 | 2,04 | 2,07E-04 | 5 | 35 |
| Q05639 | Elongation factor 1-alpha 2 | EEF1A2 | 1,04 | 2,05 | 8,77E-04 | 8 | 38,2 |
| Q14011 | Cold-inducible RNA-binding protein | CIRBP | 1,06 | 2,09 | 4,55E-04 | 4 | 32 |
| Q16836 | Hydroxyacyl-coenzyme A dehydrogenase, mitochondrial | HADH | 1,07 | 2,09 | 4,26E-04 | 8 | 42,4 |
| Q8N684 | Cleavage and polyadenylation specificity factor subunit 7 | CPSF7 | 1,08 | 2,11 | 2,71E-02 | 4 | 13,4 |
| Q99598 | Translin-associated protein X | TSNAX | 1,11 | 2,16 | 7,56E-03 | 4 | 22,8 |
| Q15428 | Splicing factor 3A subunit 2 | SF3A2 | 1,11 | 2,17 | 4,19E-02 | 7 | 24,4 |
| P25787 | Proteasome subunit alpha type-2 | PSMA2 | 1,12 | 2,17 | 1,71E-04 | 7 | 45,7 |
| P07602 | Prosaposin | PSAP | 1,31 | 2,47 | 2,35E-02 | 4 | 9,2 |
| Q7L1Q6 | Basic leucine zipper and W2 domain-containing protein 1 | BZW1 | 2,10 | 4,28 | 1,76E-03 | 8 | 23,4 |

**Table S6:**
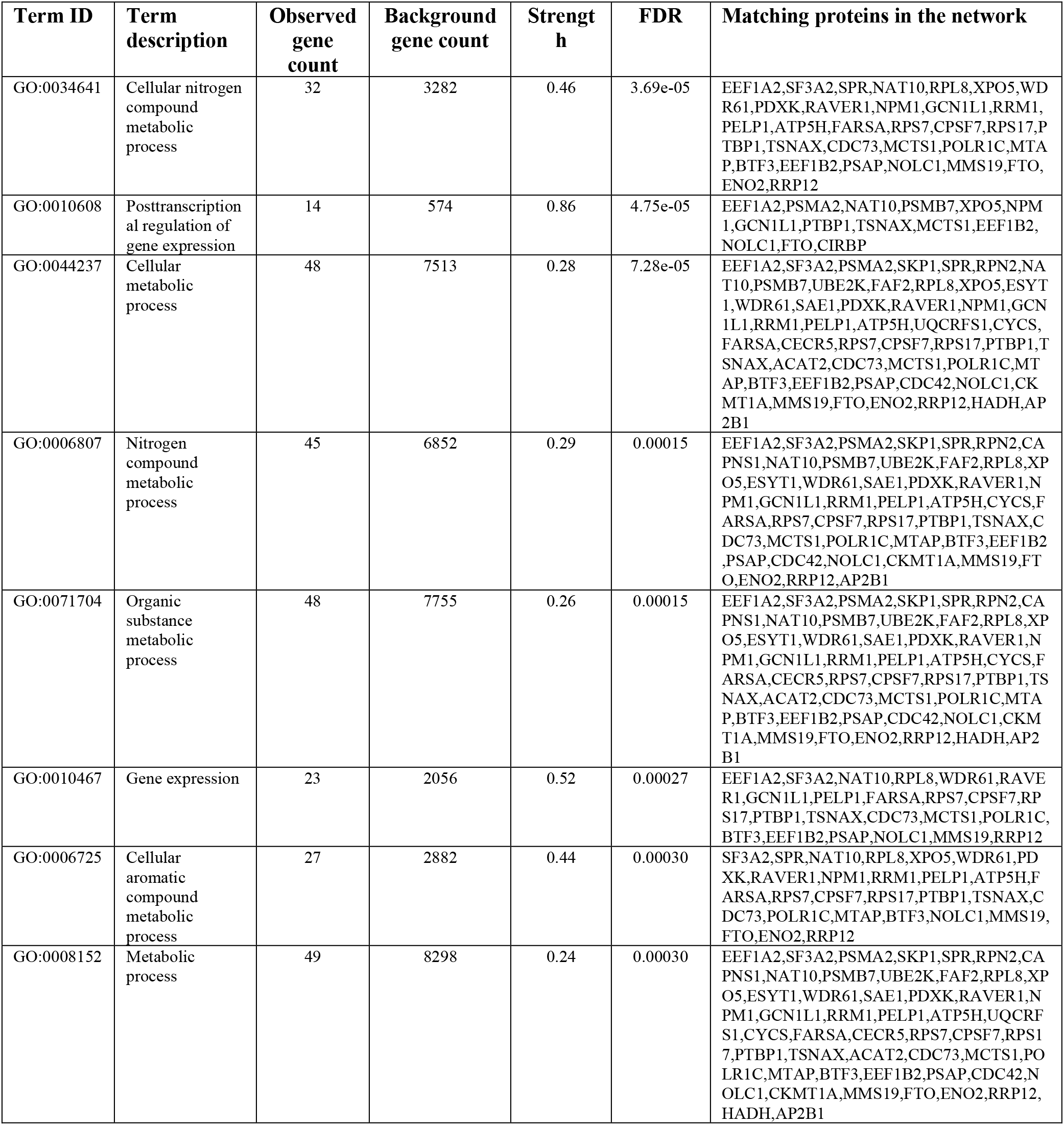
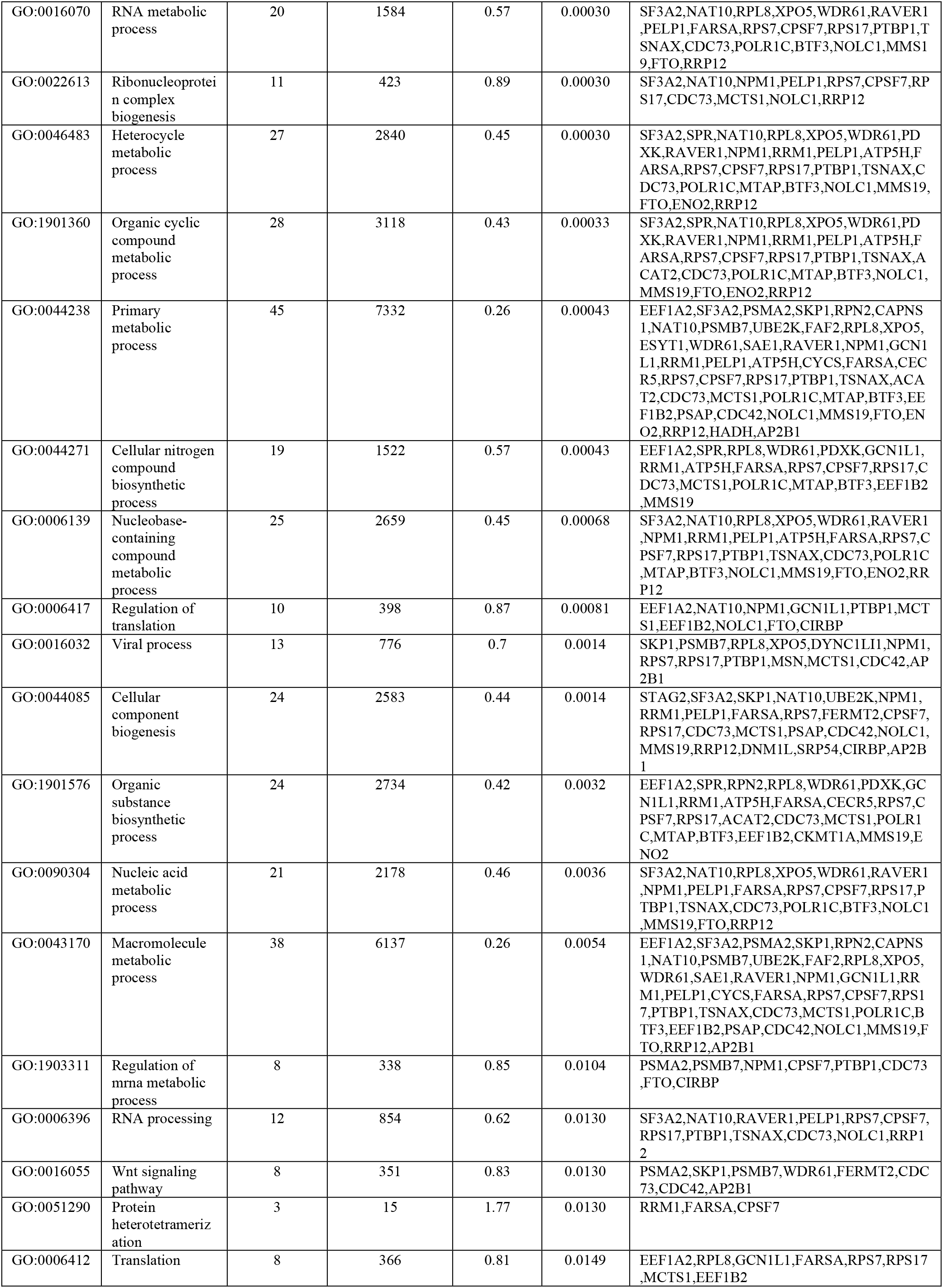
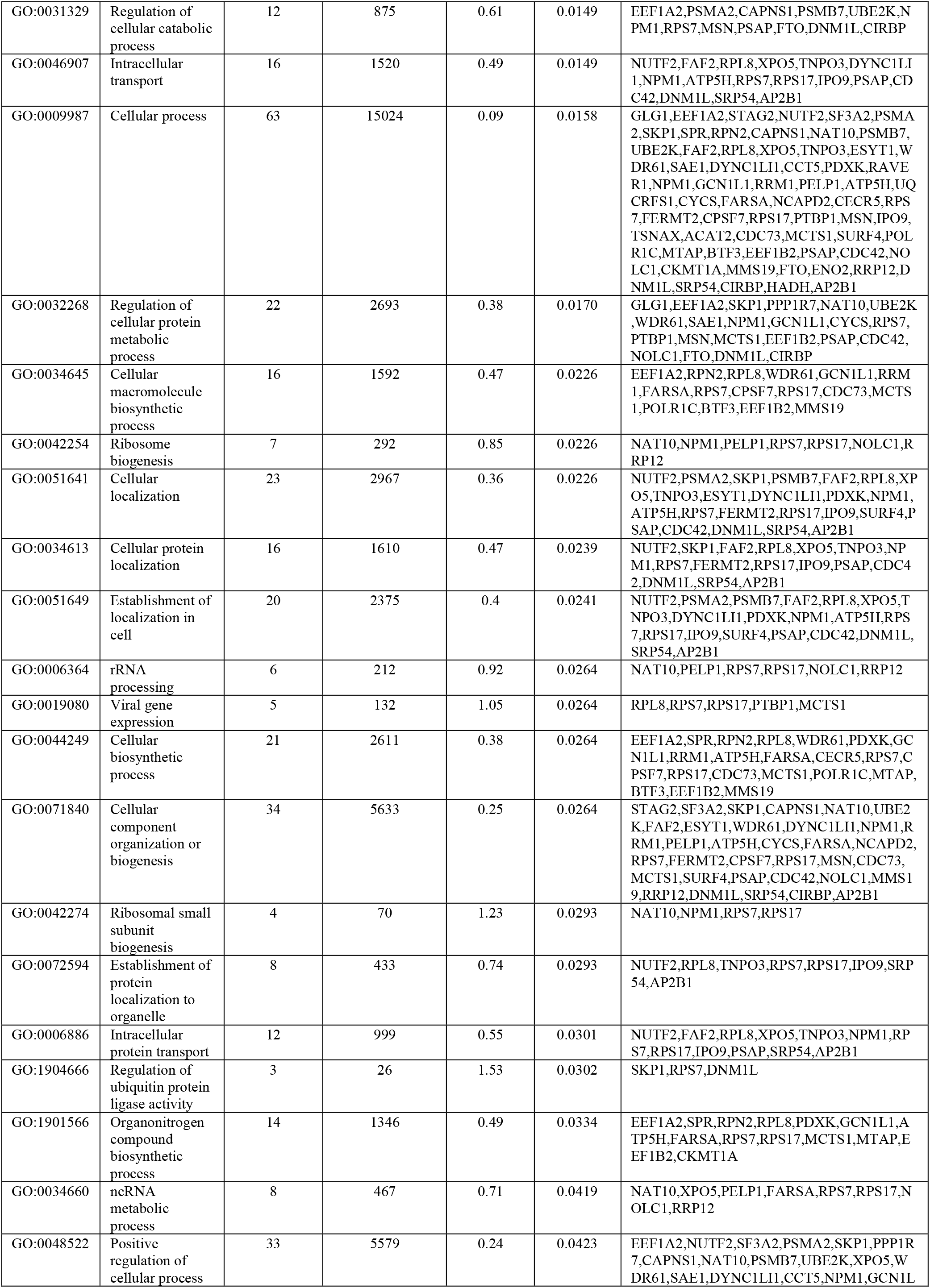
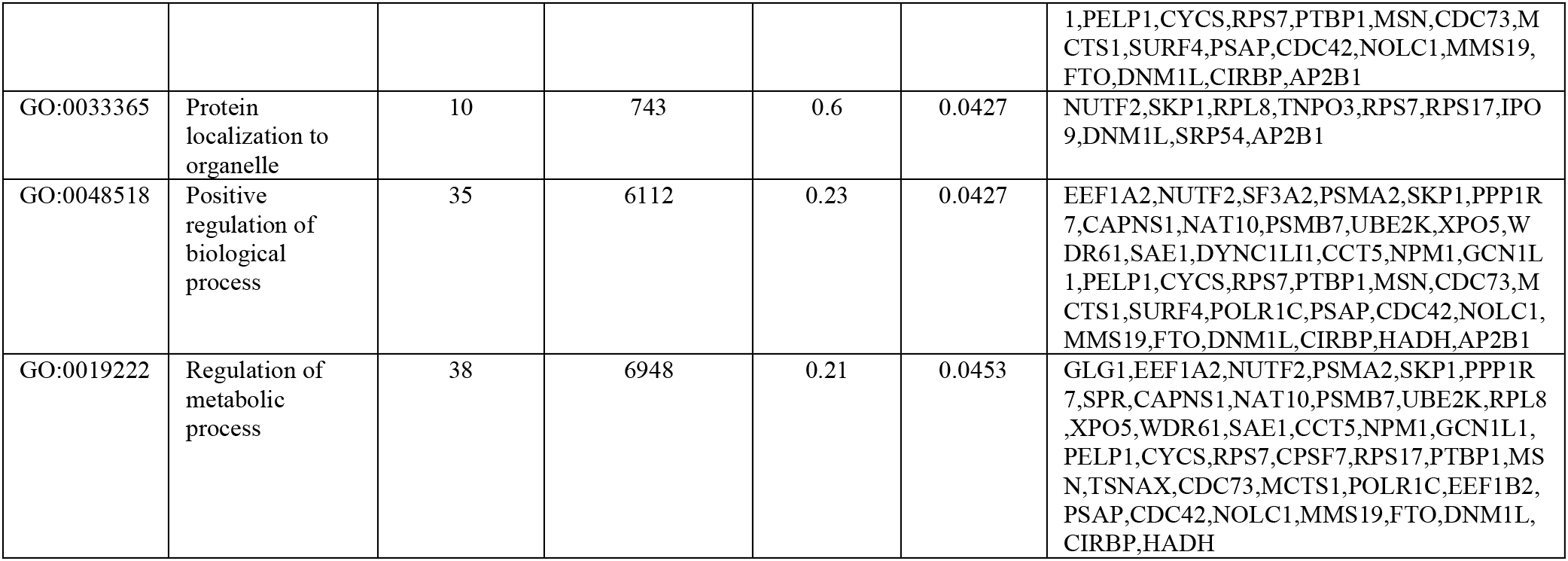
List of the proteins taking part in the interaction network by using those proteins upregulated by PI-7 compound in HEK293T-ACE2 cells. The proteins taking part to the neural network generated through the “Search Tool for Retrieval of Interacting Genes/Proteins” (STRING) database by using as input those proteins upregulated by PI-7 molecule in HEK293T-ACE2 cells after 24 hours treatment are shown. The Term ID, the term description, the observed gene count in the network, the background gene count, the strength, the false discovery rate (FDR) and the protein names are listed.

**Table S7:** List of the proteins taking part in the interaction network by using those proteins downregulated by PI-7 compound in HEK293T-ACE2 cells. The proteins taking part to the neural network generated through the “Search Tool for Retrieval of Interacting Genes/Proteins” (STRING) database by using as input those proteins downregulated by PI-7 molecule in HEK293T-ACE2 cells after 24 hours treatment are shown. The Term ID, the term description, the observed gene count in the network, the background gene count, the strength, the false discovery rate (FDR) and the protein names are listed.

| Term ID | Term description | Observed gene count | Background gene count | Strength | FDR | Matching proteins in the network |
| --- | --- | --- | --- | --- | --- | --- |
| GO:0070972 | Protein localization to endoplasmic reticulum | 8 | 140 | 1.79 | 4.64e-09 | CHMP4B,RPS11,RPS27A,RPS27,RPL23A,BCAP31,RPS20,RPL18 |
| GO:0045047 | Protein targeting to endoplasmic reticulum | 7 | 110 | 1.84 | 4.39e-08 | CHMP4B,RPS11,RPS27A,RPS27,RPL23A,RPS20,RPL18 |
| GO:0006413 | Translational initiation | 7 | 141 | 1.73 | 1.16e-07 | RPS11,RPS27A,RPS27,EIF1AX,RPL23A,RPS20,RPL18 |
| GO:0006612 | Protein targeting to membrane | 7 | 165 | 1.66 | 2.69e-07 | CHMP4B,RPS11,RPS27A,RPS27,RPL23A,RPS20,RPL18 |
| GO:0006614 | SRP-dependent cotranslational protein targeting to membrane | 6 | 96 | 1.83 | 6.50e-07 | RPS11,RPS27A,RPS27,RPL23A,RPS20,RPL18 |
| GO:0019083 | Viral transcription | 6 | 115 | 1.75 | 1.38e-06 | RPS11,RPS27A,RPS27,RPL23A,RPS20,RPL18 |
| GO:0000184 | Nuclear-transcribed mRNA catabolic process, nonsense-mediated decay | 6 | 119 | 1.74 | 1.49e-06 | RPS11,RPS27A,RPS27,RPL23A,RPS20,RPL18 |
| GO:0072657 | Protein localization to membrane | 8 | 495 | 1.24 | 6.70e-06 | CHMP4B,RPS11,RPS27A,BSG,RPS27,RPL23A,RPS20,RPL18 |
| GO:0016032 | Viral process | 9 | 776 | 1.1 | 8.86e-06 | PSMA3,CHMP4B,RPS11,RPS27A,RPS27,RPL23A,BCAP31,RPS20,RPL18 |
| GO:0044265 | Cellular macromolecule catabolic process | 8 | 917 | 0.98 | 0.00035 | PSMA3,CHMP4B,RPS11,RPS27A,RPS27,RPL23A,RPS20,RPL18 |
| GO:0006886 | Intracellular protein transport | 8 | 999 | 0.94 | 0.00062 | CHMP4B,RPS11,RPS27A,RPS27,RPL23A,BCAP31,RPS20,RPL18 |
| GO:0016071 | mRNA metabolic process | 7 | 678 | 1.05 | 0.00065 | RPS11,RPS27A,RPS27,RBMX,RPL23A,RPS20,RPL18 |
| GO:0034613 | Cellular protein localization | 9 | 1610 | 0.78 | 0.0016 | CHMP4B,RPS11,RPS27A,BSG,RPS27,RPL23A,BCAP31,RPS20,RPL18 |
| GO:0044271 | Cellular nitrogen compound biosynthetic process | 8 | 1522 | 0.76 | 0.0095 | RPS11,RPS27A,RPS27,RBMX,EIF1AX,RPL23A,RPS20,RPL18 |
| GO:0034645 | Cellular macromolecule biosynthetic process | 8 | 1592 | 0.74 | 0.0118 | RPS11,RPS27A,RPS27,RBMX,EIF1AX,RPL23A,RPS20,RPL18 |
| GO:0008152 | Metabolic process | 16 | 8298 | 0.32 | 0.0167 | PSMA3,CHMP4B,TXNDC17,RPS11,RPS27A,HIST1H1E,BSG,RPS27,RBMX,EIF1AX,CIAPIN1,RPL23A,RPS20,RPL18,ERH,COX4I1 |
| GO:0034622 | Cellular protein-containing complex assembly | 6 | 816 | 0.9 | 0.0173 | CHMP4B,RPS27A,HIST1H1E,RPS27,RBMX,RPL23A |
| GO:0051641 | Cellular localization | 10 | 2967 | 0.56 | 0.0220 | CHMP4B,RPS11,RPS27A,BSG,RPS27,RPL23A,BCAP31,RPS20,MANF,RPL18 |
| GO:0051649 | Establishment of localization in cell | 9 | 2375 | 0.61 | 0.0247 | CHMP4B,RPS11,RPS27A,RPS27,RPL23A,BCAP31,RPS20,MANF,RPL18 |
| GO:0044237 | Cellular metabolic process | 15 | 7513 | 0.34 | 0.0294 | PSMA3,CHMP4B,RPS11,RPS27A,HIST1H1E,BSG,RPS27,RBMX,EIF1AX,CIAPIN1,RPL23A,RPS20,RPL18,ERH,COX4I1 |
| GO:0010629 | Negative regulation of gene expression | 8 | 2014 | 0.64 | 0.0485 | RPS11,RPS27A,HIST1H1E,RPS27,RBMX,RPL23A,RPS20,RPL18 |

## Notes

### Competing Interest Statement

The authors have declared no competing interest.

### Author Declarations

The Ethical Committee approvals for the COVID19 samples use in this study were as follows: (i) protocol no. 141/20; date: 10 April 2020, CEINGE TaskForce Covid19; Azienda Ospedaliera Universitaria Federico II, Direzione Sanitaria, protocol no. 000576 of 10 April 2020; (ii) protocol no. 157/20; date: 22 April 2020, GENECOVID, with the experimental procedures for the use of SAR-CoV-2 in a biosafety level 3 (BSL3) laboratory were authorized by Ministero della Sanita and Dipartimento Di Medicina Molecolare e Biotecnologie Mediche, University degli Studi di Napoli Federico II and Azienda Ospedaliera Universitaria Federico II, Direzione Sanitaria protocol no. 0007133 of 08 May 2020; (iii) protocol no. 18/20; date: 10 June 2020, Genetics CEINGE TaskForce Covid19; Azienda Ospedaliera Universitaria Federico II, Direzione Sanitaria protocol no. 000576 of 10 April 2020.

